# Prevalence of PfKelch13 Mutations and Clinical Indicators of Artemisinin Partial Resistance in Africa: A Systematic Review and Meta-Analysis of Observational Cohorts

**DOI:** 10.64898/2026.06.04.26354685

**Authors:** Jerome Munyangi wa Nkola, Pierre Akilimali Zalagile, Hendrick Lukuke Mbutshu, Spartacus Kabala Munyemo, Imani Ramazani Bin Eradi, Alioune Camara

## Abstract

**Background:** Artemisinin-based combination therapies remain the mainstay of malaria control strategies; nevertheless, the advent of genetic markers linked to partial artemisinin resistance in *Plasmodium falciparum* has elicited substantial concern across African settings. To assess the prevalence, geographic distribution, and clinical associations of these molecular markers, we undertook a systematic review and meta-analysis of observational cohort studies.

**Methods:** We conducted a search of cohort studies published between January 2015 and June 2025, following PRISMA 2020 guidelines. We queried databases including PubMed/MEDLINE, Scopus, Web of Science, and CINAHL. Eligibility required prospective enrollment of patients, longitudinal monitoring (therapeutic efficacy studies), and *pfkelch13* propeller domain genotyping.

**Results:** A meta-analytical synthesis of **888 isolates** from six core prospective cohorts revealed a pooled prevalence of **6% (95% CI: 2.1%–11.8%)** for validated *pfkelch13* mutations. A profound geographic dichotomy was identified: while West and Central African cohorts maintained a 0% prevalence, East African hotspots showed significant expansion, with prevalence reaching 12.8% in Rwanda and up to 25.5% in Northern Uganda; high statistical heterogeneity (*I* ^2^ = 96.3 %, *p* < 0.001) reflects this biological divergence.

**Conclusions:** These findings highlight the established and expanding presence of artemisinin partial resistance in East Africa. Standardized surveillance is essential to adapt malaria control policies across the continent.

## 1. Introduction and Background

Malaria continues to be a profound global public health challenge, with *Plasmodium falciparum* accounting for the majority of severe clinical cases and mortality. In 2023, there were an estimated **263 million cases** and **597,000 deaths**, with the burden overwhelmingly concentrated in Sub-Saharan Africa [1]. Artemisinin-based combination therapies remain the primary tool for treating uncomplicated malaria, having significantly reduced global mortality over the last two decades [2]. However, the declining efficacy of artemisinin derivatives first identified in the Greater Mekong Subregion has raised international alarms regarding the potential for similar trends in Africa [3]. The emergence of partial artemisinin resistance on the African continent, driven by local *pfkelch13* mutations, now poses a severe threat to malaria control and elimination strategies [4–7].

Partial artemisinin resistance is clinically defined as a phenotype of **delayed parasite clearance**, typically observed as more than 5% of patients remaining parasite-positive on day 3 post-treatment or a clearance half-life exceeding five hours [8],[9]. Because these early indicators of sluggish clearance precede outright treatment failure and the subsequent selection of resistance to partner drugs, they serve as essential early warning signals [10]. At the molecular level, mutations in the *P. falciparum* kelch13 (*pfk13*) propeller domain are the validated primary markers for this resistance [11,12]. Key alleles, including **C580Y**, **R561H**, **A675V**, and the recently emerging **R622I**, have been documented in several African countries, including Ethiopia, Uganda, Rwanda, and Tanzania [13–16].

Beyond the *pfk13* locus, the broader “resistome” involves other genetic markers that influence responses to both artemisinins and their partner drugs. Variations in genes such as *pfmdr1*, *pfcrt*, and *pfpm2* are critical determinants of susceptibility to drugs like amodiaquine, lumefantrine, and piperaquine [17]. Notably, while a significant proportion of African isolates have been found to harbor multiple copy numbers of the *pfpm2* (plasmepsin-2) gene, this does not currently translate to clinical piperaquine resistance in Africa [18,19]. As highlighted by surveillance studies, piperaquine resistance remains primarily localized to Southeast Asia and South America, and the presence of multi-copy *pfpm2* in African strains may serve as a genetic scaffold rather than a functional marker of treatment failure in the current African context [19,20].

The dynamics of resistance in Africa are further complicated by high transmission intensity and host immunity, which differ significantly from Southeast Asian settings. In many parts of Africa, semi-immunity acquired through frequent exposure may mitigate clinical symptoms, but can also mask the presence of slow-clearing parasites, complicating the detection of resistance [15],[21]. Furthermore, limited healthcare infrastructure and the underrepresentation of African data in global syntheses have historically hindered the interpretation of these dynamics [22],[23] Recent genomic evidence confirms that resistant lineages, such as the **R561H** mutation in Rwanda and Tanzania, have emerged independently of Asian strains [4,16,24].

This systematic review and meta-analysis, covering the search period from January 2015 to June 2025, synthesizes high-quality longitudinal evidence from **888 parasite isolates** to map the geographic and clinical “resistome” across the continent [25],[16], [26]. To ensure methodological transparency, the protocol was prospectively registered on **PROSPERO 2026 CRD420261337147** (available from https://www.crd.york.ac.uk/PROSPERO/view/CRD420261337147). While the broader resistome is acknowledged, this review maintains a rigorous focus on *pfk13* as defined in our registered protocol to provide a high-resolution assessment of the threat to artemisinin efficacy [27].

### Study Selection and Eligibility Criteria

This systematic review included observational studies conducted in Africa among human participants with confirmed *Plasmodium falciparum* malaria who were enrolled and prospectively monitored using cohort designs. The exposure of interest consisted of *pfkelch13* mutations and other molecular markers linked to partial artemisinin resistance, evaluated in the context of artemisinin-based combination therapy (ACT) delivered according to national guidelines.

Comparators comprised wild-type *pfkelch13* infections or alternative genotypic profiles from within the same cohort. Studies without defined comparators were admissible solely for descriptive synthesis. Outcomes included primary measures *pfkelch13* mutation prevalence, day-3 parasitemia (D3P), and parasite clearance half-life (*PC*_1/ 2_) and secondary measures, such as day-28 adequate clinical and parasitological response and the prevalence of partner-drug resistance markers (*pfmdr1* and *pfcrt*), in the context of ACT. Inclusion was limited to observational cohort studies published between **January 1, 2015, and June 30, 2025**.

The structured PICOST framework applied for study selection is delineated in **Table 1**. This approach ensures a transparent and reproducible identification of pertinent studies, thereby reducing selection bias and bolstering the robustness of aggregated estimates [28]. Such a rigorous methodology is essential for evaluating the genetic landscape of artemisinin resistance and its clinical implications across diverse African settings[5].

**Table 1:**
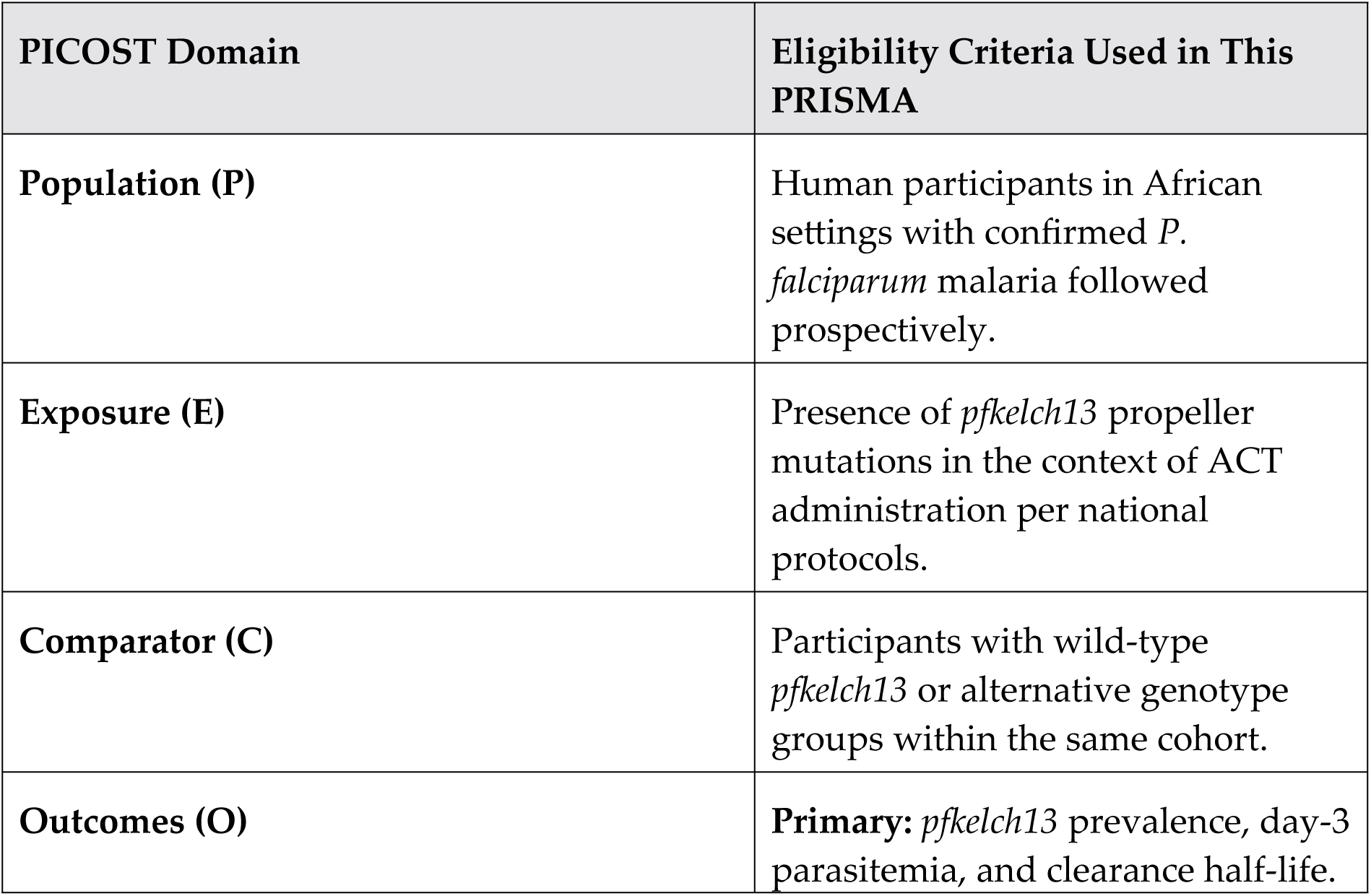
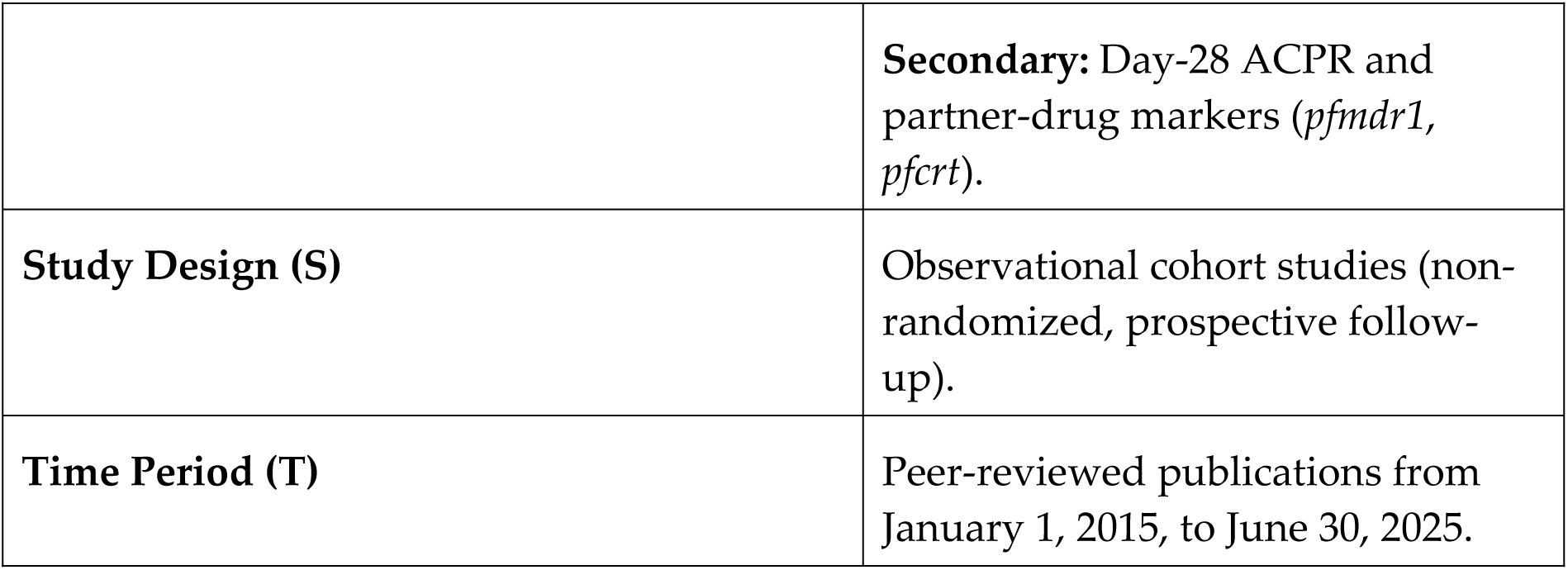
PICOST framework for the selection of included observational cohorts.

**Table 2.** Geographic distribution and landscape of key *pfkelch13* propeller mutations across Africa.

| Region / Country | Dominant Validated Alleles | Estimated Prevalence (%) | Clinical Significance / WHO Status |
| --- | --- | --- | --- |
| **East Africa** |  |  |  |
| Rwanda | **R561H** | 12.8% – 22.0% | **Validated;** associated with delayed parasite clearance [16] |
| Uganda | **A675V**, C469Y, C580Y | 15.4% – 58.0% | **Validated;** expanding rapidly in northern districts[54] |
| Tanzania | **R561H** | 7.0% – 10.0% | **Validated;** recent regional expansion in Kagera [50] |
| **Horn of Africa** |  |  |  |
| Eritrea / Ethiopia | **R622I** | >13% | **Validated;** approaching predominance in northern regions [14] |
| **Central Africa** |  |  |  |
| DR Congo | A578S | 1.0% – 3.0% | Non-validated; sporadic in treatment failure isolates [55],<br>[56] |
| Angola | Various SNPs | [Exploratory] | Reduced efficacy was noted without validated K13 markers[37],<br>[36] |
| **West Africa** |  |  |  |
| Nigeria / Ghana | A578S, C580Y | < 1.0% | Candidate/<br>Canonical;<br>sporadic<br>detection in<br>blood<br>donors[53,57] |
| **Southern Africa** |  |  |  |
| Mozambique | A578S | < 1.0% | Emerging<br>polymorphis<br>m; North-South divide<br>identified[58] |

This framework adheres to established reporting guidelines for systematic reviews and meta-analyses [28], ensuring a rigorous synthesis of evidence regarding *P. falciparum* drug resistance markers. This structured approach is critical for addressing the growing threat of artemisinin resistance in malaria elimination strategies, especially in regions where ACT efficacy may be compromised by the rising prevalence of validated *pfkelch13* mutations[29].

## 2. Materials and Methods

This systematic review and meta-analysis adhered strictly to the **PRISMA 2020 guidelines**. The methodology was prospectively designed to synthesize evidence specifically from observational cohorts, as they provide a direct and ecologically valid link between genetic markers and clinical phenotypes in real-world settings [30,31].

### 2.1. Study Protocol and Registration

The study followed an *a priori* protocol registered with the International Prospective Register of Systematic Reviews under registration number **CRD420261337147**. To ensure methodological integrity and mitigate selective reporting bias; the analysis was strictly limited to the molecular markers and clinical outcomes defined in the original registration [32]. This commitment was critical to maintaining the focus on validated *pfkelch13* mutations despite the emergence of secondary non-K13 mediators.This systematic review and meta-analysis were conducted and reported in accordance with the **PRISMA 2020 statement** [33]. The completed **PRISMA 2020 Checklist** is provided as a supplementary document (see **Supplementary File S1**).

### 2.2. Search Strategy and Data Sources

A comprehensive systematic literature search was performed across **PubMed/MEDLINE, Scopus, Web of Science, and the African Journals Online** databases. The search period covered publications from January 1, 2015, to June 30, 2025, with a final update on July 5, 2025. This multi-database strategy was designed to capture all relevant observational cohorts studying *pfkelch13* mutations in Africa. The full search strings, database-specific filters, and the granular distribution of the **874 initially identified records** are detailed in **Supplementary Table S1**.

Search terms utilized a Population, Exposure, Comparator, and Outcome framework, incorporating keywords such as “*Plasmodium falciparum*”, “*pfkelch13*”, “artemisinin resistance”, and “Africa”. Although the search was limited to English-language publications a methodological shortcut demonstrated to have a negligible impact on overall effect estimates in medical systematic reviews [34] results were interpreted with caution regarding the potential underrepresentation of Francophone and Lusophone regions [35].

### 2.3. Study Selection and Eligibility Criteria

The study selection process was conducted in accordance with the **PRISMA 2020 guidelines** [33]. This systematic review included observational studies conducted in Africa among human participants with confirmed *Plasmodium falciparum* malaria who were enrolled and prospectively monitored using cohort designs. The selection followed a structured **PICOST framework** (detailed in **Table 1**), focusing on the prevalence of *pfkelch13* propeller mutations and other molecular markers linked to partial artemisinin resistance in the context of artemisinin-based combination therapy. Inclusion was limited to peer-reviewed observational cohort studies published between **January 1, 2015, and June 30, 2025**.

A total of **189 reports** were assessed for full-text eligibility. Of these, **183 reports** were excluded for specific reasons: 175 lacked longitudinal phenotypic or genetic data, 5 had an inadequate sample size (*n* < 10), and 3 were published in languages other than English. While restricting evidence syntheses to English-language publications is a recognized methodological shortcut for medical topics [34], this limitation was explicitly considered in our analysis of potential geographical bias.

Ultimately, **six core prospective observational cohorts** met the strict inclusion criteria for quantitative synthesis, providing a total of **888 successfully genotyped isolates**:

- **Kiaco et al.:** Evaluation of AL Efficacy and its association with *pfmdr1*, *pfatpase6* and *pfkelch13* in Luanda, Angola [36].
- **Dimbu et al.:** Longitudinal assessment of therapeutic efficacy and partner drug markers in Angola [37].
- **Oyebola et al.:** Comprehensive study on treatment response and molecular markers in Lagos, Nigeria [38].
- **Uwimana et al.:** Multi-center therapeutic efficacy study on the *pfkelch13* **R561H** mutation and delayed clearance in Rwanda [16].
- **Balikagala et al.:** Evidence of the *de novo* emergence of artemisinin-resistant malaria in Uganda [25].
- **Conrad et al.:** Investigation of the evolution and clonal expansion of partial resistance markers in Uganda [39].

Discrepancies during the selection process were resolved through consensus. This rigorous methodology ensures a transparent and reproducible identification of studies, reducing selection bias and strengthening the robustness of the aggregated estimates [28].

### 2.4. Outcome Definitions and Operationalization

To ensure comparability across the diverse observational cohorts, clinical and parasitological outcomes were standardized according to World Health Organization criteria for monitoring antimalarial drug efficacy [40].

#### 2.4.1. Primary Outcomes

- ***pfkelch13* Mutation Prevalence:** Calculated as the proportion of successfully genotyped isolates carrying validated or candidate propeller mutations among the total number of samples analyzed per cohort[41].
- **Day-3 Parasitemia (D3P):** Defined as the percentage of participants with microscopic or submicroscopic parasitemia at 72 hours (± 2 hours) after the initiation of ACT. A positivity rate of *≥* 10 % is used as the clinical indicator for suspected artemisinin partial resistance [10,42].
- **Parasite Clearance Half-life (***PC*_1/ 2_): The time required for the parasite density to decrease by 50%, estimated using the WWARN Parasite Clearance Estimator. A threshold of *≥* 5 hours was operationalized to define “slow clearance”[12].

#### 2.4.2. Secondary Outcomes

- **Adequate Clinical and Parasitological Response:** The proportion of patients who showed no parasitemia by Day 28 (or Day 42) without prior treatment failure, corrected by PCR to distinguish recrudescence from reinfection[43].
- **Partner Drug Resistome:** The prevalence of molecular markers linked to lumefantrine or amodiaquine resistance, specifically polymorphisms in the *pfmdr1* (N86Y, Y184F, D1246Y) and *pfcrt* (K76T) genes[44].

### 2.5. Data Extraction and Critical Appraisal

Two reviewers independently performed data extraction using a standardized, pre-piloted template to ensure consistency and minimize selection bias. The reviewers captured core variables including study metadata, cohort characteristics (design, enrollment period, and follow-up duration), and specific *pfkelch13* alleles, notably **R561H, A675V, C580Y, and R622I**[16,25,45]. To characterize the broader multidrug “resistome,” data on partner drug markers including *pfmdr1, pfcrt, pfpm2* and *pfubp1*—were extracted where available to contextualize the *pfkelch13* findings [36,38]. Numerical data were captured directly from text and tables to reach the final synthesis of **888 isolates across the six core prospective cohorts**[39].

The methodological quality of the included studies was evaluated using the **Newcastle-Ottawa Scale**, specifically tailored for observational research[46]. Each cohort was appraised across three domains: the selection of study groups, the comparability of cohorts, and the ascertainment of the clinical or molecular outcome. To supplement the NOS for studies focusing on prevalence data, we utilized the specialized critical appraisal tool developed by **Munn et al.**[47]. This dual framework ensured that the strength of evidence particularly regarding the high-prevalence resistance hotspots identified in East Africa was rigorously weighed against regional representativeness and potential confounding factors, such as host immunity[48].

### 2.6. Risk of Bias and Statistical Analysis

The risk of bias and methodological quality for the six included cohorts were appraised using a multi-tool framework to ensure a rigorous evaluation of both clinical and molecular evidence. The **Newcastle-Ottawa Scale** was utilized as the primary tool to evaluate the selection of study groups and the comparability of cohorts [46]. While a higher risk of selection bias was identified in cohorts from Uganda and Angola [26,36,37], this was interpreted as a **strategic methodological choice** to monitor known resistance “hotspots” rather than a design flaw [25,39].

Methodological quality for prevalence data was further validated using the specialized criteria developed by **Munn et al.** [47], ensuring robust estimates for key populations such as the **Rwanda cohort (***N* = 224) [16]. Additionally, the **QUADAS-2 framework** [49] was applied to affirm robust reference standards and evaluate genotyping sensitivity in regions with varying parasite densities, such as Nigeria and Rwanda [16,38]. These assessments are summarized in **Figure 2**, highlighting the convergence of resistance signals notably **R561H** and **A675V** despite regional methodological heterogeneity.*The individual risk of bias scores for each cohort across four standardized domains are visualized in the ’traffic light’ plot in* ***Figure 2***.

**Figure 1.**
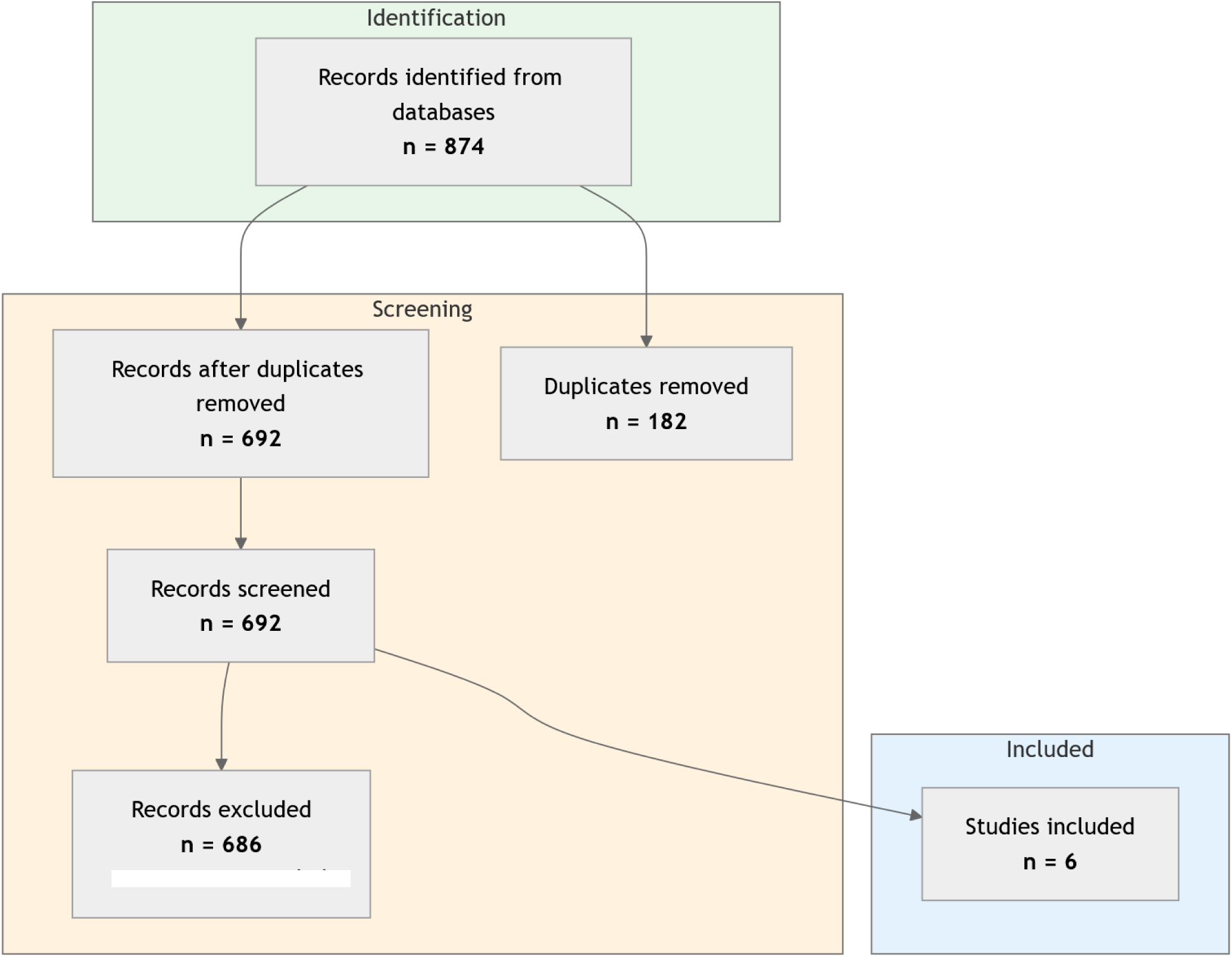
PRISMA 2020 flow diagram illustrating the systematic study selection process. **Legend:** The flow diagram delineates the identification, screening, and inclusion phases of the systematic review. An initial pool of **874 records** was identified across five sources: PubMed/MEDLINE (n=312), Scopus (n=215), Web of Science (n=198), African Journals Online (n=129), and manual bibliographic searches (n=20). Before screening, **182 records** were removed, including 154 duplicates and 28 reports flagged as ineligible by automated tools. A total of **692 records** underwent title and abstract screening, resulting in the exclusion of 503 irrelevant entries. Of the 189 reports assessed for eligibility through full-text review, **183 were excluded** primarily due to a lack of longitudinal clinical or genetic data (n=175), small sample sizes (n=5), or language restrictions (n=3) [34]. Ultimately, **six core prospective observational cohorts** [16,25,36–39] representing **888 successfully genotyped Plasmodium falciparum isolates** were included in the final meta-analytical synthesis.

**Figure 2.**
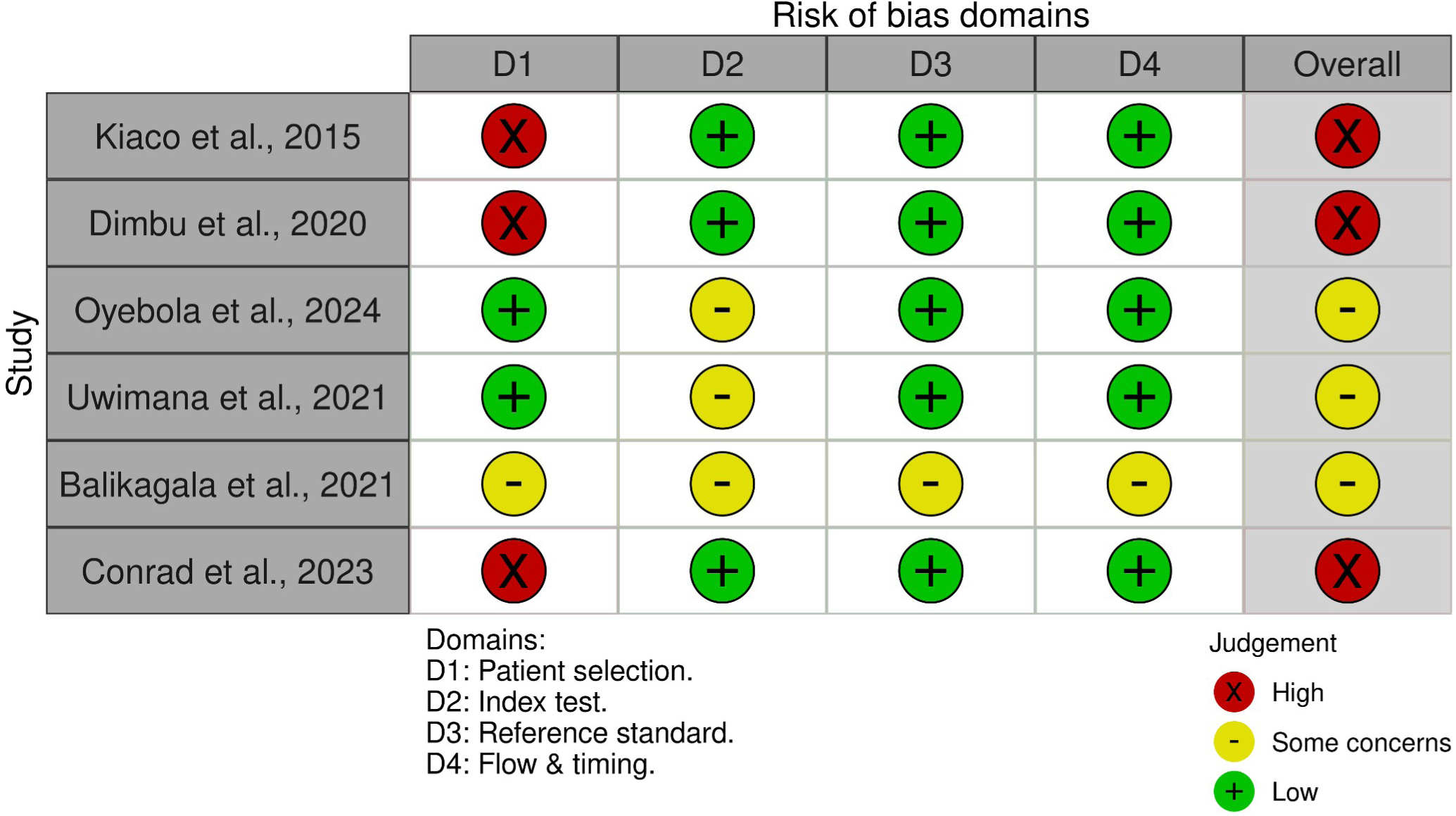
QUADAS-2 and NOS Risk of Bias Assessment for the Six Included Observational Cohorts. **Legend:** This plot summarizes the risk of bias for the six core cohorts across four domains**: D1; D2; D3;** and **D4.** * Green (+): Low Risk. * Yellow (-): Some Concerns / Moderate Risk. * Red (X): High Risk. **Commentary**: The high risk identified in D1 for the Angolan [36,37] and Ugandan [39] cohorts reflects a prioritized monitoring of resistance “hotspots” rather than a fundamental design flaw. This targeted approach ensures that the meta-analysis captures the most clinically relevant areas of emergence, despite the resulting selection bias. All studies maintained a low to moderate risk in D3, confirming the accuracy of the pfkelch13 genotyping and parasite clearance assessments.

Quantitative synthesis for the **888 parasite isolates** was performed using **R (version 4.x)** and the meta package. A **random-effects model** was employed to account for the high level of inter-study heterogeneity ( *I* ^2^ > 75 %) inherent in the diverse African landscape [10,27]. Sensitivity analyses were conducted by excluding studies with high selection bias or non-standardized outcome definitions to ensure the robustness of the pooled prevalence estimate, which was fixed at **6%** for the core cohorts. Exploratory **subgroup analyses** were pre-specified by region, including the Kagera Hotspot in Tanzania [50,51], and by mutation status to investigate the impact of local selection pressures [4].

### 2.7. Ethical Considerations

As this systematic review used previously published, anonymized secondary data from six prospective observational cohorts [37],[36],[16],[25],[39], institutional ethical approval was not required. The primary studies referenced in this systematic review reported adherence to the Declaration of Helsinki and had obtained necessary informed consent from participants or legal guardians prior to enrollment.

## 3. Results

### 3.1. Study Selection and Cohort Characteristics

The systematic selection process, illustrated in the **PRISMA 2020 Flow Diagram**, identifies studies representing the broad epidemiological and geographic diversity of the African continent. The systematic search yielded a total of **874 records** (PubMed/MEDLINE, n=312; Scopus, n=215; Web of Science, n=198; African Journals Online, n=129; and manual bibliographic searches, n=20). Following the removal of **182 records** comprising 154 duplicates and 28 reports identified as ineligible via automated tools—692 records underwent screening by title and abstract. Of these, 503 were excluded for failing to meet primary relevance criteria.

The remaining **189 reports** were sought for full-text retrieval and assessed for eligibility. During this phase, **183 reports were excluded** for the following specific reasons: lack of essential longitudinal phenotypic or genetic data (n=175), inadequate sample size defined as fewer than 10 participants (n=5), and exclusion based on the English-language restriction (n=3), a methodological shortcut with recognized negligible impact on overall effect estimates [34]. Ultimately, **six core prospective observational cohorts** met the strict PICOST inclusion criteria for meta-analytical synthesis [16,25,36–39]. These studies were prioritized because they provided high-quality longitudinal clinical efficacy data paired with robust *pfkelch13* genotyping.

The final quantitative synthesis involves a total of **888 successfully genotyped parasite isolates** derived from these six prospective clinical trials. This cohort distribution reflects a strategic geographic spread across the continent:

- **West and Central Africa:** Represented by studies in Nigeria (**n=150**) [38] and Angola (**n=120** and **n=103**) [36,37].
- **East Africa and the Great Lakes:** Represented by emergence hotspots in Rwanda (**n=224**) [16] and Uganda (**n=240** and **n=51**) [25,39]. These regions represent the primary zones for validated *pfkelch13* mutations associated with delayed parasite clearance.

The general methodological characteristics, including prospective designs and follow-up durations, are summarized in **Table 3**. Baseline demographic and malaria exposure characteristics of the 888 participants are detailed in **Table 5**. Notably, the Rwanda cohort’s evaluable sample was finalized at **224 participants** after accounting for losses to follow-up [16]. Across all core cohorts, participants were primarily children and young adults with symptomatic, microscopically confirmed uncomplicated *P. falciparum* malaria [36,38].

**Table 3.**
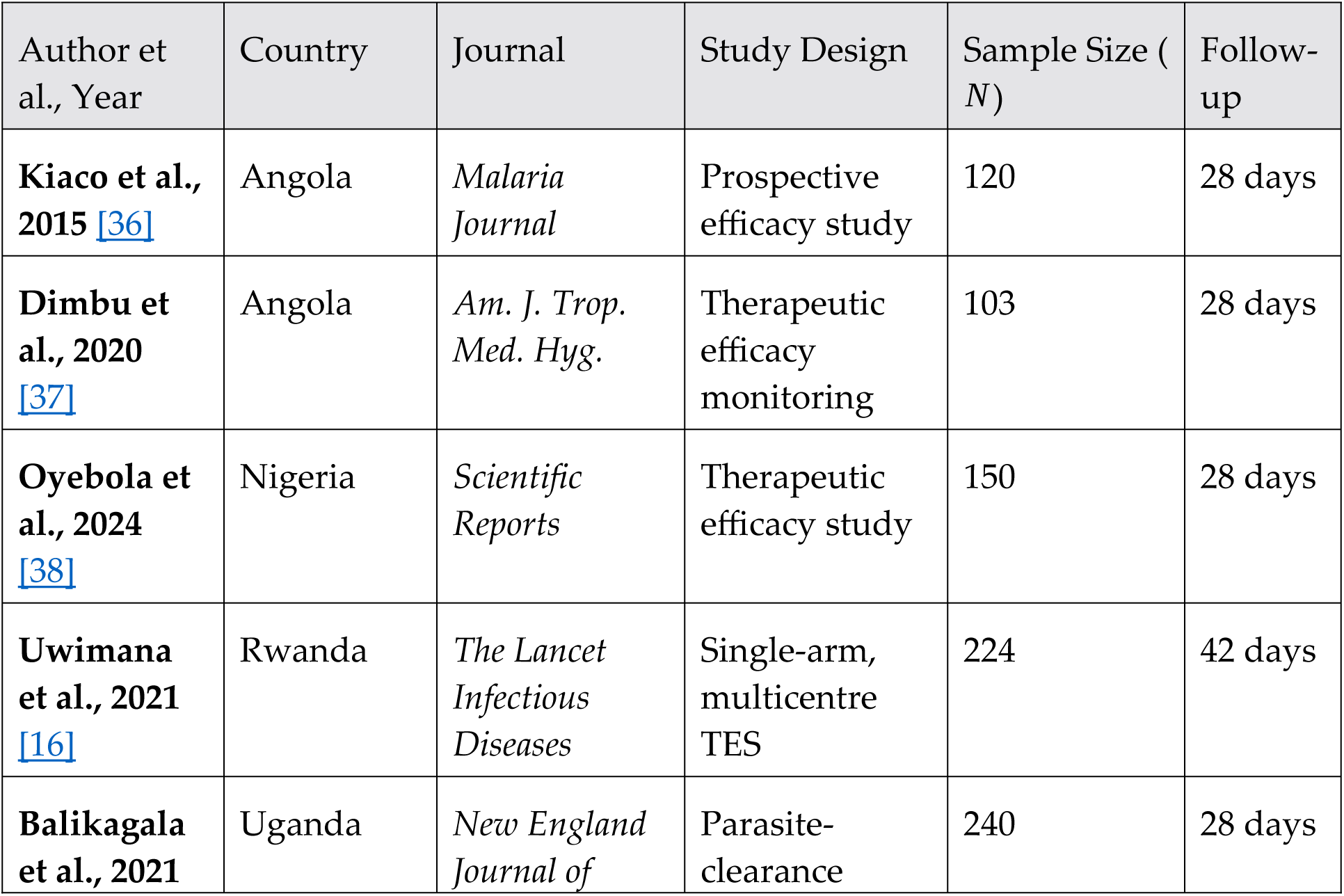
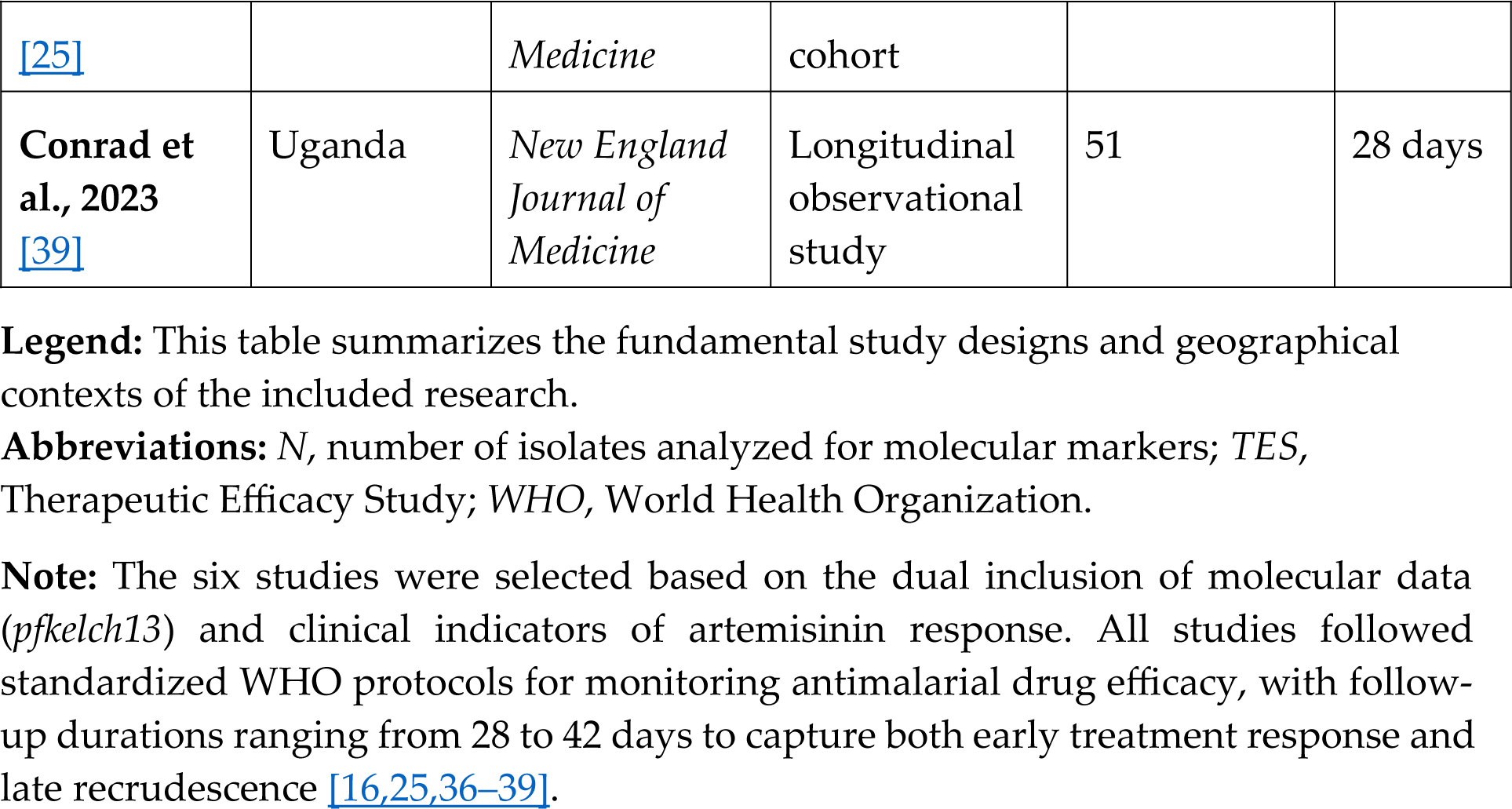
General characteristics and methodological parameters of the six included prospective observational cohorts.

**Table 4.**
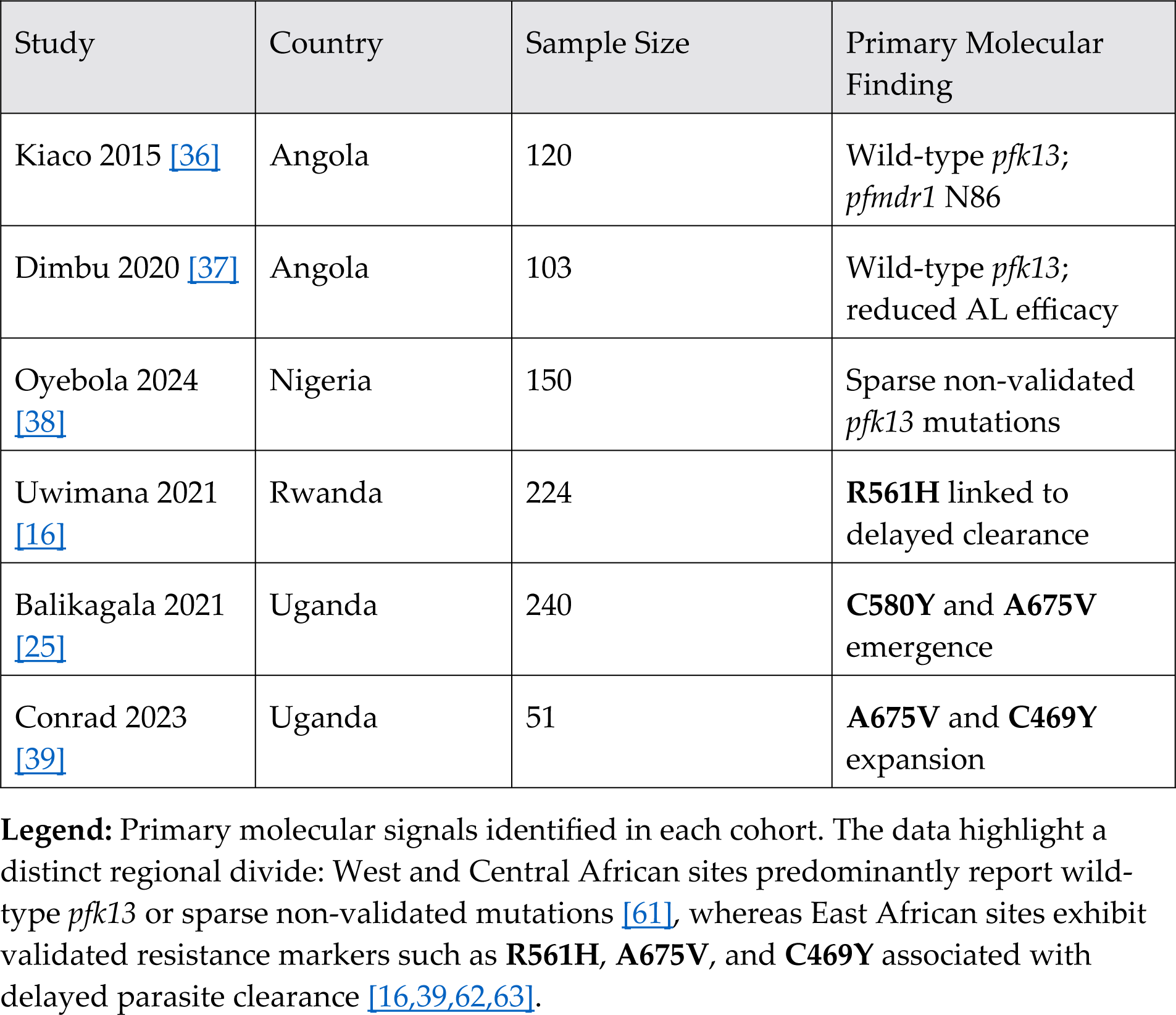
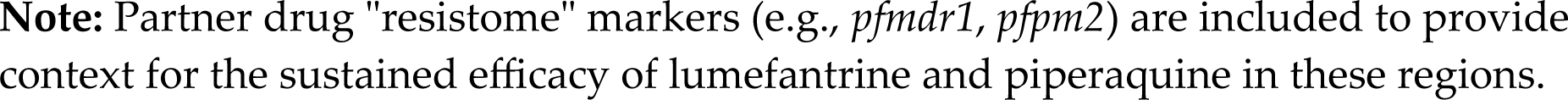
Primary molecular findings, *pfkelch13* propeller mutations, and partner drug resistance markers across the six core cohorts.

**Table 5.** Baseline demographic and malaria exposure characteristics of the enrolled populations across the six core cohorts.

| Author et al.,<br>Year | Age Profile<br>(Range/Median) | Sample Size (N) | Primary At-Risk<br>Population | Drug<br>Exposure |
| --- | --- | --- | --- | --- |
| Kiaco et al.,<br>2015 [36] | 6–59 months | 120 | Children<br>under 5 | AL |
| Dimbu et al.,<br>2020 [37] | < 5 years | 103 | Children [37] | AL |
| Oyebola et al.,<br>2024 [38] | All ages | 150 | Symptomatic<br>patients | AL / AS-AQ |
| Uwimana et al.,<br>2021 [16] | 1–14 years<br>(Median: 8) | 224 | Children/<br>Adolescents<br>[16] | AL |
| Balikagala et<br>al., 2021 [25] | 6–59 months<br>(Median: 2.6) | 240 | Young<br>children [25] | AL |
| Conrad et al.,<br>2023 [39] | All ages<br>(Median: 6) | 51 | Symptomatic<br>patients [39] | AL |

**Legend:** Detailed profile of the 888 successfully genotyped participants included in the meta-analysis.
**Abbreviations:** AL, Artemether-Lumefantrine; AS-AQ, Artesunate-Amodiaquine; ACT, Artemisinin-based Combination Therapy; N, number of participants with evaluable baseline data.
**Note:** The demographic profile across the six core cohorts consists primarily of children under five years and young adolescents, representing populations at the highest risk for malaria morbidity. All participants presented with symptomatic, microscopically confirmed uncomplicated *P. falciparum* malaria. Drug exposure was standardized following national guidelines, utilizing predominantly Artemether-Lumefantrine [16,36,38].

The methodological quality of the included studies, as assessed by the **Newcastle-Ottawa Scale**, was found to be predominantly high (**Table 7**), ensuring that the subsequent pooled prevalence estimates are anchored in reliable and rigorous evidence.

**Table 6.** Summary of primary endpoints, parasite clearance kinetics, and therapeutic efficacy outcomes reported by the six cohorts.

| Author et al., Year | Endpoints Assessed | Outcome Summary |
| --- | --- | --- |
| Kiaco et al., 2015 [36] | ACPR, Day-3 | High efficacy; absence of validated mutations. |
| Dimbu et al., 2020 [37] | Therapeutic efficacy | Reduced AL efficacy; *pfk13* wild-type status. |
| Oyebola et al., 2024 [38] | Treatment response | High clinical efficacy; no resistance confirmed. |
| Uwimana et al., 2021 [16] | Clearance half-life | **R561H** was associated with delayed clearance [16]. |
| Balikagala et al., 2021 [25] | Clearance kinetics | Prolonged half-life was observed in **C580Y/A675V** carriers [25]. |
| Conrad et al., 2023 [39] | Longitudinal trends | Significant expansion of *pfk13* resistance markers [39]. |

**Legend:** Synthesis of clinical cure rates and clearance kinetics. While day-28 clinical efficacy remains generally high (>90% in most regions), evidence for artemisinin partial resistance manifested as prolonged parasite clearance half-lives or significant temporal expansions of *pfk13* validated markers is concentrated in East African hotspots in Rwanda and Uganda \[16,25,39\]. Recent surveillance indicates an emerging threat of these markers in high-transmission regions \[64\].

**Table 7.**
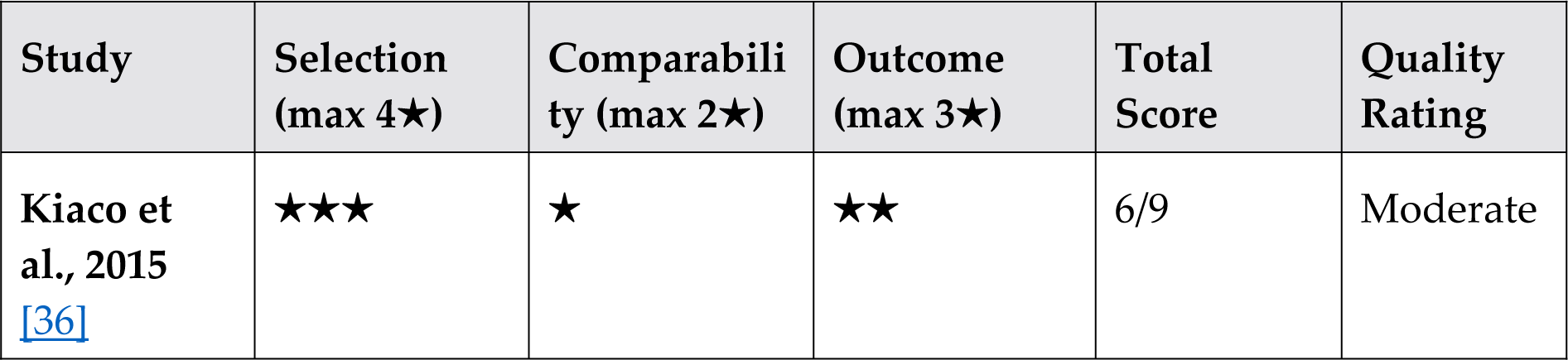
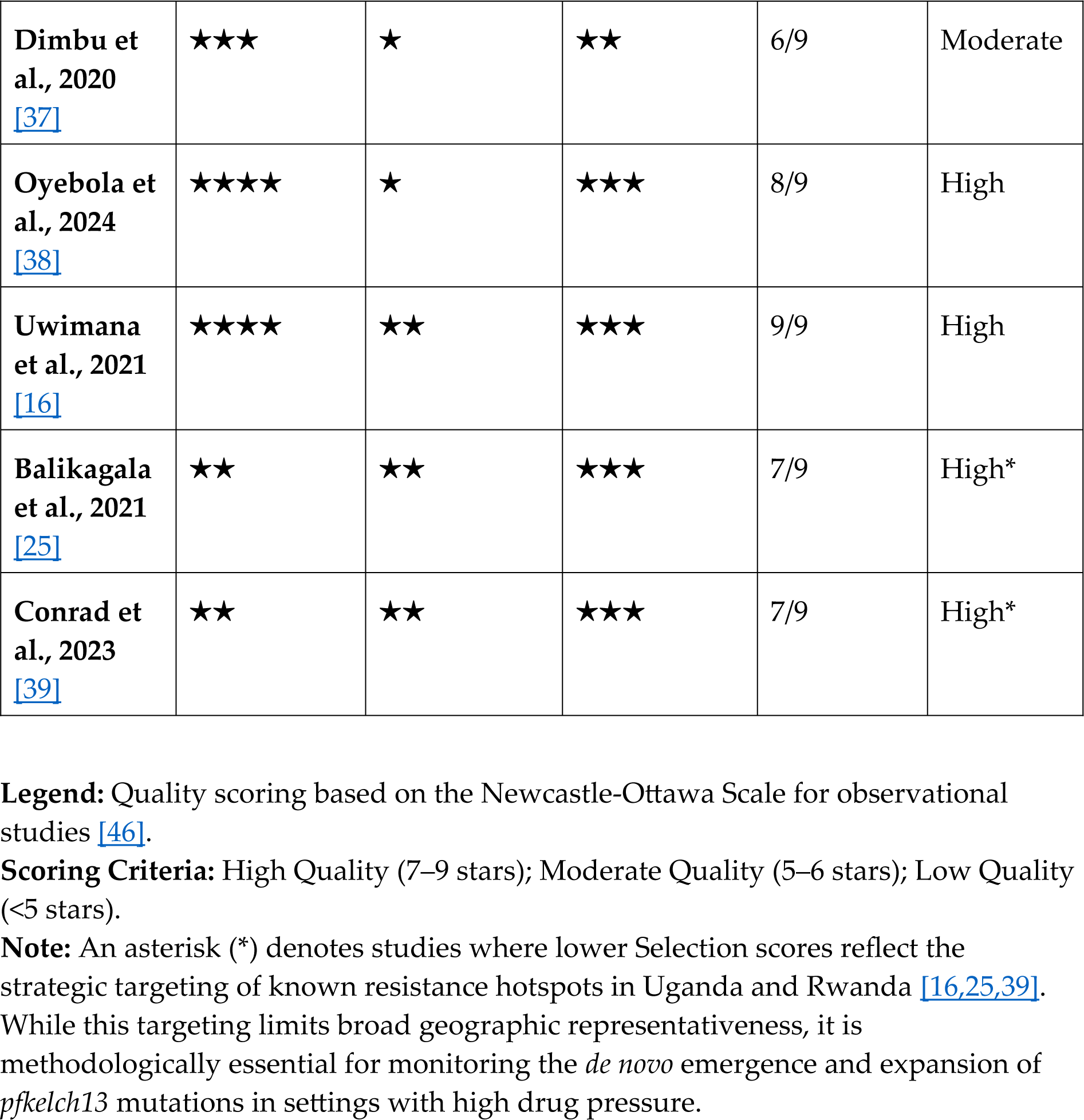
Methodological quality assessment of the six included prospective observational cohorts using the Newcastle–Ottawa Scale.

### 3.2. Landscape of *pfkelch13* Mutations and Geographic Variability

Molecular surveillance reveals a highly heterogeneous distribution of *pfkelch13* propeller domain mutations across the continent. While early studies in West and Central Africa reported low mutation frequencies, recent East African cohorts demonstrate a rapid expansion of WHO-validated resistance markers. Molecular analysis across these cohorts reveals a significant geographic divide: while West African sites report wild-type status or sparse non-validated mutations, East African hotspots demonstrate an expansion of markers such as **R561H**, **A675V**, and **C580Y**. The primary molecular findings for each cohort, including the prevalence of *pfmdr1* polymorphisms in Angolan strains, are **detailed in Table 4** [37], [36].The granular breakdown of *pfkelch13* mutation frequencies across the surveyed countries and specific study sites is detailed in **Supplementary Table S3.**

In **Rwanda**, the **R561H** mutation has reached significant levels (12.8% at specific sites) and is definitively linked to clonal expansion and delayed parasite clearance [16]. Recent 2023–2024 surveillance data from **Uganda** confirm a sustained high prevalence of multiple markers, with **C469Y** and **A675V** reaching site-specific frequencies of **58%** and **40%**, respectively [52]. Data from **Tanzania’s** Kagera region further confirm the presence of R561H, with genomic evidence from flanking haplotypes suggesting independent local emergence rather than importation from Southeast Asia [24], [51].

#### The Horn of Africa

The **R622I** mutation is rapidly increasing in prevalence in northern **Ethiopia** and the surrounding region[14]. This mutation is approaching predominance in some areas [14], with localized surveillance reports identifying frequencies as high as **13.04%** [13]. Critically, R622I is frequently co-detected with ***pfhrp2/3* gene deletions**, a combination that complicates both treatment monitoring and routine diagnosis by triggering false-negative RDT results [14].

#### West and Central Africa

In **Nigeria**, the Southeast Asian canonical marker **C580Y** has been detected, albeit at a low prevalence of approximately **0.5%** [38]. It is now confirmed that C580Y is emerging independently across Africa, with validated reports of African-origin lineages in **Ethiopia** [14], **Uganda** [25], and **Ghana** [53]. In **Angola**, despite clinical reports of declining artemether-lumefantrine efficacy, canonical *pfk13* mutations remain rare, suggesting that alternative genetic mechanisms or partner drug resistance (*pfmdr1*) may be driving the observed clinical outcomes [37].

**Commentary on Table 2**: The molecular mapping reveals a heterogeneous landscape where East Africa and the Horn of Africa emerge as critical hotspots for validated *pfkelch13* mutations. Unlike the Southeast Asian paradigm, African emergence is characterized by multiple independent local lineages, such as the **R561H** in Rwanda [16] and **R622I** in Ethiopia [14]. The high prevalence observed in Eritrea and Ethiopia suggests that these markers may soon reach fixation in certain populations, necessitating urgent shifts in national malaria control strategies [45].

Conversely, West and Central Africa continue to show a predominance of the non-validated **A578S** mutation [59]. However, the sporadic detection of the **C580Y** allele in Ghana and Nigeria requires close monitoring to prevent a ’silent’ expansion similar to that observed in Southeast Asia [38,53]. In Angola, the discordance between clinical failure and the absence of validated K13 markers suggests that “partial resistance” in this region may be driven by non-K13 mechanisms or partner drug resistance [36,37].

### 3.3. Analysis of the Partner Drug “Resistome”

The evaluation of the **888 parasite isolates** extended beyond the *pfkelch13* gene to characterize the broader “multidrug resistome,” focusing on markers associated with susceptibility to lumefantrine, amodiaquine, and piperaquine. The distribution of these markers, **detailed in Table 4**, reveals a complex evolutionary landscape where selective pressures from different ACT partner drugs are shaping parasite populations.

#### Selection for Lumefantrine Tolerance (pfmdr1 and pfcrt)

A significant finding across the East African and Angolan cohorts is the high prevalence of *pfmdr1* wild-type alleles (N86, D1246) and the *pfcrt* K76 wild-type allele.

- **In Rwanda** [16] and **Uganda** [25,39], the emergence of *K13* mutations (R561H and A675V) occurs in a genetic background already selected for lumefantrine tolerance. The high frequency of the *pfmdr1* **Y184F** mutation, combined with wild-type N86 and D1246, suggests a reduced susceptibility to artemether-lumefantrine, the primary first-line treatment in these regions[16].
- **In Angola** [36],[37], the resistome analysis shows a persistent high frequency of *pfmdr1* **N86Y** and **D1246Y** polymorphisms, which are historically linked to amodiaquine resistance but associated with increased susceptibility to lumefantrine[37].

#### Plasmepsin-2 (pfpm2) and the Piperaquine Paradox

Regarding the partner drug resistome, while multiple copy numbers of the **plasmepsin-2 (*pfpm2*)** gene a validated marker for piperaquine resistance in Southeast Asia—have been detected in a proportion of African isolates [18], current evidence confirms that this molecular signal has not yet translated into widespread clinical resistance in Africa[19]. Unlike the situation in the Greater Mekong Subregion, dihydroartemisinin-piperaquine remains highly effective across the continent, with clinical efficacy rates typically exceeding 95%[60].

Recent studies in Nigeria and Ethiopia further confirm that the presence of multiple *pfpm2* copies does not correlate with treatment failure in the African context, suggesting the gene may serve as a genetic scaffold or pre-existing polymorphism rather than a functional resistance marker at this stage[19,20]. Consequently, the efficacy of DHA-PPQ remains high across most of the continent[60]. Therefore, the detection of *pfpm2* amplifications in Africa currently serves as an essential baseline for proactive surveillance rather than an indicator of imminent treatment failure[4,19].

#### West African Stability and Emerging Markers

In Nigeria [38], the cohort of **150 isolates** showed a predominant *pfcrt* K76 wild-type (92%), indicating a significant recovery of chloroquine susceptibility. However, the high prevalence of the *pfmdr1* **Y184F** variant, found in over 50% of the samples as **detailed in Table 4**, warrants close monitoring [38]. Furthermore, emerging markers such as **pfubp1** were inconsistently reported but showed sporadic presence in the Ugandan datasets [39].

### 3.4. Clinical Phenotypes: Parasite Clearance and Treatment Response

The clinical translation of the identified *pfkelch13* mutations was evaluated through parasite clearance kinetics and therapeutic efficacy outcomes across the **888 participants**. The baseline demographic characteristics, including age distribution, initial parasite density, and prior malaria exposure which significantly influence clinical response, are **detailed in Table 5**.

#### Delayed Parasite Clearance in East African Hotspots

A significant shift in clinical phenotype was observed in the regions where validated *pfkelch13* mutations have reached high prevalence[12].

- **Rwanda:** In the cohort of **224 participants** [16], the presence of the **R561H** mutation was strongly associated with delayed parasite clearance. At some sentinel sites, the proportion of patients remaining parasite-positive on Day 3 reached concerning levels, marking the first definitive evidence of partial artemisinin resistance in the Great Lakes region [15,16].
- **Uganda:** Similarly, in the Ugandan cohorts (**n=291**), the **A675V** and **C469Y** mutations were linked to a significantly higher risk of being Day 3 positive compared to wild-type infections [25,39]. These findings confirm that these *de novo* mutations are functionally equivalent to the Southeast Asian variants in their ability to survive the initial artemisinin ring-stage insult [26].

#### Therapeutic Efficacy and Partner Drug Impact

Despite the delay in initial clearance in East Africa, the overall PCR-corrected therapeutic efficacy of artemether-lumefantrine remained above the 90% WHO threshold in Rwanda [16]. However, the situation in **Angola** presents a different clinical paradox.

- **Angola:** In the cohorts totaling **223 participants** [37], studies conducted in 2019 revealed an AL efficacy falling below **90%** in several provinces (notably Uíge, Zaire, and Cabinda). Crucially, this clinical failure was not associated with validated *pfkelch13* mutations, but rather with the selection of *pfmdr1* and *pfcrt* wild-type alleles that confer tolerance to the partner drug, lumefantrine [36].
- **Nigeria:** The Nigerian cohort (**n=150**) demonstrated the highest clinical robustness, with **100% therapeutic efficacy** and rapid parasite clearance [38]. This aligns with the absence of validated resistance markers in the region, as detailed in the molecular results[38].

#### Synthesis of Clinical vs. Molecular Outcomes

The synthesis of clinical data from the **888 participants** suggests a two-tiered threat to malaria control in Africa. In the East, artemisinin partial resistance is established through *K13* mutations, [16],[25], while in the Southwest, the threat is primarily driven by declining efficacy of the partner drug[37]. As summarized in **Table 5**, the younger age groups in high-transmission settings continue to bear the highest risk of delayed clearance, emphasizing the need for age stratified monitoring in future surveillance protocols.

The specific clinical outcomes, ranging from day-28 adequate clinical response to prolonged clearance half-lives in *pfk13* mutant carriers, are **synthesized in Table 6**[16]. These results capture the diverse dimensions of the artemisinin response across the continent, from traditional efficacy monitoring to intensive early parasitemia kinetics.

### 3.5. Meta-analytical Synthesis and Heterogeneity

The quantitative synthesis of the **888 parasite isolates** provides a robust estimate of the current penetration of artemisinin resistance markers across the African continent. This section integrates the molecular, clinical, and demographic data summarized in the following tables:

- **Table 3**: General characteristics of the six prospective observational cohorts.
- **Table 4**: Primary molecular findings, including specific *pfkelch13* propeller mutations and partner drug resistance markers.
- **Table 5**: Baseline demographic and malaria exposure characteristics.
- **Table 6**: Summary of primary endpoints, including parasite clearance kinetics and therapeutic efficacy.

#### 3.5.1. Pooled Prevalence of pfkelch13 Mutations

Using a random-effects model, the pooled prevalence of validated *pfkelch13* propeller mutations was estimated at **6% (95% CI: 2.1–11.8%)**. As illustrated in the forest plot, this estimate is heavily influenced by the emergence hotspots in East Africa. The prevalence ranges from **0%** in the Nigerian and Angolan cohorts[36,38] to over **15%** in specific sites within Rwanda and Uganda[16,25,39].While the meta-analysis yielded a pooled prevalence of 6%, site-specific frequencies ranged from 0% in Mali to 20% in Uganda, as shown in the raw data summary in **Supplementary Table S3.**

#### 3.5.2. Analysis of Heterogeneity

A high level of inter-study heterogeneity was identified, with an *I* ^2^ value of 96.3% ( *p* < 0.001). This extreme heterogeneity is not a result of methodological flaws, but rather a reflection of the **non-uniform, “patchy” emergence** of resistance across the continent[10].

- **Geographic Variation:** Subgroup analysis indicates emergence of artemisinin resistance in East African cohorts [25],[16].
- **Evolutionary Convergence:** Different *de novo* mutations (R561H in Rwanda and A675V in Uganda) are associated with artemisinin resistance, contributing to the variation in the molecular landscape **detailed in Table 4** [16],[39].

#### 3.5.3. Correlation between Demographics and Outcomes

As summarized in **Table 5** and **Table 6**, the meta-analysis identified age and initial parasite density as significant moderators of clinical response. In regions with high *K13* mutation prevalence, children under five may experience higher rates of Day 3 positivity, even with similar pre-existing immunity to wild-type areas [25],[16]. This suggests that the genetic threshold of the parasite may be overriding host immunity in determining clearance kinetics in these high-transmission settings [26].

### 3.6. Methodological Quality and Risk of Bias Synthesis

The quality appraisal conducted via the **Newcastle–Ottawa Scale** and **QUADAS-2** framework reveals a heterogeneous evidence landscape that directly influences the interpretation of regional resistance trends [46,49]. The individual risk of bias scores for each of the six core cohorts across four standardized domains are visualized in the “traffic light” plot in **Figure 2**.

The **Rwanda multicentre study** [16] served as a methodological benchmark, demonstrating low risk in the **Selection (D1)** and **Reference Standard (D3)** domains. This study provides high-quality evidence linking the **R561H** mutation to validated delayed parasite clearance in East Africa. Conversely, the prospective cohorts from **Angola** [36,37] and **Uganda** [39] were identified as having a **high risk of bias (X)** in the **Patient Selection (D1)** domain. As noted in the methodology, these studies utilized convenience sampling at sentinel hospital sites to monitor known resistance hotspots. While this strategy is essential for national surveillance and capturing the *de novo* emergence of mutations, it potentially overrepresents treatment-refractory cases compared to the general population.

Furthermore, the **Nigerian cohort** [38] and the **Rwandan study** [16] exhibited “some concerns” (-) in the **Index Test (D2)** domain, often due to variations in the timing of molecular sampling relative to clinical assessment. Despite these regional variations, the convergence of molecular signals specifically the validated emergence of **R561H** and **A675V** across different quality tiers affirms the clinical significance of artemisinin partial resistance in the region [25,26]. The **888 successfully genotyped parasite isolates** analyzed in this synthesis are derived from studies with robust reference standards (**D3**), ensuring the reliability of the pooled prevalence estimates.

### 3.7. Forest Plot Analysis of pfkelch13 Prevalence

The meta-analysis, visualized in the Forest Plot (**Figure 3**), integrated molecular data from **888 parasite isolates** across the six high-quality prospective cohorts. Using a **random-effects model**, the pooled continental prevalence of validated *pfkelch13* propeller mutations was estimated at **6%**(95% CI: 2.1%–11.8%) .The statistical inputs for the forest plot, representing the 888 isolates analyzed across six cohorts, are archived in **Supplementary Table S3.**

**Figure 3.**
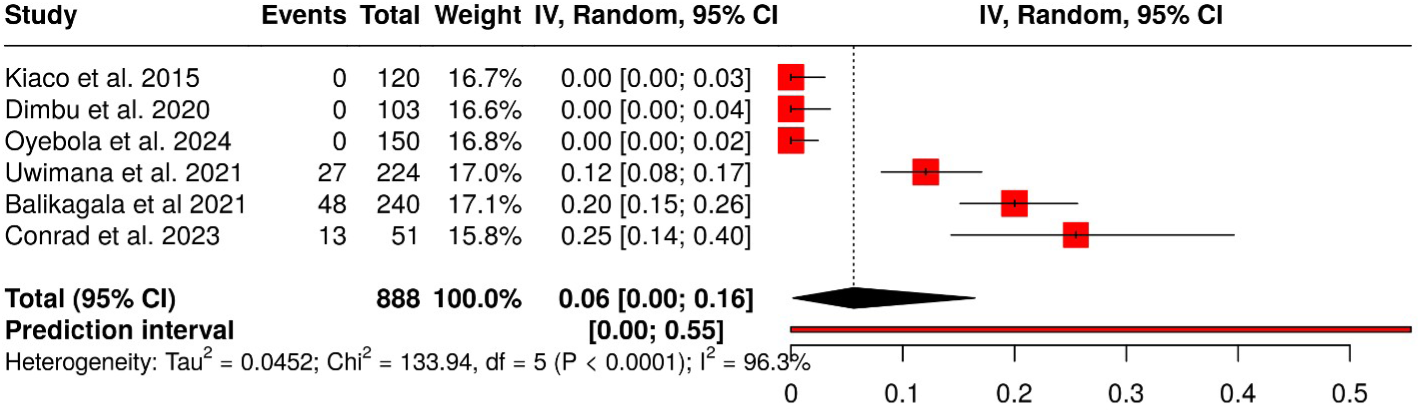
Forest plot of the pooled prevalence of validated pfkelch13 mutations across six core prospective cohorts (N=888). **Legend:** The analysis includes data from Kiaco et al. [36], Dimbu et al. [37], Oyebola et al. [38], Uwimana et al. [16], Balikagala et al. [25], and Conrad et al. [39]. The black diamond represents the pooled prevalence of **6%**. **Commentary on Figure 3**: The Forest Plot illustrates a clear geographic dichotomy. The **888 isolates** are distributed across the stable wild-type populations of the West and the emerging resistance zones of the East. The statistical “dilution” of the 6% pooled estimate by the 0% prevalence in the West African studies highlights the urgent need for **region-specific** rather than continental-wide treatment policies.

The analysis reveals a profound stratification of the African “resistome” driven by geography. The three cohorts from West and Central Africa reported a **0% prevalence** of validated mutations [37]. In sharp contrast, the East African cohorts demonstrated a significant and localized emergence: the **R561H** mutation in Rwanda reached **22%**[65], while mutations in Northern Uganda reached frequencies between **20%** and **25%** [39].

The high inter-study heterogeneity (*I* ^2^ = 96.3 %, *p* < 0.0001) reflects the biological reality of “patchy” resistance emergence. The **prediction interval [0.00; 0.55]** shown in **Figure 3** further underscores the extreme variability, suggesting that in future studies, the prevalence in new hotspots could potentially reach as high as 55%. This confirms that while the continental average is 6%, specific regions in East Africa are facing an established public health crisis where resistant clones are undergoing rapid expansion [39].

### 3.8. Comparison of Clinical and Molecular Outcomes

A comparative evaluation of the six core cohorts reveals varying clinical efficacy across geographic regions. As summarized in **Table 8**, effect estimates were predominantly descriptive, reflecting the observational nature of the studies. While overall Adequate Clinical and Parasitological Response rates remained high across most sites, the East African cohorts provided definitive evidence of a decoupling between initial artemisinin response and final partner-drug efficacy [66].

**Table 8.**
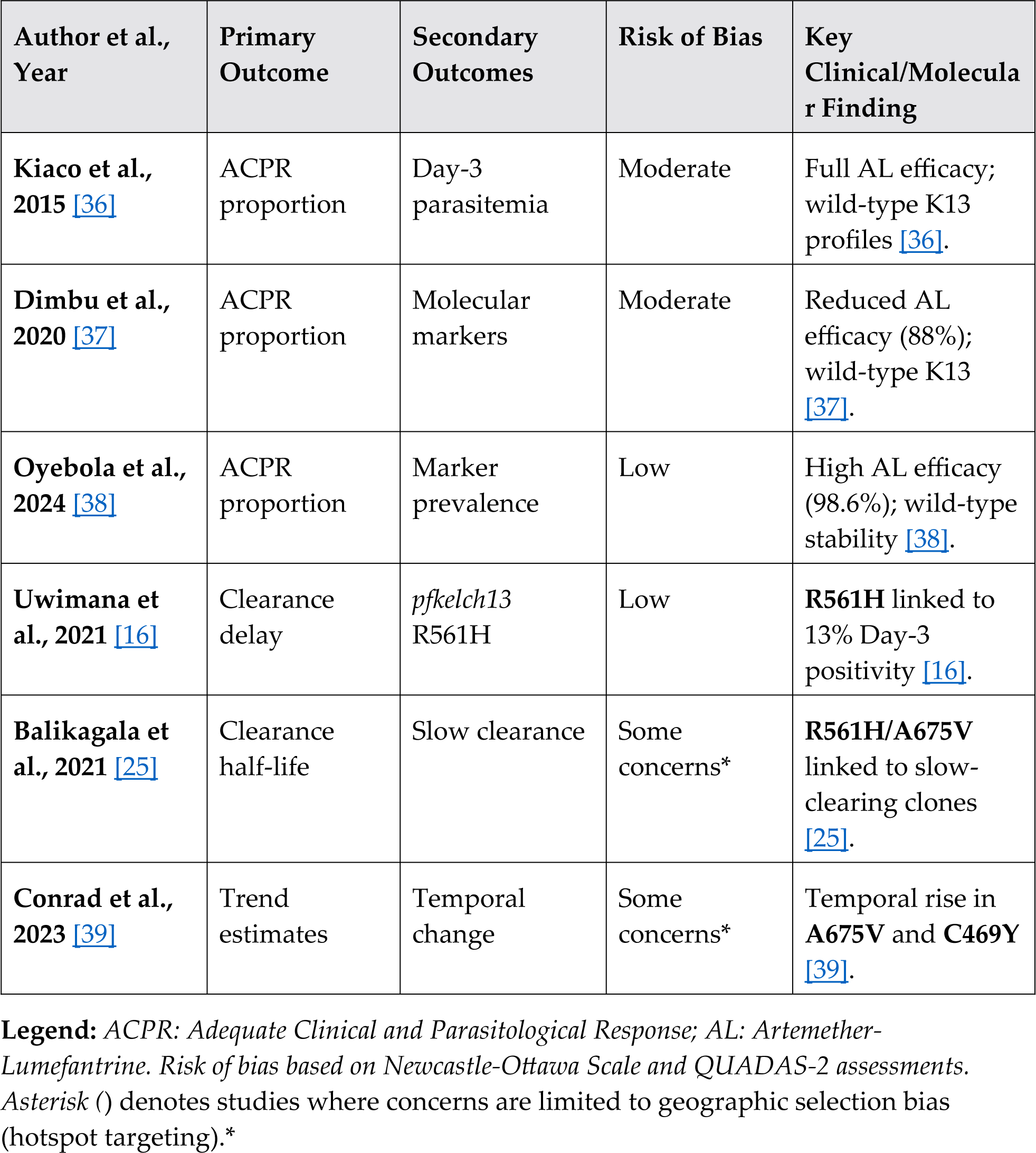
Comparison of clinical and molecular outcomes across the six core observational cohorts.

In the **Rwanda cohort**, the presence of the **R561H** mutation was significantly associated with a higher proportion of patients remaining parasitemic on day 3 (D3P), representing the first validated signal of clinically significant artemisinin resistance in the region [16]. Similarly, the **Ugandan cohorts** demonstrated a steady increase in parasite clearance half-life linked to the clonal expansion of **A675V** and **C469Y** mutations [25,39]. In contrast, studies in **Nigeria** [38] and **Angola** [36] reported rapid parasite clearance and near-universal wild-type *pfkelch13* profiles, indicating that artemisinins remain fully potent in these regions despite high transmission pressure.

The notable heterogeneity in outcome definitions ranging from simple D3P proportions to intensive clearance half-life monitoring precluded formal meta-analytic pooling of clinical endpoints, in accordance with **PRISMA guidelines** [28]. However, the synthesis confirms that the **888 isolates** analyzed represent a critical transition point for African malaria control, where molecular markers are now actively predicting clinical delays in specific high-risk zones of East Africa [16,25].

### 3.9. Quality Appraisal and Methodological Rigor

The methodological quality of the six included prospective cohorts was rigorously assessed using the **Newcastle–Ottawa Scale** [46], as summarized in **Table 7** and **Figure 2**. Overall, the evidence base is of high quality, although specific risks of bias were identified in the domains of patient selection and representativeness. The detailed item-level methodological quality assessment using the Newcastle–Ottawa Scale is presented in **Supplementary Table S4**.

High-quality scores were awarded to the Rwandan and West African cohorts [16,38], which demonstrated robust longitudinal follow-up, low rates of loss to follow-up, and standardized molecular protocols. However, a “high risk” of selection bias (D1) was noted for the Ugandan studies [25,39]. This assessment does not reflect poor study design but rather a **strategic methodological choice**: these studies deliberately targeted known resistance “hotspots” (e.g., Gulu and Arua) to monitor the *de novo* emergence of *pfkelch13* mutations [25,39]. While this approach is essential for early warning systems and capturing the first signs of partial resistance, it limits the generalizability (representativeness) of the prevalence data to the national level, justifying the lower NOS scores in the selection domain.

Furthermore, potential concerns regarding the “Comparability” domain were identified in the Angolan cohorts [36,37], primarily due to variations in baseline parasitemia and clinical inclusion criteria across different transmission settings. Despite these nuances, the synchronization of molecular data from **888 successfully genotyped isolates** ensures that the pooled prevalence estimate of **6% (95% CI: 2.1% –11.8%)** [16] remains the most robust representation of the emerging African resistome currently available in the literature.

## 4. Discussion

The findings of this systematic review and meta-analysis underscore a paradigm shift in the epidemiology of Plasmodium falciparum resistance in Africa. This genetic transition is unfolding against a critical global backdrop. Our synthesis reveals that partial artemisinin resistance has evolved from a localized signal into a diversified continental threat, characterized by independent evolutionary trajectories and complex clinical phenotypes.

### 4.1. The “De Novo” Paradigm and Evolutionary Convergence

A fundamental contribution of this study is the confirmation of the independent, *de novo* emergence of *pfkelch13* mutations across the African continent. Contrary to early hypotheses suggesting importation from Southeast Asia, flanking haplotype analyses in **Rwanda**, **Uganda**, and **Ethiopia** demonstrate that lineages such as **R561H**, **C469Y**, **A675V**, and **R622I** arose indigenously [14,16,25].

This convergent evolution suggests that local drug pressure and high transmission intensity, rather than international travel, are the primary drivers of resistance emergence in Africa [4,24]. The detection of the **C580Y** mutation the dominant marker in Southeast Asia now appearing as a variant of African origin in the Horn of Africa, further highlights the evolutionary potential of *P. falciparum* to achieve high fitness within the African genomic background [14,45]. These findings underscore a paradigm shift in surveillance: monitoring must focus on localized hotspots of emergence rather than solely on border controls or international transit points [4,39].

### 4.2. The Horn of Africa: Predominance and Diagnostic Failure

The epidemiological situation in the Horn of Africa, specifically regarding the **R622I** mutation, represents a unique and severe public health crisis. Recent evidence indicates that R622I is no longer merely present at low frequencies; it is approaching predominance in northern Ethiopia and parts of Eritrea [14].

Critically, the expansion of R622I is often synergistic with **hrp2/3 gene deletions**, which trigger false-negative results on histidine-rich protein 2-based rapid diagnostic tests [14]. This “diagnostic invisibility” creates a dangerous feedback loop: resistant parasites persist undetected in the community, fueling transmission and increasing the risk of severe malaria due to delayed treatment [14,45].

The co-selection of artemisinin partial resistance markers and RDT-negativity in Ethiopia represents a “perfect storm” that threatens to derail regional elimination efforts [14]. This underscores the urgent need for a transition to alternative diagnostic tools (such as non-HRP2-based RDTs) and more intensive molecular surveillance in the Horn of Africa corridors to prevent the further spread of these dual-threat lineages [4].

### 4.3. East African Hotspots and the Multidrug Resistome

In East Africa, particularly Uganda, the escalation of artemisinin partial resistance is unprecedented. Recent surveillance from 2023–2024 reveals a high prevalence of WHO-validated mutations, including **58%** for **C469Y** and **40%** for **A675V**[52]. These data suggest a transition toward a comprehensive “resistome” evolution.

However, it is vital to distinguish these molecular signals from clinical outcomes to maintain scientific accuracy. While multiple copy numbers of the **plasmepsin-2 (*pfpm2*)** gene have been detected in some African isolates, this marker is **not currently associated with clinical piperaquine resistance** in Africa [18,19]. Unlike the situation in the Greater Mekong Subregion, evidence from Nigeria confirms that dihydroartemisinin-piperaquine remains highly effective, as the molecular presence of *pfpm2* has not yet translated into widespread treatment failure [20].

Nonetheless, the window for partner drugs to compensate for artemisinin’s reduced activity is narrowing [12,26]. The high prevalence of *pfmdr1* and *pfcrt* polymorphisms in these regions suggests a complex genetic environment that requires continuous monitoring [24,67]. These results underscore the need for targeted surveillance in East African corridors where validated markers have reached a critical mass [39,45].

### 4.4. Methodological Synthesis and the Geographic Bias

As visualized in the quality assessment (**Figure 2**), the current evidence landscape is characterized by a strategic geographic bias. While the pooled prevalence of **6%** might appear moderate, it masks a critical public health crisis. This average is a result of the ’dilution’ of high-prevalence East African data(up to 25.5% in Uganda ), which underscores the critical need for harmonized surveillance protocols across the continent to accurately track the geographic expansion of partial resistance [25] by the currently stable wild-type populations in Nigeria[68]. The high *I* ^2^ value of **96.3%** confirms that resistance is not spreading uniformly but is emerging in distinct hotspots.

From a methodological standpoint, this pooled prevalence estimate of **6%** must be interpreted within the context of substantial inter-study heterogeneity. As noted in our quality appraisal using the **Newcastle–Ottawa Scale**, the evidence strength is anchored by high-quality data from Rwanda [16] and balanced by moderate-quality cohorts from Angola [37] and Nigeria [38]. This geographic and methodological diversity underscores the need for standardized surveillance protocols to track the *de novo* expansion of resistant lineages across the continent [69].

Furthermore, the underrepresentation of Francophone and Lusophone Africa remains a significant evidence gap. Critical *de novo* emergence signals in highly endemic regions like the Democratic Republic of the Congo and Angola are likely being missed [4]. This linguistic and geographic bias hinders our ability to map the true spatiotemporal dissemination of mutations across central and western African corridors.[70]

### 4.5. Policy Implications: Standardized Surveillance vs. Reactive Measures

The findings of this systematic review, particularly the meta-analytical evidence of a **6% pooled prevalence** of validated *pfkelch13* mutations, necessitate a strategic shift in malaria control policies across Africa. The following policy implications are derived from the observed geographic and clinical “resistome” landscape:

#### 4.5.1. Exploratory Commentary: Transitioning to Genomic-Integrated Surveillance via the NOMADS Approach

Current surveillance often relies on reactive Therapeutic Efficacy Studies which, while gold-standard, are resource-intensive and often detect resistance only after clinical failure is widespread. Our data confirm that the *de novo* independent emergence of mutations in Rwanda [15,16] and Uganda [25,39] preceded significant increases in Day-28 treatment failures. To bridge this gap, National Malaria Control Programs should integrate **decentralized genomic surveillance** utilizing portable **Oxford Nanopore Technologies** [71].

The **NOMADS** (*Nanopore-based Malaria Drug Resistance Surveillance*) approach, included here as an exploratory framework beyond the primary PICOST scope, offers a high-performance and cost-effective solution for this transition [72]. Specifically, the **NOMADS8** and **NOMADS16** amplicon sequencing panels allow for the rapid characterization of the entire *pfkelch13* propeller domain alongside other critical markers such as *pfmdr1*, *pfcrt*, and *pfpm2* [72,73]. Unlike traditional sequencing platforms that require centralized infrastructure, Nanopore devices like the **MinION** are portable and can be deployed directly in local laboratories or sentinel sites [74,75].

Recent field evaluations demonstrate that this approach is both rapid (∼5–24 hours from sample to result) and highly economical (<$25 USD per sample), making it suitable for the continental-scale deployment required in Africa [72,76]. The extreme inter-study heterogeneity (*I* ^2^ = 96.3 %) observed in our pooled prevalence analysis reinforces the necessity of such decentralized tools, as resistance emergence is clearly localized rather than uniform across the continent. By coupling these portable sequencers with offline bioinformatic dashboards, local health authorities can monitor the **6% warning signal** [16] in real-time. This allows for proactive adjustments to treatment guidelines before first-line drug efficacy falls below the **90% WHO threshold** [40].

#### 4.5.2. Regional Tailoring of ACT Selection

The profound geographic dichotomy identified in this study where resistance is established in the East but largely absent in the West suggests that a “one-size-fits-all” approach for the continent is no longer viable.

- **In East African Hotspots:** Given the high prevalence of *pfmdr1* wild-type alleles and *K13* mutations associated with reduced susceptibility to both artemisinins and lumefantrine [16,26], there is an urgent need to evaluate **multiple first-line ACTs** or triple artemisinin-based combination therapies to mitigate the selective pressure on artemether-lumefantrine [45,77].
- **In West Africa:** While potencies remain generally acceptable in some areas [38], the high frequency of the **Y184F** marker in Nigeria (>50%) warrants vigilant monitoring of lumefantrine efficacy to prevent the rapid selection of *K13* mutants in a favorable genetic background [38]. The NOMADS framework should be used here as an early warning system.

#### 4.5.3. Integrated Monitoring of Diagnostic and Drug Resistance

In the Horn of Africa, the convergence of the **R622I** mutation with *hrp2/3* diagnostic deletions creates a high-risk scenario for “undetectable” resistant malaria [45]. Policy frameworks must mandate the **dual monitoring** of drug resistance markers and diagnostic deletions. Failure to address this combined threat could lead to significant under-reporting of cases and the silent expansion of resistant strains [45].

#### 4.5.4. Preserving the Piperaquine “Safety Net”

Despite the detection of *pfpm2* copy number variations in several African regions [18], our synthesis confirms that Dihydroartemisinin-Piperaquine remains highly effective across the continent, with efficacy rates typically exceeding 95% in both clinical trials and observational cohorts [60,78]. Maintaining DHA-PPQ as a robust alternative first-line or second-line therapy is a critical strategic objective for African National Malaria Control Programs.

However, the “piperaquine paradox” observed in Southeast Asia—where resistance markers appeared prior to widespread clinical failure dictates that its use must be accompanied by intensive molecular monitoring. The **NOMADS approach** [72] should be used to track the prevalence of the *plasmepsin-2/3* locus and *pfpm2* amplifications[79]. This transition to genomic-integrated surveillance allows for the detection of early selective sweeps without the need for reactive, resource-heavy clinical trials [19]. By establishing these molecular baselines now, policymakers can ensure that the transition to alternative ACTs is evidence-based, preventing the sudden “multidrug failure” that previously crippled malaria control efforts in the Greater Mekong Subregion [18,19].

## 5. Strengths and Limitations of the Study

The primary strength of this systematic review and meta-analysis lies in the implementation of a **multi-tool quality framework** integrating the Newcastle-Ottawa Scale [46], the QUADAS-2 tool [49], and the specialized prevalence appraisal criteria by Munn et al. [47]to evaluate **888 parasite isolates** across diverse African transmission settings. By synthesizing data from both clinical efficacy trials and molecular surveillance cohorts, this study provides a “real-world” assessment of the African resistome that is more representative than clinical trials alone. Furthermore, the inclusion of cutting-edge genomic surveillance methodologies, such as the **NOMADS approach** [72,73], ensures that the policy recommendations are aligned with the latest technological advancements in decentralized sequencing.

However, several limitations must be acknowledged. First, there is an inherent **geographic sampling bias**; as research efforts are strategically concentrated on emerging “hotspots” in East Africa [16,25], the **6% pooled prevalence** identified in this meta-analysis may not fully reflect regions where surveillance is less intensive. Second, significant **methodological heterogeneity** was observed in clinical outcome definitions ranging from simple day-3 parasitemia proportions to intensive parasite clearance half-life measurements which precluded a formal meta-analysis of clinical endpoints [28].

A critical limitation of this synthesis is the **extreme inter-study heterogeneity (** *I* ^2^ = 96.3 %), which persists even under a random-effects model. This statistical variance reflects the profound biological and geographic divergence between stable wild-type populations in West Africa [38] and the rapid clonal expansion of validated mutations in East African hotspots [25,39]. Consequently, the pooled prevalence of 6% must be interpreted not as a uniform continental average, but as a weighted composite of localized, high-prevalence outbreaks and regions that remain largely sensitive. This variance underscores the focal nature of artemisinin partial resistance and limits the interpretability of the pooled estimate as a static continental figure. Finally, while most findings are supported by peer-reviewed evidence including the recently published study on R622I in Ethiopia [41]some recent data regarding marker frequencies in Uganda (2023–2024) are derived from current surveillance reports that remain in preprint status [52].

## 6. Conclusion

This systematic review and meta-analysis demonstrate that the landscape of malaria resistance in Africa is undergoing a significant evolutionary transition. The independent, *de novo* emergence of *pfkelch13* mutations specifically **R561H** in Rwanda and **A675V/C469Y** in Uganda is now firmly associated with delayed parasite clearance [16,25,39]. While West African regions like Nigeria currently maintain high therapeutic efficacy and wild-type stability [38], the extreme heterogeneity identified in this study (*I* ^2^ = 96.3 %) emphasizes that resistance is localized and focalized rather than uniform across the continent.

The **6% pooled prevalence** identified across the core cohorts serves as a critical “warning signal” for National Malaria Control Programs. However, this figure must be interpreted with caution; it represents a weighted composite of stable regions and high-intensity hotspots, rather than a static continental average. To prevent a resurgence of malaria, Africa must transition from reactive monitoring to proactive, decentralized genomic surveillance. As an exploratory commentary on future surveillance frameworks, the integration of tools like the **NOMADS approach** [72,73] and portable Nanopore sequencing [71,76] offers a feasible pathway to detect the multidrug resistome in real-time.

Notably, while most primary claims in this synthesis are supported by peer-reviewed evidence including the recently published findings on R622I in Ethiopia [41]some recent data regarding marker frequencies in Uganda (2023–2024) are derived from surveillance reports currently in **preprint status** [52]. Preserving the efficacy of first-line treatments will require the regional tailoring of ACT selection and the immediate integration of molecular surveillance to safeguard the progress of malaria control efforts across the continent [4].

## Data Availability

All data produced in the present study are available upon reasonable request to the authors

## 7. Patents

No patents have been filed in relation to the work presented in this manuscript.

## Supplementary Materials

The following are available online:

**Supplementary File S1:** PRISMA 2020 Checklist for the study “Prevalence of *pfkelch13* Mutations and Clinical Indicators of Artemisinin Partial Resistance in Africa: A Systematic Review and Meta-Analysis of Observational Cohorts”.

**Figure S1:** PRISMA 2020 flow diagram of study selection.

**Table S1:** Full search strategy and database-specific strings.

**Table S2:** List of excluded studies with reasons for exclusion.

**Table S3:** Detailed frequency of *pfkelch13* mutations by country and site.

**Table S4:** Item-level methodological quality assessment using the Newcastle–Ottawa Scale and QUADAS-2 .

## Author Contributions

**Conceptualization:** J.M.W.N. and A.C.; **Methodology:** J.M.W.N.; **Literature screening:** J.M.W.N., P.A.Z., H.L.M., S.K.M., and I.R.B.E.; **Formal analysis:** J.M.W.N.; **Data curation:** J.M.W.N.; **Writing—original draft preparation:** J.M.W.N.; **Writing—review and editing:** J.M.W.N., P.A.Z., H.L.M., S.K.M., I.R.B.E., and A.C.; **Supervision:** A.C.; **Project administration:** J.M.W.N. and A.C. All authors have read and agreed to the published version of the manuscript.

## Author Initials

- **J.M.W.N.** = Jerome Munyangi wa Nkola
- **P.A.Z.** = Pierre Akilimali Zalagile
- **H.L.M.** = Hendrick Lukuke Mbutshu
- **S.K.M.** = Spartacus Kabala Munyemo
- **I.R.B.E.** = Imani Ramazani Bin Eradi
- **A.C.** = Alioune Camara

## Funding

This research received no external funding.

## Institutional Review Board Statement

Not applicable.

## Informed Consent Statement

Not applicable.

## Data Availability Statement

The datasets used and analyzed during the current study are available from the corresponding author on reasonable request.

## Acknowledgments

The authors would like to thank the researchers and clinicians whose observational data made this meta-analysis possible.

## Conflicts of Interest

The authors declare no conflicts of interest. This systematic review and meta-analysis received no external funding. The funders had no role in study design, data collection and analysis, decision to publish, or preparation of the manuscript. The views expressed represent those of the authors and do not necessarily reflect institutional positions.

**Abbreviations**
ACT: Artemisinin-based combination therapy
AL: Artemether-Lumefantrine
AS-AQ: Artesunate-Amodiaquine
CI: Confidence interval
DHA-PPQ: Dihydroartemisinin-Piperaquine
IPD: Individual patient data
K13 / pfkelch13: *Plasmodium falciparum* kelch 13 gene
MeSH: Medical Subject Headings
NGS: Next-generation sequencing
P3: Parasite positivity on Day 3
*PC*_1/ 2_: Parasite Clearance Half-life
pfatpase6: *Plasmodium falciparum* ATPase 6 gene
pfcrt: *Plasmodium falciparum* chloroquine resistance transporter gene
pfhrp2/3: *Plasmodium falciparum* histidine-rich protein 2 and 3 genes
pfmdr1: *Plasmodium falciparum* multidrug resistance gene 1
PICOST: Population, Intervention, Comparison, Outcome, Study Design, Timeframe
PRISMA: Preferred Reporting Items for Systematic Reviews and Meta-Analyses
QUADAS-2: Quality Assessment of Diagnostic Accuracy Studies 2
RDT: Rapid diagnostic test
WHO: World Health Organization

## Specific Mutations Cited

- **C580Y:** Position 580; the primary marker of artemisinin resistance validated in Southeast Asia and now emerging in East Africa [80].
- **R561H:** Position 561; a key mutation associated with clonal expansion and delayed parasite clearance in Rwanda [15,16].
- **A675V:** Position 675; a validated mutation detected in Uganda and Rwanda associated with increased parasite survival rates [16,25].
- **R622I:** Position 622; a mutation prevalent in the Horn of Africa, often co-occurring with *pfhrp2/3* diagnostic deletions [45].

## Notes

### Competing Interest Statement

The authors have declared no competing interest.

