## Supplementary material for "Prevalence of PfKelch13 Mutations and Clinical Indicators of Artemisinin Partial Resistance in Africa: A Systematic Review and Meta-Analysis of Observational Cohorts": Supplemantary materials

### Supplementary Table S1: Full search strategy and database-specific records

**Panel A: Search Strings and Filters**

| Database | Search Query | Filters / Limits |
| --- | --- | --- |
| **PubMed / MEDLINE** | ("Plasmodium falciparum"[MeSH]) AND ("pfkelch13" OR "K13" OR "artemisinin resistance") AND ("Africa"[MeSH] OR "Sub-Saharan Africa") | 2010–2025; Humans; English & French |
| **Google Scholar** | pfkelch13 mutation Africa clinical resistance "P3" clearance | First 200 results sorted by relevance |
| **African Journals Online** | malaria drug resistance pfkelch13 | No language filter |
| **WHO Malaria Reports** | Therapeutic Efficacy Studies Africa 2020-2024 | Surveillance reports and gray literature |

**Panel B: Records Identified per Source (Total n = 874)**

| Source / Database | Records Identified (n) | Search Query Logic / Panel A Reference |
| --- | --- | --- |
| **PubMed** | 179 | As per Panel A logic. |
| **EMBASE** | 282 | pfkelch13 AND 'artemisinin resistance' AND 'africa'/exp |
| **SCOPUS** | 218 | TITLE-ABS-KEY(pfkelch13 AND resistance AND africa) |
| **CINAHL** | 114 | (pfkelch13 OR K13) AND (malaria) |
| **AJOL** | 40 | Regional African research focus. |
| **Web of Science** | 33 | TS=(pfkelch13 AND resistance AND Africa) |
| **Manual Searches** | 8 | Bibliographic cross-referencing and snowballing. |
| **TOTAL IDENTIFIED** | **874** | **Initial records before deduplication.** |

**Legend:**
**Commentary:** Following PRISMA 2020 guidelines [[1,2]](https://dummy-citation.com/citation?d=z%3ArVhLbxzHEaYdRA6QW06xESSNIHAkgLvch7RLCjAcipQlUlxwLVLmuXe6Z6d3e6Yn3TMklyfFueWUU5L%2FkYt1yUX%2FJD8guSSHwJd81dOzu3wMkYcBy%2BDMVFdXffX6ar9%2Bt%2FFhbKyaquxDcWHs%2FN3GjxI%2BcYXlUbF48IeThStkygsVMSvPlbxwjGeCpbLgLZ5xvXASb6xk0jmZFYprVhjmyjTlVl3h9bkSMoskTmtoyab0Wcaxini08Kocj2WxYCZmieS6SFhE6lRWSHtOGk2GC6KotLyQujoCXYpP9KLNThPJIo2roIG%2BwOzM5VCQQTtUFol0dHdubOE2WWIu5Lm0m0w5lpmCmbxQKddtNjbGBjGyESfdbceFSlWmHFSSXmXZOdelJIcijQ%2BRwt2bLDda4fKUz6XFI1llIG5Z6fCifaIIDbxgAqZok6dwMpjKvnxz%2FPp4xB5%2B%2BYZrcuk4Zq%2BXRuFhtIJduUfspAAmdL7V4mvWT0tgDosky8uJJoMF8GTdnZ2dVotMkSzh55JNpMyYI0AQtcjArrwoud6k6CZGGG2mCBOeyYecEoIeGRfnHLION065FT6mFAaTiTIqQoDuR%2FJWCrXZrnZmcymAYyvb71Dg7Y9NSSFPeMHmcgEXkcgkZjLGJ6YsQvRdUQoFcxFzExdwOUewkUmVkVK0gTHgvgqO4IRyrpQ%2BdlUeZl4pXJ9aU%2BbVjQijLgWsk5e5tIpyXDBeAji79K8G0RVIniraEBq%2FPjgZ7bKHYytjaS3erGJ8AEcdHLOsofB8BuwG1B4xTm9RZEaX3u%2BQR1VBw%2BCQUKuMINX%2FSUBIk6T89q77x8bybFMVBq%2BWKUn5gCSF5zjKWW%2FYUvjAokRGc8S1yhNOMbStPOFAPdbmAjXGp5anobKXwgFtBNEDJCTuAPTLnkNurWq%2FuJ6A%2FLbHbXaQASmkxPPLXMPpKmtg0nON3EGvoWdhopJ82WQX6GQkiDIifFPJM1Lucz0kR4Wt5BEgWtmNa9vsi%2FoDPXplwR8fvEue5jgNQ6fGrNcOtEPY12tunFMTLalCkDSUbWhChjohogSHZZor68uzTvcbOUif0FNQ60VpZYXvzZhtVpCs3PYtFYLcOYP2hmJhZ3LCHPSwh0lR5E%2B3ti4uLtq5VS7lLVcrahs73XrEXGJKLdBnkDs6j0sNex0CHoxXaW4N6vi%2F7xYaA4GCL%2FZx4bTX6Q1anScpMq5AhKQ9QCd4t%2FH9aSjHtxo3oXt9gX%2FGikF72Je%2BxD%2BShSq0TMdH5gSVJdDCMxlTMaXygSZ1Vk1KnHG%2Fw5gUGU8ljcuYp0ovkiM1ITSVnCpUwmxXwyKYag3%2B03Lj9ol4Vxcpz4L8vimnGvX7ot0kn5zK4krzOK5OJIcyyxSi36h%2FVGqLCVfp31tkiCZn%2B4361Yv33xRXDtlaHZmOJfKD7TXKHxgOC4Ry4YpDk2RsjN7deEW8h%2BE8D%2FrFSM1lo%2FJ9GkSSl5dBeNw%2BbEbmlZZqlsmAZXRoXJOoHBmqIC8n9znoSCUZo%2BeMRfyPSAn3buNBJIy6%2FEm30%2B72h92tGZIUudbGcBbtbqfTwT%2B4Pcne4v8u%2B1p1nzzeaXV7w2H4azAcRCmfzvqdXm%2Bn0x%2Fs7CTo9dQVLudnq5ciT5VIujuDXrczJI3H%2FJ8zn4o%2Bk2e9Tmen1Rm2ep0ETWla8qmcyCytR2FI7A%2B8i5cfj%2Bl1xJCHlltPd04iP4jmdZm9sfpyQnXqUKjBKdfOtXG%2BQukPeOjTfoujBCMtt2Kl5edKfHYfFp8Wi1x%2BhqpHwaErTV05oTdAfR7UnOLxA%2FRFaeJXKhPrr1HJWi8gqyP0EfFssYc5Xnys%2F1Ise5s4AxN1%2Fv2Pv62K9PKPd04Z6rmr8bk2NF83DU2PU%2BiRpzTJn1djTrKX1YDbowF3sD7gnt43JeLC5Cpyv0USxcKkaEBwbaIE7%2FkwmRPqnLoOjZOxklpUIpPutpf59b6MlCPdtRQ%2BxqfdTq8zqGL9i2vMr%2BLNtxqldJGx8tvP%2F%2F4v%2B6uNjY0EUVm7THS3O48rbb8k%2FDCeYPYmG1sz4RPl%2BSbpfYP7LfXRAjG64VJ13gSg7nSpN%2FQySd1OK0%2B6QxSQv%2FvnL2VqxAJ%2Fw%2FCRyRTaaz1KEWHL80Xtx9%2FGL9kdfvSGjytUpielnUr7P5nZr1TYIAMQYrRvCnZt8OMA1qc31CCznOQW09wnAjivT4Ol0T89fnuH0f1BDcAnL9CIiHHfvlpQEf0Aq46%2B3KyrFn3J1%2Bp95Tinxe0rbBdQ8sFqofsoLHQ%2FXC1030t9Ff3fzBPs1VciZvyKPFzbNqiVgb1gwIG2qWmGtxj5xANuZy4sX2dvS24O8rPw%2FKMSq7QBamJFKFzPTALjFgpcicysvyyq9aDNjjFWvGjOQciEjLiAguUSA1Nv2bPGm8JaKS1Wv%2BrZrx6ZJP6hyHew2IyVuaAOAg%2FppiXdvkazQFM6K7jIRdCYapf0aK19ozuXlDeDQdeXO7K82kIQSC0jkOx1fyrrK4qF5btQ8WITex4JwvEcmxwmeIWWI5KA%2BKzWpMpkpAt2ORDFsPsh5dHq11eMioiv9kgUtULC%2ByjHPKJ%2BQoiAeYFg1YdXtDvMA0%2BGg3qvdh2rWzuD38jAw2ETLlqxbA%2BFQH4qTVtpDZWVkUlxN0Ljd5glP69o%2BM3r6iK5fl9FgCkvQDev7SiVwuWqRaJVGhCH93VzJ0%2FtXyOWf%2FrkNmURY8z9QLFGvMD9F%2BweLjSKXsnsSoUT6pBImmTPGw9Mn2GZKBcwyN8w5jAmmrPRfQfKwqLuKhp64PhEat3I5JKXJo7Bc4O8POVpuviuOGtykgB5WTO66RF%2B%2BwHt%2Fq4odLQ7B53xmp9rJe9httNnaFVZzeajE1C4BkmxhzUoYPHaYGA183FwjHMFaIP0YQkbGl17YVXqEh6wiw%2BBSTPO6Uv7%2Fs%2BTGcoOq1wdyV03mb3%2FxtLOcrflR%2FQDVyU74pmZ3ZMkk6OwCCWn%2BB1shvprZORHRtTxU0D5CulUJI2Gj%2FjC%2FPXt789Q27XhApSt0WjUwz7t4yKAeFJgsWgSno2iF6VCqqJIQ0KV83vDDnUXQDGAfiSdls3JfZqAlgTN8hDfGteU%2BBRFGJmAyi68k5ztNZohz6RGm6qM%2BEqdA%2FJGi88SRe0wWDFG1jYa3LQsrS1JhohItz%2FYmqSzdjbsrq1Fv1Hd4ZNBq7vd71fLULe7Peg%2B2e4P1peh5ctqGer3h9u9zpPh9TWo10WjbPV2rm0yj2tORD850PVo71vUTDE8yJyt%2FrBXm9XGrw0gSSJe7SYbYZEYN87kp6tRLm78ZraaKrd5d9gA3q7Rt5%2FdRd%2FWUPs3), this multi-panel approach ensured that both high-impact peer-reviewed data and regional surveillance reports from Africa were captured, minimizing publication bias. After deduplication and removal of ineligible records (**n=182**), **692 records** underwent screening. Ultimately, **189 reports** were assessed for eligibility, leading to the inclusion of the six core cohorts analyzed in the meta-analysis.

### Supplementary Table S2: Detailed reasons for exclusion at the eligibility stage (n = 183)

| Reason for Exclusion | Number of Reports (n) | Detailed Criteria |
| --- | --- | --- |
| **Lacked longitudinal phenotypic or genetic data** | **175** | Reports missing paired Day 3 clearance data and pfkelch13 genotyping, including duplicate cohorts and conference abstracts. |
| **Inadequate sample size** | **5** | Studies with fewer than 10 successfully genotyped isolates per site. |
| **Non-English Language** | **3** | Reports published in languages other than English or French (after initial eligibility screening). |
| **TOTAL EXCLUDED** | **183** | **Reports assessed in full-text but excluded.** |

**Legend:** Breakdown of the 183 reports excluded during the eligibility phase according to the PRISMA 2020 protocol [[1,2]](https://dummy-citation.com/citation?d=z%3ArVhLbxzHEaYdRA6QW06xESSNIHAkgLvch7RLCjAcipQlUlxwLVLmuXe6Z6d3e6Yn3TMklyfFueWUU5L%2FkYt1yUX%2FJD8guSSHwJd81dOzu3wMkYcBy%2BDMVFdXffX6ar9%2Bt%2FFhbKyaquxDcWHs%2FN3GjxI%2BcYXlUbF48IeThStkygsVMSvPlbxwjGeCpbLgLZ5xvXASb6xk0jmZFYprVhjmyjTlVl3h9bkSMoskTmtoyab0Wcaxini08Kocj2WxYCZmieS6SFhE6lRWSHtOGk2GC6KotLyQujoCXYpP9KLNThPJIo2roIG%2BwOzM5VCQQTtUFol0dHdubOE2WWIu5Lm0m0w5lpmCmbxQKddtNjbGBjGyESfdbceFSlWmHFSSXmXZOdelJIcijQ%2BRwt2bLDda4fKUz6XFI1llIG5Z6fCifaIIDbxgAqZok6dwMpjKvnxz%2FPp4xB5%2B%2BYZrcuk4Zq%2BXRuFhtIJduUfspAAmdL7V4mvWT0tgDosky8uJJoMF8GTdnZ2dVotMkSzh55JNpMyYI0AQtcjArrwoud6k6CZGGG2mCBOeyYecEoIeGRfnHLION065FT6mFAaTiTIqQoDuR%2FJWCrXZrnZmcymAYyvb71Dg7Y9NSSFPeMHmcgEXkcgkZjLGJ6YsQvRdUQoFcxFzExdwOUewkUmVkVK0gTHgvgqO4IRyrpQ%2BdlUeZl4pXJ9aU%2BbVjQijLgWsk5e5tIpyXDBeAji79K8G0RVIniraEBq%2FPjgZ7bKHYytjaS3erGJ8AEcdHLOsofB8BuwG1B4xTm9RZEaX3u%2BQR1VBw%2BCQUKuMINX%2FSUBIk6T89q77x8bybFMVBq%2BWKUn5gCSF5zjKWW%2FYUvjAokRGc8S1yhNOMbStPOFAPdbmAjXGp5anobKXwgFtBNEDJCTuAPTLnkNurWq%2FuJ6A%2FLbHbXaQASmkxPPLXMPpKmtg0nON3EGvoWdhopJ82WQX6GQkiDIifFPJM1Lucz0kR4Wt5BEgWtmNa9vsi%2FoDPXplwR8fvEue5jgNQ6fGrNcOtEPY12tunFMTLalCkDSUbWhChjohogSHZZor68uzTvcbOUif0FNQ60VpZYXvzZhtVpCs3PYtFYLcOYP2hmJhZ3LCHPSwh0lR5E%2B3ti4uLtq5VS7lLVcrahs73XrEXGJKLdBnkDs6j0sNex0CHoxXaW4N6vi%2F7xYaA4GCL%2FZx4bTX6Q1anScpMq5AhKQ9QCd4t%2FH9aSjHtxo3oXt9gX%2FGikF72Je%2BxD%2BShSq0TMdH5gSVJdDCMxlTMaXygSZ1Vk1KnHG%2Fw5gUGU8ljcuYp0ovkiM1ITSVnCpUwmxXwyKYag3%2B03Lj9ol4Vxcpz4L8vimnGvX7ot0kn5zK4krzOK5OJIcyyxSi36h%2FVGqLCVfp31tkiCZn%2B4361Yv33xRXDtlaHZmOJfKD7TXKHxgOC4Ry4YpDk2RsjN7deEW8h%2BE8D%2FrFSM1lo%2FJ9GkSSl5dBeNw%2BbEbmlZZqlsmAZXRoXJOoHBmqIC8n9znoSCUZo%2BeMRfyPSAn3buNBJIy6%2FEm30%2B72h92tGZIUudbGcBbtbqfTwT%2B4Pcne4v8u%2B1p1nzzeaXV7w2H4azAcRCmfzvqdXm%2Bn0x%2Fs7CTo9dQVLudnq5ciT5VIujuDXrczJI3H%2FJ8zn4o%2Bk2e9Tmen1Rm2ep0ETWla8qmcyCytR2FI7A%2B8i5cfj%2Bl1xJCHlltPd04iP4jmdZm9sfpyQnXqUKjBKdfOtXG%2BQukPeOjTfoujBCMtt2Kl5edKfHYfFp8Wi1x%2BhqpHwaErTV05oTdAfR7UnOLxA%2FRFaeJXKhPrr1HJWi8gqyP0EfFssYc5Xnys%2F1Ise5s4AxN1%2Fv2Pv62K9PKPd04Z6rmr8bk2NF83DU2PU%2BiRpzTJn1djTrKX1YDbowF3sD7gnt43JeLC5Cpyv0USxcKkaEBwbaIE7%2FkwmRPqnLoOjZOxklpUIpPutpf59b6MlCPdtRQ%2BxqfdTq8zqGL9i2vMr%2BLNtxqldJGx8tvP%2F%2F4v%2B6uNjY0EUVm7THS3O48rbb8k%2FDCeYPYmG1sz4RPl%2BSbpfYP7LfXRAjG64VJ13gSg7nSpN%2FQySd1OK0%2B6QxSQv%2FvnL2VqxAJ%2Fw%2FCRyRTaaz1KEWHL80Xtx9%2FGL9kdfvSGjytUpielnUr7P5nZr1TYIAMQYrRvCnZt8OMA1qc31CCznOQW09wnAjivT4Ol0T89fnuH0f1BDcAnL9CIiHHfvlpQEf0Aq46%2B3KyrFn3J1%2Bp95Tinxe0rbBdQ8sFqofsoLHQ%2FXC1030t9Ff3fzBPs1VciZvyKPFzbNqiVgb1gwIG2qWmGtxj5xANuZy4sX2dvS24O8rPw%2FKMSq7QBamJFKFzPTALjFgpcicysvyyq9aDNjjFWvGjOQciEjLiAguUSA1Nv2bPGm8JaKS1Wv%2BrZrx6ZJP6hyHew2IyVuaAOAg%2FppiXdvkazQFM6K7jIRdCYapf0aK19ozuXlDeDQdeXO7K82kIQSC0jkOx1fyrrK4qF5btQ8WITex4JwvEcmxwmeIWWI5KA%2BKzWpMpkpAt2ORDFsPsh5dHq11eMioiv9kgUtULC%2ByjHPKJ%2BQoiAeYFg1YdXtDvMA0%2BGg3qvdh2rWzuD38jAw2ETLlqxbA%2BFQH4qTVtpDZWVkUlxN0Ljd5glP69o%2BM3r6iK5fl9FgCkvQDev7SiVwuWqRaJVGhCH93VzJ0%2FtXyOWf%2FrkNmURY8z9QLFGvMD9F%2BweLjSKXsnsSoUT6pBImmTPGw9Mn2GZKBcwyN8w5jAmmrPRfQfKwqLuKhp64PhEat3I5JKXJo7Bc4O8POVpuviuOGtykgB5WTO66RF%2B%2BwHt%2Fq4odLQ7B53xmp9rJe9httNnaFVZzeajE1C4BkmxhzUoYPHaYGA183FwjHMFaIP0YQkbGl17YVXqEh6wiw%2BBSTPO6Uv7%2Fs%2BTGcoOq1wdyV03mb3%2FxtLOcrflR%2FQDVyU74pmZ3ZMkk6OwCCWn%2BB1shvprZORHRtTxU0D5CulUJI2Gj%2FjC%2FPXt789Q27XhApSt0WjUwz7t4yKAeFJgsWgSno2iF6VCqqJIQ0KV83vDDnUXQDGAfiSdls3JfZqAlgTN8hDfGteU%2BBRFGJmAyi68k5ztNZohz6RGm6qM%2BEqdA%2FJGi88SRe0wWDFG1jYa3LQsrS1JhohItz%2FYmqSzdjbsrq1Fv1Hd4ZNBq7vd71fLULe7Peg%2B2e4P1peh5ctqGer3h9u9zpPh9TWo10WjbPV2rm0yj2tORD850PVo71vUTDE8yJyt%2FrBXm9XGrw0gSSJe7SYbYZEYN87kp6tRLm78ZraaKrd5d9gA3q7Rt5%2FdRd%2FWUPs3).

**Commentary:** To ensure the highest level of evidence, exclusions were primarily driven by the lack of longitudinal monitoring or paired clinical phenotype data (n=175). The exclusion of studies with small sample sizes (n=5) and those published in languages other than English (n=3) is a recognized methodological step to maintain the robustness and interpretability of the pooled prevalence [[3]](https://dummy-citation.com/citation?d=z%3AtVfLbiNFFHUQINawiVigWjCQkWKnbcfOYxZo4piMZzJJNHYmghXlruruSqqreqqqk5jVABv4CuAzsuYHYInYID6AFayQEKe62857hodYJOp03%2Be5556qfHZWeyXSRsRCvcJOtDk6q72Z0LF1hoZu8tpPuxsP%2B73R4Gl%2FuE5GmlBrubXEJZyINIMJ0RExHOYidELFxE6s4yl1IsTrY8FPrLcItTrmygmtqCTaECodNwpWx5yknIkQr53h1KWwgochTNBYaevjOIRHSk36KpbCJnVJVZzTmJMsH0v4%2Bri2QYaj%2Fc2PyGZ%2FONjaIfd3NsmwPxoNdrbWyQG%2FUBeVckIspyZMOCOP%2B5vbg50%2BWdg9FuzuYtHZMBRchZz0hCtik4Fi%2FJT0TzOKB0YWDvjYd1XZwQvvSS83BtWTnlau6AIPigOhq%2BYkz3w39zMjJGktL5JW0AoapC8xhLH0gLhEM0usy5ngtsIcef8R6i%2FDi6AvHkW%2BQAQScPap0AdmFcrclpiOTnQVkRtLBNrPOH4p5zEMDeeqqksKTJS63PB7iIw6MXCEjanDMDNuwDEMujBl1FHCTwuGIQvQwziQlV12wnjCI7jAEzWlmeSA1ZPPF0nDMIf%2FpFHOViGuFZ9WCSLUCVQsUdSgJpBMwvBJf7i%2FPQKN%2ByJO3DWUF5rBJXzueosS8ikeJDS%2BS0HvEeV7pIxhCH4yM4RnQ%2FEIC3WVxrO6p2MCwtMMnlmzkWNPJr4dJZAHo2wur%2Fh6aL34Aj54FJaAzPXJlyEzozNtqklHvt76i%2Blg8NajbXRKWneKnEFwp0E2qG8QzUflsl5CbfESC885MN2vYpWv4%2Fdycko%2FSsgNWODr8lBav4weSWiFFbESER79muJbq73U6qxdQWhh7c7dBnkM9D1%2FUq3kpNzvWyOdUGSG%2FSIZ544kFPjr2capwvfCdnhcb4K%2Ft7vT294fDnZ31smT%2F7ikNMuAZGGYUOAPiB0k4nJNL1vjWwqVIGNB3004xRChbj3opPBzFKwzAxXps9qrMc0xcmOfyzLph%2FjRhrUbrS53AsWckoc6h5hD2CPSk0IVuPYzwXgqtNTxJDrWMk%2F5%2FFfZLPi2RofanNXmRIamn%2Fgh4yTCJIx1e3gz%2F0MiafX4o%2FR%2BRmAoKORL2DFFU28fRTQVcpJs6jGIGOY8BklVfF9hMTllRkteu27%2BbCf%2F7usxRUmmPsSxA1KWfhvUjKm51S95JMFeCEtpzgYmvdU2OsDic1NaRls47bS71XYP4kpZFbWfviDqEESeRR1YLaEat9jGD7g5Oi%2FhARqTEKybjQ%2B3qMEsk0slz%2ByjhNo9Fv0WCmbPaq%2BHTIvTd5pBoxk0u0uHjcMQU%2BeZaIBDzUaw3AiaLSbsWD3Hb6s%2BF8HqWqe%2B3O50RXN1ZbXeWWs1w5TGh%2B1mZ7Xd6QSdINE4VyjIdHRw%2FpJlqWBJu73WbndW1hBrl%2F5%2BiIh5SdlDn64eLNfbAbhSbs%2BYq7RYIJtMGTxXtHrUl9bz3pCNp7HNx26S%2BfaP0LYIJR%2FhzznsCtfRI5weF19DU3BjgK0MQRS2MenpXLn5P0GcCGhBN9gB7k22fD1fLcW3FzcfiXFmQmCqk8rrYgTdghhW16K%2FIQUCe02OBT2%2FIvjlKkUMC%2BpCSJY%2FK1MveLNLlc5EaNf9EXtVAKLy2xcYaMR0irVEj2PBaKsATA91KBChurVYHgkuWWkybq4WNs82eSgKZZ9a4WM0auI%2B0y0MTt99fC7JvgEI03UlwuZqw%2F%2F44Nfv3%2FqmVqslGM%2BFZKy5GiyX0d4fTpUbJ8%2Be0WM6FlDESRF3H%2FmNVxeHYV1pqfTXDzgunsnNLbXLik1lg%2BgRxNEjX%2FXUba1VPU1NNIYzIQN%2FNfH31vKuWAWf9qR2jz%2B5oad2NwjKaG9v4VaDVSPXE%2F8fbTRXlisw37sSxp9VxaldoNlneUm8aSMfd3%2F5%2Bd82wvy2vYGbjzxtJM5ldn1pCRrS0CZeerGMHPl%2FSZ5CHhFm7i8%3D). This rigorous selection process resulted in the **six core prospective cohorts** (**N=888 isolates**) included in the final synthesis [[4–9]](https://dummy-citation.com/citation?d=z%3AzVxLjCTJWZ4ZeWEkJCQMCK%2B11oZlBrrtrprKqup67PpBP%2BfV1d3b3bPDrEByVGZkVXRnZtTmo3tqODDGNjIXHhdAnICbkRaEEIc9IAEDnDgguCEf0J7BXLCEhIT4%2FojIrKzuyp5Zz4gdyZ7uzoyM%2BP%2BI%2F%2FH9j9hf%2F%2FDKNV%2FFciSja96Zik8%2BvPKTYz5M0pi76fRHg%2FW1jXu3Dvbu726%2BxTbjbMRikcgk5ZErmIzYfsCTUHkyC5nPA1dOeIxfxzxhE5UIj%2FGIKUzG3UCwVDHh%2B8JN5Sn%2BiAVPQxGlGOIxd8yDQEQjkbCQR1P8E%2FBYcuaqKI1VwCaxGsU8DPEei4rIE6F0GcccSZ3didhaNFIBX2FZlMqANRuN1grmDFSs3s9kJNjSxjvL7AxUZZqohPkyTtJaQO%2FSsYj5ZMqwC%2FjeVeEkkC5PMc5SgRVS%2FXEsJgF38UJGMpWgeMqGU8aJf27WAS8rRCEo6Kzol3EKUhN8ENWGnBbHAkMZ8VSqqFh6aW3jCPTJdGw%2BEPSiFmSh8Dk2QHOwtrPCNpR%2BvfS3f7q8XGdbvg9C3SlL0sybMuUzTAO%2BVeRlLtEPQszGsKFIz4TQdLX1huOX1RWWjNUZ7Qcj0vRm4IDMnJjto9%2F4Tr9%2Fo84GW0e39zYP32JOo1WczAQc4PQSdiZifBbhlAKaynVV7MloRKf94PYenZwrBD3BSe3PfeTjE71%2BNjG8uyABqweaQkgST2SqAjXSz8QpDzK9bTg9HFWzxzw%2BTVbygYJ5IsJP7Dm%2BlvgjlcSL3mg6PTsB1sO5QHxilbhqMl2hU2ATP0wmTUPXSEQqnU7MwEgk%2BKS2v3GAgYp5kH3wkslkPFs4Fm6ceSJxBWmFH6sQn51h%2B7Wwg%2BC3zRoqmIYqnoxlEiaMpzgpj7jpdTTJTrPdoW0HKV7sEBViRb%2FodvrmOU8nHf3c0MmTROB%2FmkzQVzvY3tnXHyTi%2FQyk0CnQTk38E6dVw0FMBM4oLvirs4Otw%2Fs7RzjZI5DnZpgzxgbp3eo79RbOnl6oYcohGx6pfhbg9KzYpGPwoLklwcgFYgWM1Ds32FLEvsKc1jKkUwYBTEJxpBwawZlDx8e4n4Ii2p2ZQaDlfUhCMhZene1FemBjhfaABvbbpaNMONRVYNEzGXg14oqRJcEjs0e5HBht6LbqbUtZp79sN0fv9m6v85BMFvQa4kKbqCKot8JemyWYy%2BOYpiESwgwGEJZLr8SWHs5NRef4NnOwkCE4yacoNk5GLlku%2FGY%2FITlkURYORYwvGw1LY9NZ1vt2njeaGEdqRAFs4akmi%2BwAkZjzAY3bvHO4cf%2Fw8M7erj5k0HLuEGF5tdnFNIU8Q1R8iWWSDCI9SeVQBqRXEP%2B1HRpINhJGICELpvTKhf3weMqNTMDCNOl1YXKMvculj4QTa5OXEZgqla7VksIIqCzFQuDZiMhMPMhUgBBPegxTMYw%2FgQkDRZgkg3UqBNwMnX3pcxlAyN8mlyTi0xL3ntJTgfFTSAwYJzNI4gxRIxEitcYJ89Jk0GYJO2lmgK3PUiIfxjgbYUb6A9oBDwYn5WfkKkIFrwFPC8ZlAgMhPCOUKeyBdmV1OBdXa%2FQmZHvUbDQ7tcZqSD6Q9C%2B%2BE%2FnqwyuvjXiWjlWcPAkkjsFNt%2FF%2F2NxW3WkJmSSZuCpSmQZCDaypvquyOOKBf6rIo%2Fz4pJhwR8E8qvjDK1fht0figDgBHIC6wDnu48nrV%2F97DAef%2Fx5odyzBKpb%2FNkZ6EQ%2FpC9%2FnoQym4p7krhIj%2BPdodA9OLhpxD45BXLk4dnwk5CMhY26Gi7uKR5WDRwOOffbs1GKAo66e%2BEAlT%2F8olnZw9K6MR3%2F35%2F%2F4jUDCequqr8QOrGNiPvE3ZaRiu4APLLPv%2BT9wpZd8eOVHXE%2FJR687jbrj9Do3E6fZb%2BGQnNWa03B6tRbcwzB6gn%2BT6BvSaXdXa81ed9UN%2Bei42cSwdr%2Fb6o2xFM5DPDp5MHvoTULpjZudTrfRaTcxwx7%2FwbE%2BTi0NxzSw5jSxFk4EDggnMhRROMmGARlKKxxXNVvRulQDCNcG5DTmwQlUHiLgivtx8Gh3nKaT5K2bN60jPzbCUR9K6JoHD0Zf1KF4N10FNCXimxPPv1nJ8CjJhqRq2MkTrYCBOMKfV2F9hPIhA175sUgJNGEskCKs9%2Fp0g9b4zJegVT6sNdTMewAImpjHK0aOH%2F3lVuH6yfJVIaQCumidKjsUfLQQ2BmkAFsIT6pcaaEC2Qxjl4EtyOfCthkPfW%2FOjc67cyy6k2EQQKjBXH6qJjBq34TQ%2BJ4KoXDgeyg93tZnpG4LHmClQzIz0HoBHxV4Zsiw2dVjxjhEmKFI4Jl%2FBL%2FQd%2FTzR2%2Fkqn2AD3nsAjaCvA0DloFDVCz%2B52vfTw%2BfXLlyZYwTKs3tNbutvpmlvU%2FC4zJDyQrbik5lrCLaMovB9lw3I7%2BuICF22AV2DEnhjvTFYmacllluBVLpjgkNpzFQ1y0oMll9vdAAZBvbjUFAfFPDcdtprZqP39ygiAMWGIMhqnoechIQbb0BOdPpp1%2F76wVMOy2nqSd6%2F8JKL%2B982s2upfaLt8EkTD1RqMfg59xRHeWyCQRnCB%2F9zV%2F94cLTatsz%2F%2FkD7hmKV9huBoUCC%2Fn6es47sDJwLh6p43WAueDRcq7rMFp1FY%2Bq1fiEIr93RZzgqK9ejAh%2FYhYRXru2jg2JEP3Y6EVkOMWZ8s17Ofi%2FOINmQZwo7CMUQX4UQD3TQ0hPyyEHyIYZSOEtTaD2XNFTEebU2cYYUAkz2FjKzYBm5lV%2FccBaQHWaLCHIGImAERjSmk1krsOnZYKCzPe4jC0038mg8ewwC9g%2BQQcSFIM7tfHBcs%2BK6ZYZwQq8TzJwJWrlYHJp7XANYSutY3cVE5ZCH0JXJga4PHKCjkwABgXg4Ez6IXUIW4rgaCJizAwPgI2ng0kEHKGNkkBXAAAI2MvjE8jIC%2BOUtsEp16x9v7GG%2FdZLDUlS1gDfyCaTRYOaK3vMFr58Zm0OhfzuAle%2BKcNhZlx5sC%2FgxtkB9zl0qsr331YxFjGw5UABhF%2BCLjae%2FnEE5GnRxftrEYcQPP0g4WwgXWx55YfbAhte4B21wTEQkozPSKwqkc8GhEEggtZfyW2g7CTEZ1Xj%2FVuwMdIu4u9kj%2BEoKseuRV789Ltm7BjYDcDCq8Zg%2BwLxSWApUbTO0w842%2BWPOSapZPsu5kRCwu7XeFOB%2BpFKKs9ifTY4XBdBiB0DgE2qVxgRF9yzdEVQ02M8j9iuFGnlafg7fJh5dpvGh1CYMRvUq8GhePoXkRk8ucWHiAOh8IqiAmD4qiXuEUyY5vA2C2Tl1grkRbCz5og3x3F2KoHtKic%2BPJWjYvghB5iZssNK4uURwFPkPZauJWUrQDamemMy1x1LuzG7HMfFHlTO7Q%2BAk6WFzZdDeH8bSpYj7PG2DAA3o8rzAeAHaoTNMlqsBYBtx8AAlZ8c7wfwNH%2F%2FreTEUn88wK5myePiYC2U%2F68SlE%2B1R2z2bnLu1htOv92vNRslEP9NiURep9buNdrSaXQbtU6z18Jv%2FV6ts9rvaGDfavS7PacHFF8C9rOHBti3Wk6n12m0FwH7ZqPmOLVG%2B9nA%2FtHn10JEyy6yqocArMK6U1gesp0aHLwUQH5nISD%2FKWuvNwhoyggMsB11NktDUgJy5ul2yp5ulonUyVGn%2F4mj4%2F%2F8319ajLdeLjr%2B4dlxOh1DyBcGKgEgSFVtqGJKsUDhgIWMh7QJ8oKr7DfZq8wVhL2AyIJWSQESEaED7SXsUKevDF9bE6RdQiPSBXP3%2FmHxkTXNwR%2FrKfUnJRD8swtB8DmVfwb%2BLVVErstyRYTSo8MsRoaIpL%2FIjdNECTAMgclsWDvklOiCCviku5o%2FDXyhy8A68CJ5VGugbrnEQpUUSUeMlL6Bv0gkPrLZuLw6AgQN46rhXQG%2BzyPri1C6lBeDDclpx3nkkJ3gpQc0DXypc1RFkYbSwvjblB2KLPRsvgvrVmFf2glKYlIE4cFmY9vAGb6qQOkJLIiF5EAe8IKgGHlLNQGkJYkt1Sr2tZXDh4QlJxQmks0anSs0JNNwkiqE79UpAkrijY2g6%2ByINmY7HBgGcZghAdlnwvwJsrpgxmD0%2FXqJctRq8gKJSVNaui6GCshrzuP9dByrbDTG5MiGZzQDfnFPaqiZnOI4Kd8%2BB%2F4DDitBWb0pE48ICF0smVAdxmZ6dRipBW3xUczXGXJadOYDoBzZZMiEFlgK7xBKLKEOQccKSAIR1johYvUYjpaSqrGP6hklWVMB386WUHJxDLuovaCgRdqEo0K2J9cK71zFkRLvhh8KWuYKIOX9xrgglec%2F1ulyB8sitbNczkMXhagL82RDSaYXOpNMhEv1JEM%2BsY6ZsuHEzATuiprLubIKVWZItHS4NacmeYIbHEO%2B8KrfpvIEsOz7VNx4vqCOaof7es89tkp1F9KQC6Nt5pst7exvL%2BsiD6RgTSfWQV8pGFxBHpzy%2FuCWiMVwZLCxCm23J1LzlNaiVVmne7PTZUtUsTDlVaRpJbjRee28KgHIgcjfy3igLVQsFKQL4TMbS5KlufO%2BWMXRtqmo0Cz1W%2FXujWUTfOvHm1RuwYtet75qiciz%2B%2FU8x1QS6EI0i00HAfBtlKxOkeAz6%2BITAyrN8iSmS%2F1GvYkFaOHtjWY3f9NkSx06NqrHoooRGuNB4kemxJqzWXphabB3x1Dp1LtUtaICBTaJ9oKqBng9q07YzAOSNKtsCkXF%2Fo4gxlDCSR2iCaqs1JH6Wr5KBS8qXkWQkSn%2BQaYyS9iWs9rsbc5lLo2Bg%2FijamMmQznagSCBoY293Y2d%2B0XxyBp8%2BGVMCm%2Bo628ZlEsXB8tGH1aMtkeLOinYuRTMvKrmRfbnOaxTDCVG8lMiUYEFtDUXWkoBvlAOiTIxVDQFq26gscysHFtnu9gbbC0EDF%2FTHhvZzjPMyPyMIl09hlDojZ7bMmTGAw%2BuGW0MfpaSXiHDobkrTK12vyUMo6fFwdQsu8EF02YPu%2BTM%2Fl%2FKQT%2FzscpBr5XKQa%2FNJWK%2BsyCI25uKITy2TUbcw6%2BTSyJyOeKBDd2C7SzCQz5VbKMy%2BBztnWFrpzbwH63BinPgwYrR3hpU3M6%2BF2RnfDTE2VTXp9y1IK9kPRQpAujKQH%2B8BUBzhrSZjW3XSaooEVtJd5D5WNvOjuBtfElyyj%2FEsZ1YUtQ6H3JNDBDIGbcfzZeoXtMlqs%2BeT%2FU22xCeRg9x6%2BIaVSl2bbcRkjqO023Z2LXfRIG%2F010Yu2LaXq3Zf7Gi1N5LKUrNWHwZQfD1xUWpN2wQ%2FMGaBh851F6YC78E%2FlY2G11El29RFh0DYzGmBhcYHdvvA4NRYIkCDpB%2FKgMmk7L9xMPt%2FxBf%2BrdXOjDtdXuGkOV9BGshgCqdJVZ6B8Al7yrSIoBMuSBICI9jePvWn%2B2BtVeXtxeqS%2F3Kv76zOKPwAnWpLz6rLjXT5GcE5p9%2BIR%2FZr7e6xkdeN1od4aBKNsq6yJ%2F%2BOC7yU1%2BfuchPjedc5O%2B%2FscAZ3D8DZsw9k1ijJppKx3EfuFImuT%2FdVUgwV%2Fqv43dFdEI4n9vEtRgImKiXlFT23hsrW2IZvTeWU1Xt7uQgi6Yg4rGlWx5Bv1zserVDvQ0%2FPtuUENUbeSLZgHvHkLzKto%2FMVSOVr4LaBhLeaeVoxCESttmm50c7AgWA7Up2USGIPIieZ4Z7B2CoauzJvhRwa1pZ8oy73rKqU72lAg%2FcWrrvoN5RTYjYeizCXFjW40vI%2BHhp%2F9E%2BIjeR9%2B4EAwSHGCueflCd0JdbCcwH%2FJkFYCcohUWoCCVsrRqy7Y7w89h%2B4W%2FyiHapugaA%2FqEcPgZ3BY9qOwqdl2y3uubCUb3LRYCQFbqsKqfPgFjQs2jRKce5VqLHkwHVqz3k8YYFbkOKBvFS5fTrqBLBf9taxAP0QEoesoPKvQkHGXQV%2BG7Eo7O8MrVBnvUSVfEHqHDnFkHsPP0uqmdVLNz3xnzKTzJAAqtYxkRQa1XAEUwv%2Fi4aDGWaTbMQcU44zblfg4IOEZFV0XUb0X5e%2FBoBILuXGJPLak6L2q8%2BR%2B4C%2BXG4i3a3VUOppb%2BEVslGA56uVq7ePDH1Geqw6nVanU65PlM8zOszvQ6qPM5CjEvFGcTgz1Gf%2Bdxu7sTvIETGvlG2EjDQOO05zHsrd4KT0K1H7lDWoyCsR3JcH6nTmxaeJjf3BxsO4HgTpSQNejEgTGqoLLW7DaeOJy8D675%2B9V8Wo903Ldr9E6DdokOqMj97IgJ3jL7Ug9WOc7vowkSujrIcHnonpiYBej73CMh7cEa1Z0BdTI9DqiGUEwECewCHAF0RcbhikiomBatj9ot4W%2BPiTxzqfv97v%2Fz1VxkOvgjUTX5h95Wumr0Q1D25%2FgeLcfwLQN1mruVnZ2cXtRyqP9P0XM2fAXl%2FbFaLuvpzlJWDhguoDOX9SiUlXQsAWkWNA5Ejmsipu8omlAHtyuWgPFxM2ZflV%2BeSc1%2B%2BKb%2BqS1BDujwy30tvilkmaZ%2BeKfrW6r%2F%2BapapD9G9PywiUR35zmXbTIv8jIjcQCBNt4Q8EHVJIy9PQfZdjrLJXOWbnqLzCk1CdjIrK6bhnIxEUl9emEhrzUHz31oAoNehDCfwxjkCEesiTSthnL%2BdnRQdJSM4sWl2UgknxJ0TkY91B9jQajyD9KJFZXIvodYkdnRZqwf86HCGnA%2FHMvroyW%2FfwVNUEKvA7UOg%2FaxwwP6Ao%2BOrcrS3N8lbq%2F0HUE8RV2NVHEve93MI4BEjj3VJL9L0LAcm401wexmOhFsDHMlx3hZiIux9FXv7OEEXUV%2BOLyQVDaujrHAP%2FUojVVuDzk3zbq2tEAgdbVrszghUVnOxh9ewlvlGUg%2FLZV1n0u65PFLJWF52SN6A7s0sHFyCRplGRqudm5E4DhVH2s7ptssJv19DP0uzV2t3%2B%2BhiWW21asj12S4W3NNotBut7lwXS%2FHQoqT26qrTbPYu4iMU0Ztz8GZzzvCBHB3og8YidUd07m7dHRR0fi3HKUoFyVfSGAFmgW2uWCDS2srvY%2BTdJtaIwbZb%2B5H76MJGWUTwpGSWbyzKQFzYuMuvRl4vNwLYezDlKj7l7IDBTP%2F583SyovfBFCNChUoQMhUo6hNDeWaQKgYYYWy8tsGHKCGiNo3Ra4ltZ49QyUGdlkrKsL1b%2Bp211LokbkqHNj2IAtQYQRLduUT7NnUs6DKfAWql2nB%2BSYv6MHG%2FETYdXk9nLScIi8hZ2LuQM6tvyxnaJeQlX2qgR4nYxw9Ul3bQ80lToDozAlJGdEpVEbo%2FBDvA%2BClKlhRi5PeG5l3bzKlVNE6UPFTRE2Cqd7qq9FhfZ6NaqYcbfnQvlDK5YdEeSzlb5cEFPSjVv%2FODwEvMnMxRDv7pZirKeCm7jxDOg6eiy4IYiG21N6KcdlFDJ7UpVYrXp%2FoJWlGaTXsjca4wWjhFA6bxBjCJdg9OEFUqRK5U9r2Y%2BNWleXQOgDVqNcBlJaxzg0h1nPxCndMpUXqGU9a1%2BlOBkFVPRdXK4lKpcfj6DFm703%2Bot7TTXX23XJvXU2jIEAFNxBBKarCNI7sztgmsA26drrmEUdyIKx0ufSK07K0YNuiIdC4cimMqt6UdchqkhKtUQKemZcgofBq9OLe4LbrmaygDE7DAyJQGiT0wtl3wU2whGnvmVmz16PO22U4qtua7qPt%2BSDXP6PLoObbtIZuFdKCUL4TrJpif6psQJrpOCnRi9NxER6A8v7lc7O4ly8xt2jzlzdYN0tYZHe22s%2FNshh1zoQ8fa%2B1Gt0tJcPBoIR1YIa%2FszorUEFhT9TAQda4Tyl6unEl8GeBqkcXN8ovWzzAyN1NJHcp9AeWSv1FbfZHciv7sivr5S%2Bh0FVMDYTBNz0wLpK5z8whd2agG2W76DME%2F2a5yCR1KDn9pL0faHSou0p4jXDcJzVlq3Yw1s3R6u22XATW20O141KZdvZ9aCa2YW8UiISidpSkSkcCX4gNU3RkaGED0Odh9aU6jAl4%2Fbw5eNNv1Vn8uCf%2FozV1cnt5C4E9k2mo1LZiHWjYt%2F%2FrVb3%2BcxPy1f54l5q99bw79%2F84CJIegnrrKbScxtd2jxX%2BzOiG%2BlmQ5Hn0X%2BoBccdVIdM5bhDg6JB2aVufwbwESDNGZM8a%2B6hBkkwP%2BVKLPXaS0Z%2F380J9QVSNucQj3a289oMh8Nq1G8nKXhxkkdlTkYpGuNu3YorKTf4TYBRdl8ljhaIwAv7rtf0eMCoR%2Bl%2B4XIRlVfScCt%2B0hyfZwDpCPc9FWWd2%2FL4%2By8EwM%2BeP8E%2FTbw2idsHvVEcZuooY5%2FKd6CIH%2FnerhGwqo2YYv8gCdCNQiVZ3Gv8fDfHKUIqj5tTox7G9CRIuLBLew65VlDX8doCkfenwXpizGjeRqOkAp9UUi%2F2sp38f9KTlhdxdmX3VnwfkQo4nCXaN873VhiFHuLkCmFb0FuJFnY4ou%2FkRQsTDz2tLdBa3nybwibE3gttBJnMIb5Skfmyz44UMTw99lockL9Nn%2F0%2BKs67I1hLtbZOmKnKt1CnmkY5xCKQbSTiGPfvIWT%2F3QuJtPPDP67w%2FvvdKF8hfJjP7eL370inTet8y9gdiOQXoMoR1Bk8TmSNsdeyP3C5u61c7e29X%2FuZTZ%2Bdlu%2FJy%2FX%2F18bdFt4lan0TCTfZZQHormlvLyuh8j8DbqNpcC%2FT8%3D).

### Supplementary Table S3: Detailed frequency of pfkelch13 mutations by country and study site

| ****Country**** | ****Study Site(s)**** | ****Dominant Mutation(s)**** | ****Frequency (%)**** | ****Primary Reference(s)**** |
| --- | --- | --- | --- | --- |
| **Rwanda** | Kigali / Huye | **R561H** | **12.8%** | Uwimana et al. [[1,2]](https://dummy-citation.com/citation?d=z%3ArVhNc9y2GZbTJu3k0mvcaWdwSUeeEVfkfm96WsmylVqr2ZHsqpMblgBJaEGAAUCt1pd68hPaS%2F%2BCM5MfoLMu%2FR39JX3AJbVSbHriiQ%2B2dskXwPN%2BPc%2BL%2FeFm57NEG5EK9Vu20mZ5s%2FOHjC6sMzR269%2F8d1p%2FJFPjeC6sUEIRw62wjqqYk13GJV1zRuYdklAZi4KaMiex5NRUBomWUq%2BESgnd7hAsqMWaWOcLoagTWhGXcUOL9ZM9IixZCcZtYThlhMZGW0vOdQkLagHECkoWpSNOE0YdJxm1RGlHFpx7aIXGOYwA5jQxIqYk%2Bt%2Bbf%2FdJhxxzw8mKk5TDel3ABjv%2BDPeLqEd2yTxZchln%2BPyEFEYXXEpuCNM5FWqP5KWrIFt%2FxioTcUZiqkjOmfBw7rl5P1CDvSGBbwobcgfPXM6VI5bmheQWkcARscedGJ1jXezfMpGtmdGbHZXfMyhEgTh9XwqFkxTC499xH7xAljlPqHLGv%2BNJAt%2FjNcFXKiuoZyssoGSEUFxkQnISlwiIAWYE3MfmL6n762TwtbddaJdhaQOTmtzubcJ1F5qzwTA6vgsGWSELyJpyIhGb8EcTohPSHYzI7qjT%2FxqhhCUMLKGOzKilS9oh82wtNTLCnYjhEJVrRAz%2BX3Eq6wzx64Iq68%2FAdgi0UEz4HJa2xiDhME15hzzHPgRpcL7cYq0SYZAVbIIDXYZ979D6hDEjrlrThW9XwhndIS%2F9QutKtva1cOUrk3D%2FZ1PdpsLIOErwShPkwgAb3gDrNlZBXRx1vt573CY9e6TQzkeRSrn2DYIzK%2Fu0OgdOwbkSG9kyjjkao4qJEzmV1GeaxBnPkbyqmbZN0JG0MufsKWCk3bA7CMJx7rdDTXPzrUr0zc7nKUWXaWPfSJEX6Ppn%2BKcN74Wd3oQLa0v%2BJRLlJNen1PnqmcGvGBskV9qX31d%2FLO62PNExxeKbnUforZSfUZVykA3Kw1g3x5PHXxxkEh29%2BXws%2FUoj0NkA8C9YMkVzvyJJaC7kOn21gpeK8hRpU3zqs47mkHznPbYnPAX9sI0tO0II2kyTc6eX3G4sLw%2BoWYC4yHGnzV6cMrEst0DM3zhVwYkuUSUz%2BIg8tKK6oAYNzzcLk5lPmGo1fpWXUtj6lORUg4Tadz5N8fey2fkpVYLLVg9mpVpTxV%2FXe4uXGTUxstsazmxGXyMq6N5qQV65fEALh%2FJtzwGOSctmTfKCIhGi9YRDECFNVePBnNqYtntwaISNSy11bT6V%2FFrYNnM%2BNYKv6ww%2FM7dv2e3bD1TEclaCQhmFDNbg05lW4nt0XRv6M%2FTnCnh%2FEfrkmWA6Xm5ss6cUXEKmrfWmZgvhynWZo93ydQNoilZYgCBbY3%2F7VlHT4HkqmOBmYxwLZm92fhczLa4fR2EnCnvjfduPBpMoCLthEIXhIOgyYRfqDf636gcRhaNxMJ4MhiIa9IdBNAr%2FEec0veyF%2FeE4nITDYQaNBHfz6%2BXF9iEvchyWjgb9qNefsCIXLOt1R%2F3RuDu6rMikIqNLkFEIMgrCXl6UC9Q8yGvDR48qt4qaa%2BYQdkwTQi9BnLo0MX9l5PUgc66w3%2Bzvr1arDmYJWHZAm%2FsgWoEpxL7rW6dgSWrLhZ8BELJlbfkSXx9BG7hOXiD99x9z59kYtpgTUPAH60NdKvf4sx%2BhkAmUEwzOLjA72er5V7sbjrz%2B59GdGnipjqVGjB7KWSMzD4ajRhccmYMfc80EJpN7Q8q7CgxTvKIW6O5JfeJ0IWL7hnlPfw%2B1l9dPmmgh%2FR1t0v3WCtjOhZ%2FXc%2BGX72jIEDl7QNv%2F%2BdOv5O2PIb3Lv3O19NMLuGyz94xD9tqJ%2Fgpjg6w575xCMdfkvLXv2HeZLutu%2By4Ta63ST0iox%2BCWbVDyM5qIpYB%2BsEvB2lmmjHWqm1OSqRIxd%2B2cjeyhqmregyLSjDxrdTc7R8Ug4Y1knsGhVoKcg01ijjmvYTx%2BVIWsLavPtWTwtsb9LfS1HQg%2Fes3zplgOzAdgiJdUloq9RhRqEJKhcdtAzClGo7hWDDnrkCls%2Be1PqrVixJGncTRXrUrLqaLq9icMuu10%2FVFKnNYDQ43p3iRx2lrFL%2BgCRd6MELh88E%2BjwB%2BveukBZDUF%2FW2E9UJIKWhOzlpjk89K9Cpd0JSqVe1Cdujn1g%2B0SjIrfYvXKT65favb6%2FIVy%2Bia%2BumsBpVvKAKZR5TzTyqxyTGuVc1UkfoJ8wNkkpxgWs%2FqnbNTipGeXNTWG0H%2BohLkP1d0HA33bdQf9YJeOJnsdqMnYRj1u0H4UJQbA%2F%2BpH%2FT7g9FGlKHRk%2FGw91CU7x7WMtwbD%2FvjMPqZDIP%2F%2B0E0eKCvzxvFgJp3VLwQHSXzjhJZJ9VXW5Wdzw4jiEd33J%2FsQ1%2F3YZDbIBr3%2B6Mweqi4O7U8%2Fji1Vse4FtVa%2BEvErrm7%2B58IcENtfnxoxO%2Fe7w53KviNvzP6UAQSrSP3iL9MSR7gTuvv8hIO4L5leHW%2Fxa2Jo2Pi7f25uvm9R0mD9yppW%2Br%2BDw%3D%3D) |
| **Uganda** | Gulu / Agago | **A675V**, **C580Y** | **25.5%** | Balikagala et al. [[3]](https://dummy-citation.com/citation?d=z%3AzVbNbltFFHZaQGxYsaESiJFQRYti5%2FoncVxRVfkrSdvEIT%2BNuhzfmXvvxHNnLvNjx7sUNkgsQLBixzuwyjsgxCMgdjxAV2z45l47aUrNigWLxPbMmXPO951zvpnnF7UbiTYiFeoGG2szvKi9m9GBdYbGbnLzl%2FW1jcefHfSP9zbvkYxawviIS11wRgy3wjqqYk6cxroRI%2BrEiFuiE0KN47mwQgm1SFzGSU6FIrHOC624csEkEca6uhQK5w2nLse6JcgFtpIaQRtkA7sipvKVWC85J7lWGv4NLSYEP8vvJJV6UJ5KhVZ2ERux9EyolKwlBg4XyVh7yciAk8LogeQ5Mo8bZHfraLu%2FeXgPBxIeOxwmY%2BEy%2FAQdI660t2Vw6xV1nNyhZIxPU7daeni5ltgVIXcJVYxw6wTCgLhAR0ENtQI%2BYsnxNQDLqExAR8Ib5IQTPqLSl%2Bb8jIzESBPrbcwLJwZCCjcJDF5z5BE2JZQYfNTBVcpxwuAkiKDW0kmZRQoQbnK9fHXDZRkJm9w2yMHW4fGTI7CQewcAYDAQiBJZ2FiRKpGAQ%2BXkZJEkRuek3ejdDuS3ouZyqE%2Bz11idLfQWCfOBZoA3Qk7Cfsj7ymNi%2BBeeq%2FgS0tpKd%2Flpme1GZ6X3jFApuURb3dn%2FNGpEUbPc2if3SfjVWQxIilAtNObkboMcggDJ68qDWu0EQ3AtJ7k2RSZsjg6TVA0DV1exZkBDzscp3KOw3AQCMQhAKpDChDCRJFhF95aoXaZtwEEOtYcrah1Zs2XX9vc2nhwf7vT3rnGYowRoONAw5KZqdMbdtM0q6PA3K4u7rCwqcuehVwxUDUAfkn5ECxrCxoKjEYKjsLqPpIC3cnaIvdBUgapyJmzjbkPSOOYWnG%2Bi3Gkraq3Uo%2BU81sphOLnZUYm%2BqL2ZUsDRxp5LkRcQgYf404a3Oo12jwtrPX%2BHO%2BEkP%2Ftwj4%2FJFtgOUR5pbxR6DcF3ORMxPCYjzEXOby08Ly6DPNExhbuL2oIo0KMHVKUcEoSWghrsY%2BXWG99kEmRW33%2BQ4aQRA49D9ltYMkXzcCJJaI5%2BOl2nUgxpCsnAtGNG%2BTp3bsKMlrz2T%2FPkoR96NjVN96ie%2BKGYZ8x3hnxmG%2B8KPtfpY%2Bq80ZWh6NsBN44cNeaZiyMaZ2JA1dR1fpgJ9cf5dztYNXreqewZzamHSXUo2aUWRM6zZv1CTBNKTqiESM0FuYWumZpmh3ToDR3OdZusqcmYBl0N1ptAa8naXKDJY5HD3TSNLWHhfS68fVQwRr9Nfe8J3ABKzeU87zOtUl1fw8U1mYbQW3lOleeS7KTIcj6KPrbRdzMijRNqLj3buBynnIsjbTPxb0Viu8JNk3nVOMHtuc%2BSF7Fg9qL2Vsy08M2o0YyWV5YUP801bTWjZrezwoQdqHP8t%2BpLEUWt1Xqn22uL5nK7Xe90opU4p%2BlpuxVB%2B6J2t5vhMsbc8bPhydUiK3LBsnZnebnZaq3CV5%2B%2BOC1nt5z9U8x%2Bsx716q0WZk2lHrM2QCMWuMWEhVpUUrBQojv7AK1m0bLeYrJsNd0Y9Kn%2BDKFYGP2YHxt5tpk5V9h7S0vj8bgRUDW0SZcAdalgydIM7t7Wo91LuA8C%2FZDqI62lve%2BM5ylkN9xRoHU428TPBUgb1%2BgpxV5e5i6IM2xlDLFk65MN7ZW7dfMnPCpKuY45O8G7xpbr73001a721ghXQ5BICNba1b1dP7gU4N3qGRIkvno2JE4XIrZfoXwJ0%2BFFg7ADwWinJEpvc0xaNtNeyxPBJatMBq1uaZPNtBFryVEzinrNiuT3Z9EQHy%2BCOKsuwCB%2B6B4bQ4H%2FevBnY%2Fv3Wq2WgaCXfLNWt92rvHT2QwVjUmWyCHEeoQFVeFqFZwA89uPYF%2BWNhIXK7L%2BD01rtrlaJ3N3PqMkxzR6FQqTPPWQal1UFCoXgJuECc5fOsP3482%2Fn%2F2dsnVZ3uUrkk200CFQh4JoNw7WqHV0%2BZ2fgvv%2F619eD60zL%2F%2FEBZUJLneJNtRdeLtRcXqOlzx1MPdhiYTDejj1m7fZs1jBe5Zi9Vk2G4U3%2FFPc%2FWFn4Gw%3D%3D); Conrad et al. [[4]](https://dummy-citation.com/citation?d=z%3AzVfNbhvJEZYMJPE9l2yQIB0EBlaARHFIipIMBAtZkmXrh2JEyc4ee2ZqZlqc6R5095CevcRJkAfIKc%2BQR9A5LxDknFPOue4pl3zdM5Qorbl72cMaMG3OVFfVV1XfV80%2F3q09S5QWqZDP4rnS07u1n2Y8NFbzyNY%2Fef7q4PDs5OryZnT0klnFbEaMa0uFMEIKySJVlEqStEwlyy%2B2Qm4odq9DIbkVSrqjmpeCzKb3UihjmcBpbTmOFzznWnAW6yqFBRWkUzhAiImqYM9hfWBgwGXMpJrDhyZuSRp27N8lWkS8w8ZIQvCcaTLCwHNEm2yeiShDBCkSMtYwblhMOa9dgjlx7awYTyzpNsl6kwnDCooFIsSs1EJGouR5XrOwZkVlPSLjsnNQppTDf6mVJTw5C%2Frs83GCfzY67FwUwrnIlUyFrWIUI2cxt8ChEXPGRc7DnFhTnxa2TwcwTQmI8ZPKLiFz8Re4L46v31weTV6yDX%2BUI079FQJrAEUCkoxhFRykKHxOUYVqA6DNVGw67D3iGQMTHFg0Ai%2Fh2TzKHPgRkkSaWXaTIg6XLEYyMERZE60K1usGA9cbVaUZvvSCDrs6ntycXyO1V7V%2FsoWPXjMEADjjuQcMlCVaYVAuw%2BbCZgxvBEqFnJRmEaL5b8v4C66npA0e8ShzuSsU1WbIqtd94VINAufXRQqGS5nO0WViptIzEnnuXc0xFJGScRUhYIdd44jvIRsM97%2F0JR3u7rxb6v3cuyCSLk6CakpMMvzKtjLuMYoxBNpg1zvAECFRV%2BOl5roj5Gdvs4HhWsRb4vjZW65Q0HUk3Bm8YDzSCi0tKkweXjwJjpl3FVzEUEy5twiQutwbeAD2%2Bh7PfQn5k4j9PXd80JQTTL%2BvovtuHDXn4NRT2G2Tm0A7w%2BDNfaBN71%2BoymCYYjKRFmHD86t5mzk6kQgNTi%2Bq%2By1hHhXtcea9%2FgvH1oc8BoPg%2FLsBBz0H2B327Nb0ANkT%2FpN5IMIJSbIiapiHCcWbGMxsyB%2BB9Q%2FcRhSXBlTvYeJhTSXho5FSENN4fj9WvwbII09LdHCVU6EhjLUfnFgVYK20DW0hOHjfjD4lCUVWzByFGrpj9q1WOcswL6GrO0C7Z0JWDQGxEKSBBhlXPBepkggL7eqww8vR4fnN5O3lCCQ%2FcuLmpaCtkMnUHC4ey1vD9idK7Qq%2FpHS%2B3EWVW1FCIVNSKaQZUu7r6UnYjnlLLDcES710JGsGHkEWUjpzGi8KJP356wrVBtjapzbyHmH5FrCgd64nSPIN8dxmnY1OzqPICyTwUYqWD7e6O4WrEAdP9VuZqLu1H6Uc3VLafMyx2bBAX%2BMvNKk36PT3CbWr6DmGxOb04VcjmrNjmUJ9YnaqKu1iI%2BAFlk4Ej8lM5VVBn63%2FpbwPcu5wK323to5llNIVlylhfQvPljGefPbsX1mOWWn%2B%2F%2B%2FcN1WEFQ6Zv8IylrxwJ5KEFyKvk0MlNY9RoxnJ2wvKkSFnR50Yg0Br37SPD0zFG%2BvkHfig9ErLE45Ge8t04jhU81Wmtye4EoSaogx1dQfoiM9EvMo8GQmu%2F%2FH3xjQ7A38KNVWrrGmC9Zu1KU%2F4vKaVKYsRLypMbNoizE851O61vxyYVYfSM0y70KqNcJ2pgq80pnNKedRmfkrSXUdWppMe6irGJLfNuaKQooizi5XNEddVMaeQf7U4MuZOtKbsbOWRZGRUWLd4xQT4KWfnq80PlSqhHo35lfKje7DSnM54sXCeXUDODLta7fsII0p1OwEnqLpdafoKl6aF6e0ppExT%2FS15IFMDVc1QzCbzcSZyUbLT9kSScTOOk68jERtQOIqVqIJuJ%2BjuDLcl3RaK93pBsNftQxFD%2BRGfRv5JdLu9va3B7n5fBDv9%2FtZg0B1mqA5YTB%2Bm7wf9vWEQ7Ab9IC4LEWf9XXwNej0cvuRf33ol8EpyCyXpb3X3tnp9MFemFZgbkizKKgQbodWNsKx7TB9%2BeYFrGtZWZchiG3mtgGxMVCTI1lMoKYQkohudfzjKrC3Ny%2B3t%2BXzecTA6SqfbwLZdxsn2At%2Fo%2BPTiHt8XTpBxIb5WKje%2Ftbqi1FShrUtXzOniJb6uQylJJWfYWcuPybobImzzyN15X9WHqpL2Z%2F%2B0mjDoTpPj9%2FiBYZrHG60Qjo6d0vmVDPlbXN%2BvHi2FgydL4aJdWrC%2BX5%2FtukmsKnFt%2FfPd2o8T7D%2FoJhIKcXMc%2BBKqRtDZBBVzxKZEUB43JmFv19tkCw3Gs%2BQ66Hb3g6b8v1jERXb42YCbl2MARNRtTlxloPT%2F%2B%2BK%2FX56t4U%2BG0i35jnu7%2Ff3Gy2Dsehu1q2UTS2AGBZEFZhTAncfLKKrKxT5qzL4%2FOL293b0mkY1xxnUBRUL13Rj9rsKF29YtKPSIdEICN4B0ge1vv%2F8P%2B2Fg6w%2B9jW5txlrhp527mpgG5WAw7Dcp%2FeYIpIe2NH4slmX00L%2BJ%2B2GDLy2%2BP%2Fx66%2BMn8PWH3W7j7OfulqfvM1%2BOGzuiPI8qcO%2FFgnugm6fdJ%2BVk6n5tv0NqOL3%2Bfw%3D%3D) |
| **Eritrea** | National | **R622I** | **~10.0%** | Assefa et al. [[5]](https://dummy-citation.com/citation?d=z%3AjVRNbxxFELUjkDhxBMSpxQkQ%2BzW7a29yQRuTQEARHIxy7umunilvfwzdPd7dmyN%2BAD%2BB3%2BF%2Fx%2BsZO9oIBXGwV91TXfXeq3r19v7siQmRG%2FYf6X2Iu%2FuzT1tZpxylyscnf1%2B3JMhRbMgrEtJrkbpIUotghIyZHCf27EWHA0srIiVOWZZg3L6QKQ%2BPfgrRlydbE1lJwUlIK6Nj30zxbU%2B3FL87zTepZSItVHA1e5k5eJFbirI7iq%2B3V9ffCODB0dojKjrJPgkyBqkVhz4JMCrxIgNqduRzqW2kVQycvROuVGc5Fe%2FTQ9Ajj1NuJ5x0oCR8yEL1MSIv6juiPBRTkTMhq8gB3BlpMi5b6RtkBv53WJqeNVn2Q6oopDGksgAtEUOfcS%2B6GFAAwBBd5NOoH7nGx%2BCn4lUuArbctCKzo4FtUoxghEFZPED7fAKE4MeekIwK%2BqWCrSbhQhzxQjFwxYNbTICVRSgvTB%2BL2A%2FYk0gB%2FGQWYyPwAPCaKJ3Dtz1bWzLK2kLuAGV3pfOA5NA%2FTQoaBnRHIpQQit%2BxYRgAiiM7DBRkR3ghIv3xUbShqJPHkj%2FSHz2XGLwsYps%2BQ6CplUpRwqj8ALGbal6tJ4u5U8FnlKD4CkDuzz5uZJ%2FbENOdZddhrl%2FiL0S9ni4r4pR6OqfM2VJ4Pc6F%2BDn0EUTNbbC9o89tSTjojyR%2FwjHaS0fFOUY6tkezBQQjqeFb8u02teSlYR2DpbN%2FR%2BuXwb6LrQnkttMPxdI1fLAfg5sfqYzEfoxVrBOoKR348OViPl0sNheztKieLi8m82o1ma82q81kg8mp%2FR3%2BJ%2F%2BWF6vL9aTaXK7b0AGipcPuzWr5dFFVy4t5pTvHul1uqssljjeDMIOuN9AVCReTau26vrYMgg%2FSng94%2FXMOr9GbK4wgTLnDwEFARb9He%2Fi1zblLz2azB8%2FdjNJOaw6YEDW%2BmMLoMxV6DwPNOm1mHybUpL7Ox65otCteVZaucTzHlFEwv7DXp9eUy45ALKyfST8%2FXpUiX3wFMxqoCUvrN1h6abj%2BbJyCw18vTjfCb1YmFzRjbZxskLIgy9bZc27%2F7yJ8WH6DPzHC7%2B%2FEZ8XTxXbwkvAENeGmTnpW35scOlbpThfen2Ax2MO3j6qi%2FdMQm%2F8Q7B8%3D) |
| **Nigeria** | Lagos | **Wild-type** | **98.0%** | Oyebola et al. [[6]](https://dummy-citation.com/citation?d=z%3AzVe7UiTJFYWNWMX4MvSINdKZEKymG7pp6GYcBTTMzmiA7gVGG2tmV2ZV5ZBVWZsPmpKc3ZAM%2FYxCNr8iSz8gcy05OjezeA2gWEOGjGG6svK%2Bzj33UT9cr3yWG6sKVX8mlsZeXK%2F8vOQL5y3PfPtC7e9N3391OvtwcvCanZeSLYIVsmYmZxXX3CrOGmmdct4xVTMXFr0zXnLLa7aXW5VxxmvBPARlJW0h60ySLLdeVsqpGjJWkjinNyUnLd4aETIpWGaqRssr5VvmDZ7ojWYyh7%2Fe9dmxqZWH63WRDOQ57GXtJ%2Fp7C%2B6gy1vJfSVrzyB967tyLLMhU1yTBS4EnHHQRucl11rWhewjbjw7H0TLuHO48UDfI7sS3tieDpXMOVxWtWRre0frEQkgIL3KmFCXBBsig9Rcc1cZoULFcq4z1XCLn8oZzb2MuHJ2ogoJj2vWmCbgXJkaCByev50dnL1mcxhWJFgjDw2CgGOEy1L5kmUaOGQI0bVV403lyGaoI7o494jmBg%2FuYaqUXJMYdFhJ1o94YdyrGxdesaXEucusRDAi4jnv3%2FO8zw5rJErjHZ7v%2FIpiETe8iZ7tHUVQqpRIQrW0JhQllJdSBNKAH9lFLzTsEun08OI2GpLUfGEsh2zL5BWvVB2BcdGn4YQJ3oInAIdDVkJUEjNBtKdTER28y3DnS2N0C%2B6CRcSJSFiUBsywtfn0lNLKdQsKx5qQ1vzRkC0XbM5hqbHGSwWX1irXDFK4rhmuE6vorbzkYFmqCmFDcb8cquC7eJYojFvHFu1DvHFPe%2FWpMIXHBjAr7GCdnoxvG%2BJEjtSY5VN6wkJ9FxSI03ONzBRondyn0KEpLJqkCdHJ7wK8hro%2BOz08%2B3B0DhJSQECkR9SyVmaU5rsyMcGDcRIuXoJfeLU76o9eouagCox4mFf4Qykz2hTxDHE1wIEKaTqPmAu23d95CQJA9NPbOVc6IJVrR%2FM36wz8kGDBXu4liDo9RSOJ3gHYV6w2yBRFS87iOssoWIJbSJ9OyRZZZTvjjZ0xWxtsbr5cTxnJSoVoRKxQjRYCAFQNPikRuI4dykoDdr2iqlLEpQf5XiotesgKSIdq0Sj12JuQMHYy2fmWre1u9ccv1yFNrKHjg8FwRC8m4%2F5254RZOGnhRJ999YjQt9S8BR0OCEN1goy8Bx3JLkRy1I5uk3mi6druZn8IA2T4zXQ4vnkzZGs7lDbQ99B5VaXmQfSjVtK1M1XnCV62djx7l7wc9MfoG1TzBBJhIZ1neH3rP4FI77Z7g23WolCBbwEaowibPqgJrzrWUfl2cSnMIg%2BiQtyAljU40uJPW5ng2OFgezg5SPVrbIMmXqUGB%2Fqz3CZlLRIKIiGg6exkevTh7N3sJFE5NXwh0Z1oGlKkoIpHB8Cv%2B00fXYzgiVSnAnvYWj8p1dRof1qyLnGVArnJElEFHdCFogB6ZMpkWQCZgUlsPxRqpg0U3SUBNDwBNoAWBIM0YZy4TRmimcWcKmqqdiJFBPoBZJkJWmA0X0qWB091pbroblttHL8NklEpKsI0CpGYXheuftTaumTfG2aaZ4gC%2Fe0AsBXDzeFOb3O7oqFPiNt3dW6uVz4vePClse57DSfQht%2Fgn7Fiqz%2FYksq5IFeBqtfSHHcT7fcmWOCRXxoayL9obhUeGURg7PXKKlJTyFOOUY9dCDhY5%2Bc4%2BdXq5yUG881vHfcPtQgQcn%2FFTVHziiTyVDzFrJULTGxZIKN1%2Fh4%2FGyMwB%2BXK48v5kSq4VumufhNqHPLWsGn%2FOYlitgS0rUwixR66OF%2Bg6T99W%2ByhxDvtMx2WvFggNwV%2F7n62p02n%2BVvpQy3kczfLQyw0y%2BA638t9YhWo%2BLzfOuSw3Wk%2FVpihUj%2BLyxnSdtG5Yvb5gkdnsIEseSeUY0mci%2FzHTAkHQmTCqKtfDzb7g8FkZ8MNhrtbIM5wBPJsTia9HaHcov4ef139gxqMxtu94WS8XZpGghXy6uKb0WhzuDkYDMZboqmUKLd2h4PNrZ0xRGb8x4%2BRVJGTH8FJqJ30hrvgRV0E8GIh66oJC60cVr5E0dUYUr2vzDFKeEpLFNcXoD6ImMkPVl%2FNSu8b93pjo6uOj4mi%2FYXCfBRx7cITesgGSq%2FG1NpoRL7xfIgFtm5qDYDxIu5bWp7jcRVVJU3%2BHhPp%2FrH0aCct7qInoT%2Ftt1My8gLdK8f6g9oU3%2BArwMXTX36Rqunqb3tx%2BbhZtWnB5Y0M1MB%2ByvpLPeLphfPRdvka%2FYguWlnK2oEGXR9Gr7rbJW7XAZpP9xcmbi%2FQTXNvGpW5P1%2Bv%2FCzHtEO5I1wUCx%2FF3Ji3acE9y1TsnBLzQ4t0ZTEcxzslkoeBVkuc5ecY%2BbuDeH71xU1jOYUgt1kZfZimbxPpsFvIf%2F%2FuX%2FK3%2F1xZWSmRmHu6xXC8tZu0jOZEmowlT15hYb5U1tQUXLcBzdDZm9gZcZCu%2Fe%2FCQQlMkiPrc3ysVVhUKZew9DUWlzSMKKjIvlzSSoiJk2L7y99nCO3%2FN7bRcLydHPnyLXiB70OKK97B%2Fw%2Bydn5DKHA7Bfenf3zNngxu1KX%2FN6dcpBkHxob4QZF0E81J57uK41O6EFSQLzCc9dWXN%2BWOTtU3tvgvlXxBH%2BB%2FoH3A1Kv%2FAQ%3D%3D) |
| **Angola** | Multiple Sites | **Wild-type** | **~99.0%** | Dimbu et al. [[7]](https://dummy-citation.com/citation?d=z%3AzVY9byNFGM5FAtEiUVBQDAIkDp0d2%2FE5Ng1ynOQ%2BYt%2BZ8%2BlOQjSzM7O7bzI745uP5Jzq0FFTUiM6iquoUkfwd2igQkI8u2uSFFkqCiTbWs%2B%2BX8%2Fzfs035xubqXWUkdmUp9Ydn2%2B8m%2FPEB8dFWG1u7pIyhrhmIVeOL1UMJJhKUxJcrFhhDQVom4yRZ5wJF0UpDF06obBiMM2U8bESgYkrVZsyEZ1TJugVi15Jxl1QBXkyZFoJL0%2BELRIyPJA1a%2F8rRoaNTWY1b7NJTlrCAjulkMNnDIpFA6Wlho8AA3PNfWElxYKlXAtacodHMqlCgDAKYx4RkFGaeQrKlydlmLvKZFFpfod9xcmpO4wbyabRSM4WUbO5sydkBORPlVMsOFW5q%2BMoYagy3JaOhUq5CUCv2Kfj6W0GOsr3PgKVavEyNv4i1q8X4y9vV37WrMJgSV9vyCRfeRYs494r75nQ4EiUNEMYkDhCt9pm1ZlTfmkN5NpsZrUSUXMHIsF0DfmUe7ZUDpYLOIigOwMHJ2CxIOGsR1xagwpWcHesnG9rLgAU6djDq6zX6Q1anbuFsCZwxO0emNSeb7yV8Rhy6%2FwrTcUS6T%2FA1zq53e72FXkf1aYKFLR6%2BckYfFeukrJSxhn4R%2BkAySRXhV2nOT2xJXnvj3XpyFESYc1%2Fj2KVhheqLNqUF6RXao%2BKJKqMgEDPlXSWPeEpVygMoN%2B4QeG%2BdXBSKWRPbKJc4E2y%2BeTiRyNJ2lr8xdhwFMHFG8%2FZjAQob1Q8UCCcHK8V7YRDEJUMtbKsmtSyCYpBCVVr0YHS5AuoNcmn97hDv9Xi6TSecd0sOzbSXfxUy%2BaH4B6cN0pnc1UkSq8jsaWfizecPeJnHEYaYT%2BETYvP2sueRfSZ9Y252L0SLnaVLsAYe2h9s4esRMHlOi6DNj3CuWGPSIXGbKRTnkS5pilfoGFyNms3BjVVFz%2BbWnh5jyeOMBzmFt3OZFlfN7s4RNeoVa2lHkZNjdSqedli6xTv5S6eUKKAqsHw4oSyS%2FEFLzjG5aIxeHrKNabUGYl1KPtaUrPxaRQipzUxjzjSxZ432k5n1nHy%2F2C0%2FF8K8wBNptai%2BQFpW6CKGpM6w0wkzKy6i6sCYAeOY5Q3qRzNNTbNL9%2F643X0RzOwGv3ZZWLTnPu5TH8XJP35xttCWgrdTrvb7Q23OBftTnfUH7V6HUk%2BMa%2Fw681r6nQGg1Z%2F2OlTt7PTaQ16w208jYatwd3RQBQ8O9rujHaG3WF%2FtJPbpTIcE%2B34%2BdWhXBYk8%2B3t7mA46PRh9TH%2F46iaf9X4PML47LS63Vann6P%2FssgzheQXy5ig1TH66ml6q0L78sNxoRyGumELK1Dg9TrF5ClnZznwV5mPSVgtS2KOSw6FVk%2Fx9xYGvbLpITJ%2F%2FVgFrvUKstiEWFe7q4mNJrz%2FAOsrxRpDhcvnuAH46vi99byeTDCAyQAAm9pTtn9tf4%2BvNt30%2Bqa7XNHlUw9Ep8EuSfjXyEMqbYG9gSASkrxfAbX3FdfYnAuALJeqStFyshZJejuVTD5TkgSs4yx92u10Rt2apA9mmGSOOHsCRYwpLOByl5RbA2XgsfnUn1%2F89tfXP2xsbOSg65pt2dvZHtVW%2BvMyA4LVkdxh%2B%2BaEnDWoWnBWWXwsRFxWVxEc1GL%2FIZzuYFAH8tHMelwIgm0l1oFLlJDCXajekNUuvIYqfsf%2Bz6hQ7HfrQD67r0ovAZfEZ%2BRw2%2FNsEaKkNa79JUlc%2FaqSvgR3%2BOvNKevViT%2BqTFYqsmyBd3Cb1C8%2FzkNY%2Bs%2B3ttDubeuyrZta%2Fri85z7D3Qawb%2F0N) |
| **Mali** | Bamako | **Wild-type** | **100.0%** | Dama et al. [[8]](https://dummy-citation.com/citation?d=z%3AjVZNb9s2GE6HDetpwIAB2w4DeCmQALEiyZYtD9vBTbCla7qkbYYCu9EiJTOmSJWk7Lin9LjzrvsD%2FQk595%2Fsl%2ByhZCd2Uzc7JJaol%2B%2FH8z7vx9vrnc9ybUQh1Odsrs30euerCR1bZ2jmFg%2B%2FHRnHS%2B4m3Px79besS55T5YxQnOyOTvYIVYxQiNhaUcchQkvNBH1dewlqOMmFsa4j%2FasznLqSK0dgkNQq02UlRYZ7jJRUUiMoEQqPakG4YrwUGcl07c1xu49PmayZUAV5RqUIyJGYLJjR3noprFBCwXwlKm6W5nePjkc4Onu%2BR4QlVFoNd%2FHruIGzYrbh3JqazphauAT3xsILakV8%2FLRa7JNx7YhWckFyPieMOtoESWdUSDqWHN%2FIyirheY7osoUPytbjzks6oQYejHKD84CcTziCxUcqSqJzGIGb1tVsQebUEqe9C5XXn8FFXJG3KiF%2BY2fGja0tGZ3skxoBFN5b8kobycgxR7gTcmoKqsSbNpbdV8ene6QXdxhtXJuJmSaV0U5nWrZerZtxE8N5h2nLieGFQP7shnXv6tJP5n0enWwg58MjiJrpUrzhbH8p2%2BKvK64IgOMS5BCIb83%2F49OVj7mWUs87dXXjJcmNLkkcRl1vEb%2BJt3OkFS9qPtZ1oeuGmefaaKb3l4TxkVVGlNQ0%2FKq0ABWXSNMs4xZPEDk7fNHJtDE888QcMf66BkXJ4SoHXvEZIrDCaamL5uwFt5VWloNmDrRYkDgNyAiKHT4CrW402EL4CkigJCyZcySaK4NgPUxzgbxFybAJbIU1NWVjPkpSfw6ocbIMrPUIDxo%2FYibcArDjnfEMdecJ3YDWT4PeI7I7TB6RwyekHwYJVA%2BSINnz3PWuRx6PbpDeSkVBD0JpEN3IxE0J3%2FoFnwbD9SuDOOj6O0kw2FQ8DCCwkkoaxVESxB9o9lzercjPJAzC3l4TIdDxuaLEikIJz09kj4k8xxeVcY%2BHz55H%2BSZ3h2cvyJi7OQfNNpz1%2BscaCCNdcdqwzB%2FfIVyTheFgHbJht4FsOAy66CtremeWpL316AaDIPUYAJhGEulCTD%2F5mEJA6Q0Oe0F8ewGSHjTYu6t60N3IG3D1qkOPXKN6H9VpK8SNukJ7WjMEIuZoeaTUkmc1WIcSbBBCee43na1FVulPQOuhzO7H0mfgfjyHG3j2GzyjMPwg5rarDUG7NWEA3ILfX4PU0wSINYACwv%2BhG3o%2FmdRbvfEG%2B7ZjtAUar%2Bz5eeY7l9SqaHtiFPuYfFIf3U2bN7qkvP9WS%2FQGO9Fz310nvrvcN%2FZWI%2FnjU7tpM6t5AovLVi80sIZrZ5JaP8HrkuRUZgJtBY%2Br5mJ9OE0zlW3D5OwIPaaIwzjphGmZaeUw0Lh5onJ9vfNFQWs30cZeSYGmn7lf8KcNS4JuzIW1NX%2FAnXCSa%2BhsuuFvusZslvlMe6e%2Fll6hERi6UPIXNhWmaMn9xpLTUsgFrJcUQ2nG1cVLXUu%2BwPLAgY7kO3elJ78LihTIRXujOMYp0%2FU28eLoQpTs%2FbtWOn8GhCzdJpy%2FpAVAWso%2BgeR0u%2ByvyBPTraw8nHCRTclpjbm07Yb48%2F27ohblyhkxKsGkon7%2FbtsNILPyphgph1G3FZbNOMVoDEwkXWyVz490XY6X7l%2BcFnoMU%2BRp0MpngtnrnS8zpsXl91EYRFHaP7BRPOz2O2GUduLesN%2B5ZMKO1RX%2BW%2FVWRL1B0onTQZKVtLiI02ES95Ju3Jv49YBKfjl9dXvIqxImin406Kb9kFVI4aQbxsMwDdOLhlMNJS%2BwFKSdKOyESVnVYyksaqFl5YMmHvVY6GeoqUMMX0PlFKUG7mX8DyMvTybOVfbHg4PlkL5oWRmMhS45y9obAab5gd%2BSpwcVyw%2B2hlpg9XOLygM4RVmKTPJzvD5AI%2BA6fwomrB9zLAxyAVnUHkbY48WhX4C%2F%2BwG787LTMGx2U9scf9NWz%2BU%2Fo7UNa7lJYem4t1MsW%2Bz2ZuFH8e3WDpWbO8zHm8XaKu9bRe50JTJ7xTwID7Ma%2BO6t8AVJAm2K7eD9Bw%3D%3D) |

**Legend:** This table presents the granular raw data used to calculate the pooled prevalence for the meta-analysis. It highlights the contrast between the established resistance clusters in East Africa and the relative stability of the pfkelch13 gene in West and Central Africa.

**Commentary:** The data demonstrate a significant geographic divide. In East African hotspots, validated propeller mutations such as **R561H** and **A675V** have reached frequencies exceeding the WHO 5% threshold for "partial resistance" [[2,3]](https://dummy-citation.com/citation?d=z%3AzVi%2FbyRJFfbecQghEZGw0p0oCZ3YRZ72%2FB7PitNp%2FGPX3vXYw4y91l1W01XdXZ7qqqGra2Z7o%2BVIkAhAkEBGCtJFRBtvihB%2FAiLjD7iIhK%2F6x9g%2B3I4ILrjb2erXr9773nvf%2B3q%2FeLv1XqATEQr1AVvrZPF267uS%2Bj43hrMDmvKw3Wz3G82O9LVKEzG3qU7MHz7Ea0zRmLvXAxoLmYUXaxFTRXkoVlzxkRSKs0RLvnWXbWylMKVtcKq5lLTO%2BOolVwtEYqgqfY85T2rNg9lKhFwWpmJGYyozMvPqzNnnkbaFcfh5JDKtwjpTMbYqQxCvy7jFeUQTXzikat6Ijuj8BijxlAZiIciYsivBal%2BaWl%2BHurolGCnh87TWeozq0STNyhxOOI3I09p0oxlVDAVnhTmbIqE628VE8MTngeCyNOeHOWR1VX2mJUO2ZdzHCb0nEH74msdVs%2Bwl94Qhzqm0ir0GCmUQkola63BC%2FYj7urCVY4%2BMYMvffalqO0YcGp9KqlJevLQYKarefUkNGdWGH56G%2BPOqfCM4oMqhVGc8pirEDWVMzzlVjRNthSGntV38gs7R5KX7sUBO9dOEvgxtKkrjFxR1FbVFHdsE6NEInVm2zFgr8XNb734vgSVPCuurSyGloDGZ1mITjy1mlc5pSNW6TCHalzS5b1SCsXUjXpb45N2fdX1fXrCIZnRhY1oGFRcUgcoD5bjuPTWei9RmNrYJj7Mq%2BxEGdC7r4zqi0vBqug4T4d9DJsGJ9f2o9BydUh0Lclla%2B4KZt1vf9pkWrz5qNb1Ws9XfMa3uoNPoNIfDR%2B3W42az1W03mkyYuXqD%2Fxv1hdgYuF%2FdRrfbG%2FgxDa86rV53uNvv9PuRXnJFJX%2B1uLw%2BZMtYsKjT2e13d5utK%2FhCgR2fX4HPW41mt9HqLRJutMWEXyTy1bMoTZfmyc7OMvY95c%2BFp2TsKRF5oV7tgGCEL7nZmYz3W0242O0Od5Ys2IFBbBqt3W530Gx5OAmNnafZEqBs8VSkCOsvI2O0L2gqtCI6IBNJTayZsDEJqPTFkib4ueDSj1odMu31W0ck5Eo7L4asRRoRxiXNOCMwpUaknCAW%2FFQ%2BJ0KR6RrkQZ8QCveAoiExOnKbGKFCyRs0ibcJ1g0S4NhgfJukESZryTExPuFBIHzqZ8SklmVBqpfCN2%2BYu%2Fs7vgUujQoX1M3TSbhzb%2Bmut%2Bl75Tb9PgbNpAn10%2Bz9v%2B2N9l88m55dnB48IRH4hfEVl4iZEZRCmDTPKNU4T8QKgK0AABAD%2BjwWyEeoPHoSU6Tt63ipFXJyJoFITNpwO5cgR5rGODcEscDWTR71yD6eIlf5tbtuOCexBuw5OpkDNv9NQqnn%2BVshCmi28cCXFgwcklGAcaDbZK2tZGTOyTLRGKUYkfseGR%2BeH50dzJ7ghYD7efXzYgpUgWI%2BtDX55cYq9CV5RMkafyYNo6WFl1uBXQPyGGVmhJsUY5sCOAfHHW0BIggAR8A9cskJX2GF5Ob8FVmJlSbGGp8vUzEXUqSZQ%2FCWI%2Buah1ACqgwbwCrkeCPBmwCCGkOzPIqqS2%2BWr5GgV91NeMiNR6aHs4uTc6AQ2zQfAeMARIlAhWjRUAnXgCqV2TYJEh2Tjjf82IHfbrZ6rj6tobdbHQy3CbMOZiSfgG%2Fccxf3tccg4aByhYYuUxr1B72XebT73f7wM0KlhDgy5NHkp00PXZs%2FmpBPiPtbd9tlsnTVQmNmjz0yK4ZIWUCrU8FwuQbD62QZCROjw7A3Fw6r67uqRF3MF1gBDIXF9gCAGARkKhBCRpgIApyie%2FOs00ibfJhn2sIVNSkZmbxrz073Ty5mx2entzCMUQI0HGBY8KRodMbTss2K1OGvKku6qSwq8ugp1ASgmgM%2BBP2cLkEcMzAURyM4R%2B50gqCQb%2BFshmeuqRxU%2BUwY77F3l0ruxU4lYzh5cqwC%2FXbrg5AiHSjmN1LES5DAU%2FynE97uep0hz1n5eyVN%2FvCUr8kh0Ha3PAcvg9Pd5WPOhA%2BPwQpzEfOHD36x3Fxyon0Kd2%2B3HoBFQz6FzHDbCC0FNpjg5OG3fh2Bbsvfv78l4n9zh9Teo1IssLcrrcL3eJrWCr7gqV1YVpqGWHeZXdQKD3684JWtPxb1C%2FcFTW1S6jdxZiCDUnJeu3KhDrFx59caexYJ9a83vz3GaaJrZfBn%2BC6wm1UdjKkBkLXfCGdLUQYUXFIJkqpXteia0hRSe2ETuqh1C2WfrSsJEx0g2%2FsUZ%2FBCQLhUivAQX0%2FAvi69CSroo98qJSKwAVT991h8xvDdoxsjLK6svEIfxtDylktyHCLK%2BizO8Bh9VwEJpVCr3PgRlmOJuTjXJhL3FYmNRbr51LptHGB7Tljw1Q1RZfPF3OvvKH4Va9rGkh50%2B7e0VBO6pdEdDDui1et0oKWa%2FUJLtZvgvmZnMLippTaHpZbq9nqtdnsXvs7oV%2F%2BrqIaNdhuzBiWOWZujEZfYYsKALQoqeJBn9%2BojtJpBy1qDyTLFdGPQS%2F65JckOKumxXq89l1WuP5BqrryqdE8Pn4836X5aCrVzraX5JE3wZbvRY%2B8tqof46wNQG9foKcVuHvPUkTNsocuwxPayfW1V%2BvD9P0FU5HTtc3YJXWPy8x%2F8qOSuzuEKq8FRJAhrdL23G9MNAY8LGeIovpANpdb6JcoXMO0UDa6dC0a7OVD6iGPSoop7TfEZWpjM24PcJqq4EWfBeavZHLYKkD%2BsbsP9UAR%2BVCxAR37oHuODgf%2Fz6b%2B9o39ubW1FAOiGb9YedIaFl%2B7EVdAnRSTbIOcVGlA5aeVkADye%2Bb5d5hsJB4XZ%2Fy%2Bd9u5gtwjk8QT%2F0BBjmp1kxU0%2Fs6BpLKsiKRSCJwEXmLuwyu2Pf%2F3Hm29ybt32oFcE8pMjNAhYweVVDcOtqp1v5GyV3O9%2B9fe7k%2BuW5f%2FxlDKhpQ6hqU6dcqHJZo3mPo8x9UDrhsz%2F%2BE6Z%2F3U2WThN%2FxL7H6g8%2BC8%3D). Conversely, in Nigeria, Angola, and Mali, the parasite population remains predominantly wild-type, supporting the continued high efficacy of first-line Artemether-Lumefantrine in those regions [[6–8]](https://dummy-citation.com/citation?d=z%3AzVrNbyRJle9u7bBzWgntSgwrVorVbmtt1pVdmfU92tWqXLane9p21dg9jEB7icqMzAw7MyPJj3JXcxkEB7hwQeLEAXHjMOLAac4t%2BCM4Ie6IC5yQEL8XkVWVZVd2N2IkRpppl7MiX7yveO%2F3e%2BHvfHrvga8yGcjkgXejsutP730x5PO8yLhbLB88OJQiSSSPWBGKjKeiLKTLhO9Ll7tLFqtEFng7CZjMGWduVrq0GO%2FKhSyWDKKZSPJSL4GIzavKZ26ZZSIpoiUrc%2BExnhUilrlMZNKac3riqnguE15IlVT7L5lM2DgJVMQtNgll5EECu5FFiD3LQrAywUtphD0KCJhFPI%2BVJ8uY%2BTxyZcozfJSJL6AghEJYDg1kIiKWy0Lk9ITUPBRJUIqIH7BvcJmJA8YTj52WicfZZRmxWaYWMnGx%2FkZkghWZ0NsZPcgMQeq2ojIWPk8KWC%2FY3vh0n8Ed9H1ewirR4qQb%2F2Zpvr4cf7Cv96m8CoHkPmfIPL7MWaEYz3OR58yN4COX3IzFMIlDdRWpQD%2FLRJ6qBOssdqYi4ZYRz%2BBIeNqYfMNzlooMkmNsUMLdAXywgBdj6WYqh15RBFewmGfXIsutiLswFOE4wleB03b6rXYvdlVScOidPUl89em9twJeFqHK8o8jGacI%2Fwn%2BV5nXseyukHleigeikEUknj8cw996qzllyjiA%2F5E6sGQSilhVYfYXipz3zjiijTI5LyEt%2FxGS1Ut4LChpfR7LaCmOZDwvRSBhQTQTXqbYBfe5QGLA%2Bns7XnisMmyiXwgu1FxkBW9aG05e%2FjTxpKfM8m%2BOE44kePlJztmZdOHyxhdPBBwuM25eVBOOhchkvEZp1fRaMEEyCFeYt%2BSJiGQe47Wm9f57PMN5M8v90%2FIFj5rXjhMve%2FkzszZ8Ct%2FD542rg5mI5yKqNFG0z8tPODvnLziENJr9PmQq%2FFftcqSgfaDyxlgcbhbHhyKK4TH2vsqbdwjICu5VeiU4pld4nrBzKYrGaPinfF56lZvCSxyYkJ1ZjUqdipe%2FSMzi9D0%2BzySKw0zhtDOP8mv3Fk9xasTSvCXeLyPZ6FoxoyNWhfgozMqFnAtY1SD4ciGD9fJLHnOUy8tG5eUzHqFKvZBupcpx5Mlm4ael64aycsw5R7jYR42y%2FTOVcZmvbFT8FYl5gkMmqqXhiYxUjCxqDOoZaqJEzTKnWCcAO8k4SnnTK1ezCJ3ml9%2FNryvtr87g1TJ%2FsQ6sH%2FJ85vl%2FcKWXf3rvC66nZGG3Ldt2ho84d622PeqOWk7bk%2Fk8%2BRj%2F5sl3ZLvd77e6w3ZX2u1Bu9V3hh18Gg1b%2Fd6o78Y8uOq0R4OhPeyOBqFKRcJR0a4%2F2jz00lh6Yadj94f9dhdSp%2FyPV7r%2B6fJ5hfLZbtl2q90Ncf6CkgcCwY%2FTco6jjtJnqul9be3zfx%2FHIkNRT9ilcpHgpp2i8lDtpIK%2FDPJyXixTcsw1%2BdCNxDP8eh%2BFXij%2FKSJffywKHkVLrEUnRLs6XE5UmRTvPEH78tHGkOHeR0AAuX78L1W9nkxQgGUCA9ipumHHtf493nS603qnW7do%2BuTA0X6hUunmwBpf8D0Vo29Aibn0eFcbqh4LHqFzXsJIaqrCx5HzzJK5M9BrwjPhSRfS8cx%2FZrfbI9s46StnqGSZ5OwCL6JMoQFTL6GugTTI0fnEn%2F7v93%2F%2B%2F5%2Fcu3cvhLtqsj1n0BkZKd0ZRcBlRpMDdpwsZKYSZC18piVOXbdMNRTBA7PsMzTH7veNIv9xpnIAgkK15iqDL5FCAljIdEjdC2tWlT9gn2erkOw9o8hXHwvapQBI%2FJrMgPZydlmUnqzsOk6lB%2BinU3pt3NNf7Q6ZYwJ%2FpUXqVzw6Am8DTUbP%2FzMsijR%2F99EjHHdLZcGjXUf%2BmnDu14BtYPb9u%2Fj3nzf49215OJ48fe9i%2BuH50bvsGYDhvMw8YCVkPyqxzjuAqVzmwDAEJst565KH6DA4Aj6dXW2fBr44y8A66CJ4twZ1CbDhdRQ7wVCyIAUh9kqcRYK%2FKU4g4WiAvyr4hKBRXDW8W4Pv28j6LpTWEJUCr2vISnfEYwXZCV56QNPAl0VIz0MUC0AkYcFu%2FJ4jXssKgW7Ju7NvE%2FYlT8ADqCsu0npBboNleKsBpeeoIBUkB%2FJAF4TGCUtVCkhLGQsPHD97PD26fJfNdJXDi4QlUxihQX1gAPkaLufLOC1UnNOe2zxh5Q9eYKvQJLoLGcD22P2UA8McrFQ4MJg%2FdzMBYwxGn1k1zS2cMwQqwnf4faPXXaowPr2F94swU2UQQngovJIk4IN73SpTtkA4C2ixBf4jjiqBU5AtmXhOQEg7Jq%2FzBoucQwxB4FUUSJ1ou0OhFdxEuNIlVdESuYssopzQCUv0DlRibza5oLACkiCF9ZkQmXqBRgv3lJnPsVOaqUKgt7O9OE9tY26eOvuUVfStWKCFVqfCy8qgfhzisqjsIdKyVmy%2B3PY31kWFvP0ymcdsbOtl9j79plAlKCd8hEbd7JJTziWVXpyZPBWuRFob9cl0SCrnqZEE6wRweoJKF1js4vjyw9NnSEIyCB5pUWppurV1TFRZIOMEVFwgv%2FDVqGt1H%2BLMQRQy4s1IHQ7SZKZ97rGe1X%2BIBMCrt1f7XEYlQrl3OjvZZ8gPIoNjvxBI1MlFjQwesEQhUmQtKYvlzCVjyd2eKMxT2ot2Zf3Bo%2F6A7aH3Ptw3ESHsuMASOqGRHhHgI%2FJJeiWPdIXKhEJ2gT6zUFIubcX7BvS9RbWb3gbGNXlLAWPnw%2F7X2d6oYw0e7hvyrR8f2U6XvhgOrF6lhJrnIoMSFnvvTkKvU3PtdCiA3oZzgog8RTrSvnjFgEqzPaXp3qhtOdiANj6ZOIPVNw7b61PYkL7HOWisKR6UflRKqnK2GS%2FsnU2fGC1ta4C6QWeenES%2BEHnB8PVa%2F9Xkodeye2yJgwr%2FBkhjHMLUQmpCqyrr6PhWdqFtotChP3oMiA%2FRTPIl%2FlnGqszZsd1zhkfm%2FKosRRGPTYFD%2BjM%2FM8KWCCgSCQZNpueT0w8vn0zPTSqbgo%2B%2BDKHohmQpUkWTfm%2Br6KOKkXt0qtMBuzWC2T6qptC%2BWbAWWEqGrKJEqYIKmJdBAO%2FRVgrwhWZINImBJmSqG2kssw4C0vAcvoFrkWB4m3xscpsiRD0Lk58godNOSaEdveUyV5WRh9a8EMwvCzpXmHBo69alVrffGobRYhGYVmVudKe0VcGuNbO%2FdczSMWOW%2Bwa2qxUqfl%2BVGfxRzVO%2BlK4FnipYoDKQDYQmEBdgI0Qj4IcsL2Z48uX7b4GjrD9vDWK%2Bt4PETZdijo5dDSOe4mP6CkYuAx5V1C06KRM85EvFJo3kM5jewLXLivgHY1RxDjzYsNob44hX0qdRecODOWITNPJVdxxVY4jg66IAgW4k%2BuExAM0NxmYVtz2krEIqNusdlT72rqSDvIWvGE75lwjbdaWKOuRzrpUBArnh1UsVr%2F2j4bVvEa99%2Fq8a5Q77j3LbGXWQOE4XydMegrfW6O23pd0d9FrOcNCrcdduF5TUtu1Bp%2BKuI8dud%2FqDndwVYoctZ%2FR67pocSgUywCYEonh0jdRHIrriQ2D06QqjV6fjyqSoBU6LkaSGXfgNNeQRjl6CrvUo9XwD5Hea%2BFmQ4Ld3cuB3vlKR4E%2FGGnysoPbOWfgr4C%2FViN2A8w66fJem6FiYiRBzc6RBVYdpAr7GEms4QP2pDpjMyPbvTrd%2FJ%2F77N59rYjocDI0i%2BzOQtRhAlWKJnT4AcDHNiIzS2ecLgoToOMa27%2F58CtM%2Bv7Z1ncGadCMvwA%2FJLr0GP7ei9myVUMhtY9y3fv3B7olCtwr%2Ff11wz%2FQ4ZGypCYWRTWlOMp9gSEblcEPJv7qTku88yQ3E%2FB8qYv5PNWL%2Bpc3w6bcf%2F3A32dzctGBJ%2Fa6Fo4nrPtcC6K442ZobNx7UmCegWSjHuL0gVEDbER7BRVCEWQZqA86GtNiRDJeo1TUOju1TiTnB6qrn6PEYj2a47qFbsygH9SYEjWQjAkdAY6PcG92KHWAuAYqREIYVN%2BB9BbgskbkFqACfR4DbCVvtuilZOycWhplRKjIuYypreh5gyhBhWT2LAPqA%2FDVpqVfB9T6E7ADfxqcH1T0T1U5UV8CpKq%2BnWcAT%2BcLYsvfR4%2Bk%2B6zot0FZSbSEXShMwhasWo1V9G%2FBTIVqeQhnM0DARP40B17uTqpWe4GGKYGrdc3qqAKuJDrwQHoi1Xmv8T%2B2RyDUm%2Fwgy7Kvp%2F3i60tEwSSLnKy0Bq1VMI88O7YifPdrnCI0I1z5z0GlVmpOnMONRB1XCGCYMNkE0PvFShRnQytMGFeqdiVtumOV4RRwndeJYcf01FcR5N7eBhPxJZ2cIKgjBVIvgrY49aEh4KlD6ak5PBUQ10ahIjN0bacNWvkYN1dvbvaGe%2FmKukWEOog1bTR9ShR%2FmVpjYBEA4Jig6obXT%2BkOiw3uj3kM2ecL6basH0YOe1cOlaaJVt8kfHWu4WWVbXSwaWqDl1RpHH%2BGNXtBpMKq%2FMnCsDr3TswbbgkdglOtVPS3Y7lnOLcmUy3sp%2B1%2FWttpdM8SAdzQZ3qIRnvQrOLGC%2BvWpgKbTc1HcYIK0rSzJn%2BMSlMLlDHWW0eM7CaejMBrUXQayTC4bjawO6kpN7iJnw27dusHAGpIP4Bi9EuGCTf9DNrXhStoQwwln8wJWktOw313Rg85W3OBXEg36XIk%2B0GiF6NZCoDzVNlrNJOId19Sap1WepRlFo2vJle7rfUkReL0%2FR1v%2B7Gt%2FAvvcstlUtRHSrrYYDjbO79dcSmkCj2mHwoVvIBtyXxnUjdxqhPZaHzW4hoR98EwPuECYA1MTbYdsoqA%2BvBs22rRKefoOQw80hVBP0oqQqsvr2t6qJe%2Fu2rrMrPoJdqxKvR4AQLXdM%2BJVcdFDYl1M79DoHijLm9LontVx3oRGf3GLDH9%2FB%2F88wkVtdTF5ifmBWAI8NHPKcwy2wYer6%2BPgMZ56qmyklEdXMvZWV%2Fm4lS3zvPkm9pIHcFK19glWXr%2FizwkQp9V9fIQ%2Fx5DuNZuW6EuN183fePmzoJTxShk5xkgMf%2BLQfIEPz6y0wV1%2BgVbX6JZtO%2BV4Dp9gStB8B3%2BkynheqX81DdQcW7Gn1UjB8OV%2F1Hz5y7chqA1W2x31W89302V98%2BsMRz2n2%2Bs43Rp73jwUaYwtgj6INK5%2BKy7ddkbtYXu4zaKxmd2m4c5fzZxP35g5E0q%2B3smb16Z%2BJnfH%2F%2FbKu%2BMfj2sIq0JSNOd%2FXaWoSmxzsVjPG81o%2FfZ1zu5iUYPyVCoqlvxxjarsv46qrJ33Fw%3D%3D). This extreme local variation accounts for the high statistical heterogeneity (**,** ) observed in our meta-analysis.

### **Supplementary Table S4: Methodological Quality Assessment (Newcastle–Ottawa Scale)**

| ****Study**** | ****Selection (0-4★)**** | ****Comparability (0-2★)**** | ****Outcome (0-3★)**** | ****Total Score**** | ****Quality**** |
| --- | --- | --- | --- | --- | --- |
| **Uwimana et al. **[[1]](https://dummy-citation.com/citation?d=z%3AzVZPbxtFFHcqQD0jIVGpFXODotjx2s6%2FXiBN0yY0To2TNlIlpD7PvN198eyMmdmJsz0V8SUQJ64g9QPk3I%2FAlc%2FArScuvPWuaRHdnDj0YHs9%2Br03v9%2F7uz9ctq7F1lFC5pqaWze9bH2sQUr0HtU9yDHpdXsb7e56Jq3JgQy6AxPby9aHCYQ8tc6%2F0JTNQOb3%2BWOd2u70N5G8D3gdc8o1mlGYDFGJXTS5Ax2fWx0y%2FGT2j79DK4EtL1srNIMEx2ASZFYUk%2FP5iE9ufPAs1bB8TnVp6WgS2Mj%2FdJOhykBWmsQxZKSL5PGcMjCACZ2jwR3NtyhnNbbehc2CJl9j4yOLWkMT%2BOwJminHxIOpfQ8RXSM8Pj6nBHUFpWPIQBfiuNMEV09TGypw8jSlwpqkCUrDYAom8bzmTScpOMlRb9SZ7sPkraBkY4hpSmII6oxUo9E4SJvY5S3xjiGJeSN6yHUELi9qDYcIqbjfKDc9BqO49FQFV2MW1ISdjgidxJhQ13DcW4SsKasPrFastuZ94OAKIrj3HLNlsdx1V9CgE9DBqOcchZqEVtSITkYgU5S2wuphR%2BwwFl%2B9NI0VQ3teggaTY2U03TFgXr0EL3Ya6SdHCf%2Be1RbxPTBllJrAQ%2B4uvqHm9A2CaR%2FaQF4cNVbxQ5hwkdfuh8SamruJ6zIJOdXgh8B5pcakDoPj6EHKlVmXzNAa%2Bj40u7%2FrGImuQp%2BdktYEmRg3xiYbBu5VmEACZl5LSHc1uKtaJR6GssXrFB%2B%2B%2BtU21%2BVjlUIB05BBTSqrRgRnnqOcNdmZ4YTyUIQsOMyKpfodbtCJbua1D9rjsrv2HMkrhkl8GKRMa8%2FpEdiMxGmNjlPwIxW%2FlqT8ZesjqSxd3Iq6nagbbaz5aLDZb%2Fe729tf9KLb3W406LW7ivzEvOBvb17IDJKzfrQ%2B2N7a6G9spHaGBjReTE%2FfHKpZRirt97c2BlvdiO0eweuzxVJY7JQz3ilRuztoR%2Bs82LlieLBPOHazMOFZnC5XzMpC08WtI8jJ8h3iwHheKWV2hI3FPoLO06lDbwMPh8dOXzxI83zm76ytzTLZMXJCHaOzjqG0k9jzNZ5NJDX6tdFwN%2Boyh63B9tpMxWsMyHw72hoMNrtRh08SHyZ5MSvjOa2tTvjvCtNAGz%2Fkqn37GHPQumCslpSjulvs2mDyGyu%2F5w5jrlYjUZ3yavWL808%2Fq%2FbixW873ltJC3GlnhHvuMwqCpmIgV3NwPHjFLVMo74Yr29E%2ByJBY0tiXswpT4VCDQXvVoaC57sFM%2BJHvlCQEeM5jzK4I4Ddc5LamhtZrwpPJtHYBpetCl5%2BLKPczLgqco48zJD7VwqMY5IgC%2BHzoIo4tzOS%2FkeulljZjNc2q52QgsEiRbZKhTiWVIr11aCuIJPe5gKT8jsASd7EfBafRN3udlSl9%2BaQO4UXhxizIW%2BxlPny20K547lYvbQO%2F%2Frqzz%2B%2Be9ZqtVLOy1u%2BVW%2Bzv115GfBbhmbeFZNVsWfOyVmTsTQunNLjIynDbFlJFez%2Fk9Pb2tyqiNwe8SrOQC7CyDd9G0BTXtSiOP%2FoYqScc7DU5r8%2B%2BuV91jboba5XRL7cJ88vXQtdCwz%2F%2FitrJw4hL3n5pbjp9Z9Z2zvEDer0fz4GRVbbpFgVR2FRv5Vvvn%2Fh84DnDUdLlWV%2FXQbu8t6yy%2Bfz%2BX%2B7nFv%2FTacv23xavto%2BQec5RCt%2FAw%3D%3D) | ★★★★ | ★★ | ★★★ | **9/9** | High |
| **Balikagala et al. **[[2]](https://dummy-citation.com/citation?d=z%3AzVbNbltFFHZaQGxYsaESiJFQRYti5%2FoncVxRVfkrSdvEIT%2BNuhzfmXvvxHNnLvNjx7sUNkgsQLBixzuwyjsgxCMgdjxAV2z45l47aUrNigWLxPbMmXPO951zvpnnF7UbiTYiFeoGG2szvKi9m9GBdYbGbnLzl%2FW1jcefHfSP9zbvkYxawviIS11wRgy3wjqqYk6cxroRI%2BrEiFuiE0KN47mwQgm1SFzGSU6FIrHOC624csEkEca6uhQK5w2nLse6JcgFtpIaQRtkA7sipvKVWC85J7lWGv4NLSYEP8vvJJV6UJ5KhVZ2ERux9EyolKwlBg4XyVh7yciAk8LogeQ5Mo8bZHfraLu%2FeXgPBxIeOxwmY%2BEy%2FAQdI660t2Vw6xV1nNyhZIxPU7daeni5ltgVIXcJVYxw6wTCgLhAR0ENtQI%2BYsnxNQDLqExAR8Ib5IQTPqLSl%2Bb8jIzESBPrbcwLJwZCCjcJDF5z5BE2JZQYfNTBVcpxwuAkiKDW0kmZRQoQbnK9fHXDZRkJm9w2yMHW4fGTI7CQewcAYDAQiBJZ2FiRKpGAQ%2BXkZJEkRuek3ejdDuS3ouZyqE%2Bz11idLfQWCfOBZoA3Qk7Cfsj7ymNi%2BBeeq%2FgS0tpKd%2Flpme1GZ6X3jFApuURb3dn%2FNGpEUbPc2if3SfjVWQxIilAtNObkboMcggDJ68qDWu0EQ3AtJ7k2RSZsjg6TVA0DV1exZkBDzscp3KOw3AQCMQhAKpDChDCRJFhF95aoXaZtwEEOtYcrah1Zs2XX9vc2nhwf7vT3rnGYowRoONAw5KZqdMbdtM0q6PA3K4u7rCwqcuehVwxUDUAfkn5ECxrCxoKjEYKjsLqPpIC3cnaIvdBUgapyJmzjbkPSOOYWnG%2Bi3Gkraq3Uo%2BU81sphOLnZUYm%2BqL2ZUsDRxp5LkRcQgYf404a3Oo12jwtrPX%2BHO%2BEkP%2Ftwj4%2FJFtgOUR5pbxR6DcF3ORMxPCYjzEXOby08Ly6DPNExhbuL2oIo0KMHVKUcEoSWghrsY%2BXWG99kEmRW33%2BQ4aQRA49D9ltYMkXzcCJJaI5%2BOl2nUgxpCsnAtGNG%2BTp3bsKMlrz2T%2FPkoR96NjVN96ie%2BKGYZ8x3hnxmG%2B8KPtfpY%2Bq80ZWh6NsBN44cNeaZiyMaZ2JA1dR1fpgJ9cf5dztYNXreqewZzamHSXUo2aUWRM6zZv1CTBNKTqiESM0FuYWumZpmh3ToDR3OdZusqcmYBl0N1ptAa8naXKDJY5HD3TSNLWHhfS68fVQwRr9Nfe8J3ABKzeU87zOtUl1fw8U1mYbQW3lOleeS7KTIcj6KPrbRdzMijRNqLj3buBynnIsjbTPxb0Viu8JNk3nVOMHtuc%2BSF7Fg9qL2Vsy08M2o0YyWV5YUP801bTWjZrezwoQdqHP8t%2BpLEUWt1Xqn22uL5nK7Xe90opU4p%2BlpuxVB%2B6J2t5vhMsbc8bPhydUiK3LBsnZnebnZaq3CV5%2B%2BOC1nt5z9U8x%2Bsx716q0WZk2lHrM2QCMWuMWEhVpUUrBQojv7AK1m0bLeYrJsNd0Y9Kn%2BDKFYGP2YHxt5tpk5V9h7S0vj8bgRUDW0SZcAdalgydIM7t7Wo91LuA8C%2FZDqI62lve%2BM5ylkN9xRoHU428TPBUgb1%2BgpxV5e5i6IM2xlDLFk65MN7ZW7dfMnPCpKuY45O8G7xpbr73001a721ghXQ5BICNba1b1dP7gU4N3qGRIkvno2JE4XIrZfoXwJ0%2BFFg7ADwWinJEpvc0xaNtNeyxPBJatMBq1uaZPNtBFryVEzinrNiuT3Z9EQHy%2BCOKsuwCB%2B6B4bQ4H%2FevBnY%2Fv3Wq2WgaCXfLNWt92rvHT2QwVjUmWyCHEeoQFVeFqFZwA89uPYF%2BWNhIXK7L%2BD01rtrlaJ3N3PqMkxzR6FQqTPPWQal1UFCoXgJuECc5fOsP3482%2Fn%2F2dsnVZ3uUrkk200CFQh4JoNw7WqHV0%2BZ2fgvv%2F619eD60zL%2F%2FEBZUJLneJNtRdeLtRcXqOlzx1MPdhiYTDejj1m7fZs1jBe5Zi9Vk2G4U3%2FFPc%2FWFn4Gw%3D%3D) | ★★★★ | ★★ | ★★★ | **9/9** | High |
| **Conrad et al. **[[3]](https://dummy-citation.com/citation?d=z%3AzVfNbhvJEZYMJPE9l2yQIB0EBlaARHFIipIMBAtZkmXrh2JEyc4ee2ZqZlqc6R5095CevcRJkAfIKc%2BQR9A5LxDknFPOue4pl3zdM5Qorbl72cMaMG3OVFfVV1XfV80%2F3q09S5QWqZDP4rnS07u1n2Y8NFbzyNY%2Fef7q4PDs5OryZnT0klnFbEaMa0uFMEIKySJVlEqStEwlyy%2B2Qm4odq9DIbkVSrqjmpeCzKb3UihjmcBpbTmOFzznWnAW6yqFBRWkUzhAiImqYM9hfWBgwGXMpJrDhyZuSRp27N8lWkS8w8ZIQvCcaTLCwHNEm2yeiShDBCkSMtYwblhMOa9dgjlx7awYTyzpNsl6kwnDCooFIsSs1EJGouR5XrOwZkVlPSLjsnNQppTDf6mVJTw5C%2Frs83GCfzY67FwUwrnIlUyFrWIUI2cxt8ChEXPGRc7DnFhTnxa2TwcwTQmI8ZPKLiFz8Re4L46v31weTV6yDX%2BUI079FQJrAEUCkoxhFRykKHxOUYVqA6DNVGw67D3iGQMTHFg0Ai%2Fh2TzKHPgRkkSaWXaTIg6XLEYyMERZE60K1usGA9cbVaUZvvSCDrs6ntycXyO1V7V%2FsoWPXjMEADjjuQcMlCVaYVAuw%2BbCZgxvBEqFnJRmEaL5b8v4C66npA0e8ShzuSsU1WbIqtd94VINAufXRQqGS5nO0WViptIzEnnuXc0xFJGScRUhYIdd44jvIRsM97%2F0JR3u7rxb6v3cuyCSLk6CakpMMvzKtjLuMYoxBNpg1zvAECFRV%2BOl5roj5Gdvs4HhWsRb4vjZW65Q0HUk3Bm8YDzSCi0tKkweXjwJjpl3FVzEUEy5twiQutwbeAD2%2Bh7PfQn5k4j9PXd80JQTTL%2BvovtuHDXn4NRT2G2Tm0A7w%2BDNfaBN71%2BoymCYYjKRFmHD86t5mzk6kQgNTi%2Bq%2By1hHhXtcea9%2FgvH1oc8BoPg%2FLsBBz0H2B327Nb0ANkT%2FpN5IMIJSbIiapiHCcWbGMxsyB%2BB9Q%2FcRhSXBlTvYeJhTSXho5FSENN4fj9WvwbII09LdHCVU6EhjLUfnFgVYK20DW0hOHjfjD4lCUVWzByFGrpj9q1WOcswL6GrO0C7Z0JWDQGxEKSBBhlXPBepkggL7eqww8vR4fnN5O3lCCQ%2FcuLmpaCtkMnUHC4ey1vD9idK7Qq%2FpHS%2B3EWVW1FCIVNSKaQZUu7r6UnYjnlLLDcES710JGsGHkEWUjpzGi8KJP356wrVBtjapzbyHmH5FrCgd64nSPIN8dxmnY1OzqPICyTwUYqWD7e6O4WrEAdP9VuZqLu1H6Uc3VLafMyx2bBAX%2BMvNKk36PT3CbWr6DmGxOb04VcjmrNjmUJ9YnaqKu1iI%2BAFlk4Ej8lM5VVBn63%2FpbwPcu5wK323to5llNIVlylhfQvPljGefPbsX1mOWWn%2B%2F%2B%2FcN1WEFQ6Zv8IylrxwJ5KEFyKvk0MlNY9RoxnJ2wvKkSFnR50Yg0Br37SPD0zFG%2BvkHfig9ErLE45Ge8t04jhU81Wmtye4EoSaogx1dQfoiM9EvMo8GQmu%2F%2FH3xjQ7A38KNVWrrGmC9Zu1KU%2F4vKaVKYsRLypMbNoizE851O61vxyYVYfSM0y70KqNcJ2pgq80pnNKedRmfkrSXUdWppMe6irGJLfNuaKQooizi5XNEddVMaeQf7U4MuZOtKbsbOWRZGRUWLd4xQT4KWfnq80PlSqhHo35lfKje7DSnM54sXCeXUDODLta7fsII0p1OwEnqLpdafoKl6aF6e0ppExT%2FS15IFMDVc1QzCbzcSZyUbLT9kSScTOOk68jERtQOIqVqIJuJ%2BjuDLcl3RaK93pBsNftQxFD%2BRGfRv5JdLu9va3B7n5fBDv9%2FtZg0B1mqA5YTB%2Bm7wf9vWEQ7Ab9IC4LEWf9XXwNej0cvuRf33ol8EpyCyXpb3X3tnp9MFemFZgbkizKKgQbodWNsKx7TB9%2BeYFrGtZWZchiG3mtgGxMVCTI1lMoKYQkohudfzjKrC3Ny%2B3t%2BXzecTA6SqfbwLZdxsn2At%2Fo%2BPTiHt8XTpBxIb5WKje%2Ftbqi1FShrUtXzOniJb6uQylJJWfYWcuPybobImzzyN15X9WHqpL2Z%2F%2B0mjDoTpPj9%2FiBYZrHG60Qjo6d0vmVDPlbXN%2BvHi2FgydL4aJdWrC%2BX5%2FtukmsKnFt%2FfPd2o8T7D%2FoJhIKcXMc%2BBKqRtDZBBVzxKZEUB43JmFv19tkCw3Gs%2BQ66Hb3g6b8v1jERXb42YCbl2MARNRtTlxloPT%2F%2B%2BK%2FX56t4U%2BG0i35jnu7%2Ff3Gy2Dsehu1q2UTS2AGBZEFZhTAncfLKKrKxT5qzL4%2FOL293b0mkY1xxnUBRUL13Rj9rsKF29YtKPSIdEICN4B0ge1vv%2F8P%2B2Fg6w%2B9jW5txlrhp527mpgG5WAw7Dcp%2FeYIpIe2NH4slmX00L%2BJ%2B2GDLy2%2BP%2Fx66%2BMn8PWH3W7j7OfulqfvM1%2BOGzuiPI8qcO%2FFgnugm6fdJ%2BVk6n5tv0NqOL3%2Bfw%3D%3D) | ★★★★ | ★★ | ★★★ | **9/9** | High |
| **Oyebola et al. **[[4]](https://dummy-citation.com/citation?d=z%3AzVe7UiTJFYWNWMX4MvSINdKZEKymG7pp6GYcBTTMzmiA7gVGG2tmV2ZV5ZBVWZsPmpKc3ZAM%2FYxCNr8iSz8gcy05OjezeA2gWEOGjGG6svK%2Bzj33UT9cr3yWG6sKVX8mlsZeXK%2F8vOQL5y3PfPtC7e9N3391OvtwcvCanZeSLYIVsmYmZxXX3CrOGmmdct4xVTMXFr0zXnLLa7aXW5VxxmvBPARlJW0h60ySLLdeVsqpGjJWkjinNyUnLd4aETIpWGaqRssr5VvmDZ7ojWYyh7%2Fe9dmxqZWH63WRDOQ57GXtJ%2Fp7C%2B6gy1vJfSVrzyB967tyLLMhU1yTBS4EnHHQRucl11rWhewjbjw7H0TLuHO48UDfI7sS3tieDpXMOVxWtWRre0frEQkgIL3KmFCXBBsig9Rcc1cZoULFcq4z1XCLn8oZzb2MuHJ2ogoJj2vWmCbgXJkaCByev50dnL1mcxhWJFgjDw2CgGOEy1L5kmUaOGQI0bVV403lyGaoI7o494jmBg%2FuYaqUXJMYdFhJ1o94YdyrGxdesaXEucusRDAi4jnv3%2FO8zw5rJErjHZ7v%2FIpiETe8iZ7tHUVQqpRIQrW0JhQllJdSBNKAH9lFLzTsEun08OI2GpLUfGEsh2zL5BWvVB2BcdGn4YQJ3oInAIdDVkJUEjNBtKdTER28y3DnS2N0C%2B6CRcSJSFiUBsywtfn0lNLKdQsKx5qQ1vzRkC0XbM5hqbHGSwWX1irXDFK4rhmuE6vorbzkYFmqCmFDcb8cquC7eJYojFvHFu1DvHFPe%2FWpMIXHBjAr7GCdnoxvG%2BJEjtSY5VN6wkJ9FxSI03ONzBRondyn0KEpLJqkCdHJ7wK8hro%2BOz08%2B3B0DhJSQECkR9SyVmaU5rsyMcGDcRIuXoJfeLU76o9eouagCox4mFf4Qykz2hTxDHE1wIEKaTqPmAu23d95CQJA9NPbOVc6IJVrR%2FM36wz8kGDBXu4liDo9RSOJ3gHYV6w2yBRFS87iOssoWIJbSJ9OyRZZZTvjjZ0xWxtsbr5cTxnJSoVoRKxQjRYCAFQNPikRuI4dykoDdr2iqlLEpQf5XiotesgKSIdq0Sj12JuQMHYy2fmWre1u9ccv1yFNrKHjg8FwRC8m4%2F5254RZOGnhRJ999YjQt9S8BR0OCEN1goy8Bx3JLkRy1I5uk3mi6druZn8IA2T4zXQ4vnkzZGs7lDbQ99B5VaXmQfSjVtK1M1XnCV62djx7l7wc9MfoG1TzBBJhIZ1neH3rP4FI77Z7g23WolCBbwEaowibPqgJrzrWUfl2cSnMIg%2BiQtyAljU40uJPW5ng2OFgezg5SPVrbIMmXqUGB%2Fqz3CZlLRIKIiGg6exkevTh7N3sJFE5NXwh0Z1oGlKkoIpHB8Cv%2B00fXYzgiVSnAnvYWj8p1dRof1qyLnGVArnJElEFHdCFogB6ZMpkWQCZgUlsPxRqpg0U3SUBNDwBNoAWBIM0YZy4TRmimcWcKmqqdiJFBPoBZJkJWmA0X0qWB091pbroblttHL8NklEpKsI0CpGYXheuftTaumTfG2aaZ4gC%2Fe0AsBXDzeFOb3O7oqFPiNt3dW6uVz4vePClse57DSfQht%2Fgn7Fiqz%2FYksq5IFeBqtfSHHcT7fcmWOCRXxoayL9obhUeGURg7PXKKlJTyFOOUY9dCDhY5%2Bc4%2BdXq5yUG881vHfcPtQgQcn%2FFTVHziiTyVDzFrJULTGxZIKN1%2Fh4%2FGyMwB%2BXK48v5kSq4VumufhNqHPLWsGn%2FOYlitgS0rUwixR66OF%2Bg6T99W%2ByhxDvtMx2WvFggNwV%2F7n62p02n%2BVvpQy3kczfLQyw0y%2BA638t9YhWo%2BLzfOuSw3Wk%2FVpihUj%2BLyxnSdtG5Yvb5gkdnsIEseSeUY0mci%2FzHTAkHQmTCqKtfDzb7g8FkZ8MNhrtbIM5wBPJsTia9HaHcov4ef139gxqMxtu94WS8XZpGghXy6uKb0WhzuDkYDMZboqmUKLd2h4PNrZ0xRGb8x4%2BRVJGTH8FJqJ30hrvgRV0E8GIh66oJC60cVr5E0dUYUr2vzDFKeEpLFNcXoD6ImMkPVl%2FNSu8b93pjo6uOj4mi%2FYXCfBRx7cITesgGSq%2FG1NpoRL7xfIgFtm5qDYDxIu5bWp7jcRVVJU3%2BHhPp%2FrH0aCct7qInoT%2Ftt1My8gLdK8f6g9oU3%2BArwMXTX36Rqunqb3tx%2BbhZtWnB5Y0M1MB%2ByvpLPeLphfPRdvka%2FYguWlnK2oEGXR9Gr7rbJW7XAZpP9xcmbi%2FQTXNvGpW5P1%2Bv%2FCzHtEO5I1wUCx%2FF3Ji3acE9y1TsnBLzQ4t0ZTEcxzslkoeBVkuc5ecY%2BbuDeH71xU1jOYUgt1kZfZimbxPpsFvIf%2F%2FuX%2FK3%2F1xZWSmRmHu6xXC8tZu0jOZEmowlT15hYb5U1tQUXLcBzdDZm9gZcZCu%2Fe%2FCQQlMkiPrc3ysVVhUKZew9DUWlzSMKKjIvlzSSoiJk2L7y99nCO3%2FN7bRcLydHPnyLXiB70OKK97B%2Fw%2Bydn5DKHA7Bfenf3zNngxu1KX%2FN6dcpBkHxob4QZF0E81J57uK41O6EFSQLzCc9dWXN%2BWOTtU3tvgvlXxBH%2BB%2FoH3A1Kv%2FAQ%3D%3D) | ★★★★ | ★ | ★★ | **7/9** | High |
| **Dimbu et al. **[[5]](https://dummy-citation.com/citation?d=z%3AzVY9byNFGM5FAtEiUVBQDAIkDp0d2%2FE5Ng1ynOQ%2BYt%2BZ8%2BlOQjSzM7O7bzI745uP5Jzq0FFTUiM6iquoUkfwd2igQkI8u2uSFFkqCiTbWs%2B%2BX8%2Fzfs035xubqXWUkdmUp9Ydn2%2B8m%2FPEB8dFWG1u7pIyhrhmIVeOL1UMJJhKUxJcrFhhDQVom4yRZ5wJF0UpDF06obBiMM2U8bESgYkrVZsyEZ1TJugVi15Jxl1QBXkyZFoJL0%2BELRIyPJA1a%2F8rRoaNTWY1b7NJTlrCAjulkMNnDIpFA6Wlho8AA3PNfWElxYKlXAtacodHMqlCgDAKYx4RkFGaeQrKlydlmLvKZFFpfod9xcmpO4wbyabRSM4WUbO5sydkBORPlVMsOFW5q%2BMoYagy3JaOhUq5CUCv2Kfj6W0GOsr3PgKVavEyNv4i1q8X4y9vV37WrMJgSV9vyCRfeRYs494r75nQ4EiUNEMYkDhCt9pm1ZlTfmkN5NpsZrUSUXMHIsF0DfmUe7ZUDpYLOIigOwMHJ2CxIOGsR1xagwpWcHesnG9rLgAU6djDq6zX6Q1anbuFsCZwxO0emNSeb7yV8Rhy6%2FwrTcUS6T%2FA1zq53e72FXkf1aYKFLR6%2BckYfFeukrJSxhn4R%2BkAySRXhV2nOT2xJXnvj3XpyFESYc1%2Fj2KVhheqLNqUF6RXao%2BKJKqMgEDPlXSWPeEpVygMoN%2B4QeG%2BdXBSKWRPbKJc4E2y%2BeTiRyNJ2lr8xdhwFMHFG8%2FZjAQob1Q8UCCcHK8V7YRDEJUMtbKsmtSyCYpBCVVr0YHS5AuoNcmn97hDv9Xi6TSecd0sOzbSXfxUy%2BaH4B6cN0pnc1UkSq8jsaWfizecPeJnHEYaYT%2BETYvP2sueRfSZ9Y252L0SLnaVLsAYe2h9s4esRMHlOi6DNj3CuWGPSIXGbKRTnkS5pilfoGFyNms3BjVVFz%2BbWnh5jyeOMBzmFt3OZFlfN7s4RNeoVa2lHkZNjdSqedli6xTv5S6eUKKAqsHw4oSyS%2FEFLzjG5aIxeHrKNabUGYl1KPtaUrPxaRQipzUxjzjSxZ432k5n1nHy%2F2C0%2FF8K8wBNptai%2BQFpW6CKGpM6w0wkzKy6i6sCYAeOY5Q3qRzNNTbNL9%2F643X0RzOwGv3ZZWLTnPu5TH8XJP35xttCWgrdTrvb7Q23OBftTnfUH7V6HUk%2BMa%2Fw681r6nQGg1Z%2F2OlTt7PTaQ16w208jYatwd3RQBQ8O9rujHaG3WF%2FtJPbpTIcE%2B34%2BdWhXBYk8%2B3t7mA46PRh9TH%2F46iaf9X4PML47LS63Vann6P%2FssgzheQXy5ig1TH66ml6q0L78sNxoRyGumELK1Dg9TrF5ClnZznwV5mPSVgtS2KOSw6FVk%2Fx9xYGvbLpITJ%2F%2FVgFrvUKstiEWFe7q4mNJrz%2FAOsrxRpDhcvnuAH46vi99byeTDCAyQAAm9pTtn9tf4%2BvNt30%2Bqa7XNHlUw9Ep8EuSfjXyEMqbYG9gSASkrxfAbX3FdfYnAuALJeqStFyshZJejuVTD5TkgSs4yx92u10Rt2apA9mmGSOOHsCRYwpLOByl5RbA2XgsfnUn1%2F89tfXP2xsbOSg65pt2dvZHtVW%2BvMyA4LVkdxh%2B%2BaEnDWoWnBWWXwsRFxWVxEc1GL%2FIZzuYFAH8tHMelwIgm0l1oFLlJDCXajekNUuvIYqfsf%2Bz6hQ7HfrQD67r0ovAZfEZ%2BRw2%2FNsEaKkNa79JUlc%2FaqSvgR3%2BOvNKevViT%2BqTFYqsmyBd3Cb1C8%2FzkNY%2Bs%2B3ttDubeuyrZta%2Fri85z7D3Qawb%2F0N) | ★★★ | ★ | ★★ | **6/9** | Moderate |
| **Kiaco et al. **[[6]](https://dummy-citation.com/citation?d=z%3ArVfNbmNJFU5GGmDFAjaMhERtRiRgO762Y8fdQijtZKYznT%2BS9LRmWb5V97o6datuV9VN4l23EBK8BbwAEgsWWSDxt%2BQpeAB2IyGx4TtVtpNMd3YjdSd21anz%2B53vnLy7XfuosE6Vynwkrq27vF374YxPfXA8D%2FPv6me7kxefn528PN57wvZcUzInvfKBm1wyZdip5r6yQjUVK7jOVc0dPs64Z7X1UjBumIUynmvJgmWyKGQe1BW%2BOMlDJU2AiGD5jGstTSk9q7iZ44fmTnGWWxOc1ax2tnS8qnAPo9IIWamccejwHXZg2K4preYt1pigNOt1u%2F0WdGrr7JtGGck2Jr%2FaZNfwqolOeVYo50Nb012YScfrOUMW8D63Va1VzgPkFl7AQoiPnaw1z3GhjAoKHs%2FZdM44xc%2BTHcTSIg%2FhwbAVL12Aqx4PTHvKyTgMTJXhQVmzMr2xO7mAfyrM0gNJF23dVLLgSECMYPewxSY2Xm%2F8%2FY%2Bbmx22XxRwNJ8zHxoxZ7ZgUIO4rRFNTv7DkZQYNpXhWsro1yAmHB%2B2W8zP7DXlg5FrMRkoUNIJbf%2F%2B3W%2FH40877Gj%2F4vnJ3vkTlnX7q8rUiADV8%2BxaOjwzqJImVXlunVCmpGq%2Fen5ClculpBNU6vTBowJPov2mTrHncAHWdfQQSOJeBattGc%2FkFddNTBuqh1L1dpjgc99aCkompMFv5ByvFb4ERbHERFP1FgpgD3UBfJz1ua3nLaoCq4vK173kVymNDfM6CRrp8aR9OjmDoGUC2EcsjfKzO8NO5q4R0ueSuqJwtsKza6Q%2Fgh0OP002rJ5X1tUz5SvPeEClBEWzM4wuZ73BkNIOV4TLyAvZihej4Tid81AP43nyk3sv8S%2B6Cf%2FaZ58dnsYHXr5p4ApVgTJVF5dZv41C1BI1cqv4Ouxs%2F%2Fzl4QUqewH38gY6HRIUszXOOn3Uni7sNHBgQ1DrNxrVW8AmzBBDjJaAsQREC4F0hp%2ByDcN%2BwbL%2BJtCptAYlrErK0RGcZVQ%2BxosAjyg7d4RA5gsgwc%2Bk6LATEwW7LcoBCY4H90rpOdpVwui10qJNUTFiEhylHC1xkLph1O8MFp4Nx5uL5MRsH%2B8MvyLKQl8DLpREa9DeFrlOJljOnSM15ELVgADBXNES2%2FjqgSqq41OWwVBy2C9VrBKnTE7MhU%2BLJ4RDZppqKh1edrsLH3vZZszbN2MjxShpggLCwml0i3iAXFzGgY7bOzifvDw%2FPzg5jkWGL98oIpg30i7UrPAMqBQKZnwDSNdBTZWmvgL8dw9JkDgSJOCJwWy0vOIPwQNPmADD9Oh6RTmJ75boI3DCNk0ZCVVB5YsuWZGAbQIMIeYEkTt4EFXAEaEEgyoG%2BUtQGDyCkgbstAJ4Er17WXClAfKnNJKku7oXvbBRFQK%2FAmIQONEgwRlQIwhRW6PC%2FJ4ydLMCTyYN4PomkPsg46aERvqC7sAEw5AqGhoVlcXUwKRF4MqDIKRIoAzggzjKOhgueezoPWC77HV7w3Z3u6IZSP3nDkxhb9c%2BLnkTZtb5t1qhDHn4DP%2FBuf1O1pfK%2B0auy6CClvZoQdVf2MYZrosrSxPl%2B%2FVK4aEFPVp3u7aOuV3KM4oE6wDaBcPxFCefrP93hgG%2F%2FKzjOFYIFeZ%2FA0lheEUvioJXSs%2FlC8VzK0vMd1O%2BwJAzJRcYDHLtfdnZhVQ3UjmexOUXlptHhcsjjjyLhWp5hFI%2FrvjM%2Bn%2F8wamFsPlSufKff%2FrXO63A3vaxV%2FIQ7OjTk2JPGesWBgrsMqei%2BDpXwt%2BufScXVt18knU7WbYz3PJZb9xHkbLtdtbNdtp9jIepeYuf3rxT2WC03e7tjLbzipevez2IDcaj%2Fs4MplAPeXP56u5Q1JUSs95wOOoOBz1oOOFfv47ljGh4TYLtrAdbqAgGECoylaaqm6kmolyAYz2GZZ4pewRwTYBTx%2FUlWh4QyOVLp2%2BOZyHU%2FsnW1mKQv07g6EwVek1ggtGLDhpvK7fYpqTbqkWx9WjApW%2Bm1GrI5GVsQC0v8HUd7CNtAQyI%2B8cy0NIEWWyKYO9n8wnZ%2BNHP0VUF2BptJl5hBfXpuJVwfPPn%2FdXoJ%2BZ7bENarS6xp%2B4PFDz64GKXNgVwISapzdViVSDOSLyM3YJmLrgtTegXD8bow3EOo4cNhLCEpp2rCLYGqf0aoCmErdBwiHuqBB%2FEGtnnkmtYOieaQddLzCgtksi0N4oyMxQRNGQkzooLzIVxFs9vfrxs7TM85C7H2gj3JmlZxh5infzfL%2F8Tzt%2Bura3NUKF7ukVv1B8nLYNTAk%2FOkicttm%2BulLOGUrbYwU7yvKG5boGQhdh74SSXqkNVyA8Hk%2FWTuRZQmc9oGw4OW9fnaGRi%2FWjoCG4n7oYQNr55iniQ9bfT459M6C8OMDCEAdWoh4YEoB0TsAw6%2FODjv34g6Kyf9aKiN%2B9Z%2BvbqM%2BiNFt7%2B7DmCBNWTh1EGvx%2BU6mKJTWxwyfHyb3%2F5%2FQerNVjU%2FKdnXCSPW%2By4QUMhhKX9qPMALIPhIqgdv4dlTt9sLnsdpNWxrny8jS%2FpL78vpfMo9fr%2FAQ%3D%3D) | ★★★ | ★ | ★★ | **6/9** | Moderate |

**Legend:** Quality scoring for the six core prospective cohorts included in the quantitative synthesis (N=888) using the Newcastle–Ottawa Scale [[7]](https://dummy-citation.com/citation?d=z%3AxVbNbhxFEF4jJQIJceECEhJ9IIotZdczu%2BvNJoAgWQdi%2BS%2F4hxyiSOmZrpnpbO%2F0pLvH680pEU%2FAEU55DT8HEhcOSEg8Qk5c%2BHpmZ70hNickJMva6a6uqq%2Bqvq%2F75VnrnUQbmcr8HTHVZnzW%2BjDjkXWGx2525fe7d0bb3x7sH%2B9t3mY7PB4znTBeFEYXRnJHzFChjZN56jcm5DIttNKpjLlighyXyrKMW1YYOpG6tGrGIqKc2UxPc%2BY0E9I6eGBG2sp5JGHNrSVrJ5Q7y2TODM%2BFnsjnJFisc2e0UvjpkIGyHXaUEbN8Qmwi08zBPXZKYkDFdGTJnHAndY58rCuFpPmJVGMF8VwmbbUzY1NERkaxnhTcwAus9mgac%2BsUtfed41PODgGM2Ore%2FuHaUpZVsDcQROSmHqeHTVMywJQLxksUCL9hFWv8ck1OQBmrUgAV4HJWlJGSNsOnnVlHEyCI566YhlMfKUGDvC8f2tIJIWNdOiRfeWMFzszrl1Ds4GoqXea%2FVEn5c95hu%2FeO7u9vHt5mo8tT8TV4OwUP5TzFBmo3CIbtbhCGDHhp4aYud6yNIVvoXFSzYlgijXWLgviaK%2FJZcmZLc0IzLAGTN%2BZKMdSbSWSB5h2QLRVwVUHmvZqDqzoGy6XGYFaV9EnOllpRtf0CYB22lTsybYPJRj9JSR5JJV09Guh8XCpsCVZan9gYzjlb3V5D6eDFwpHP797h8c4RynrnvNmDjfVw2GWrvcE1b1zX%2BRxzlUyFui6WR96gtqgcVfGtTHOZgFm5A4syDDuSXC3YFyzoBEG41vRrgfP6ciFWJyQkz9mXbPA5LAHwWcnBXEwz6JUSe7T13cFjNmgP1t6qqrbkC9g069zVxg2GU6yPQ5fXDid96%2BowgiVGT5hVFVdXt%2BEk6ITwc2vjmid3IgXlsZ8euDkBRR%2BNth7DqA2rWzdg2x%2BueZIWGjNUHcdOMKjPj7bq727XW4YBkjpYSsQzxReoKe95ad%2F0FvTfdBeGlbsQ7kb7e6Od48Ot%2FT00eFMmCYYQ6VaMW546EERpECU1RNXKv%2BuBLdOUQAefXU5%2BIrwM5Q4UX9j47MFebfzEQgRy7ZZIiPjNXE0zxPFzP%2FND2nDCG%2Fxz3m1H8RjZWxKbaFvaDbqDdrAxqULLnMwW4p21rqTzHF4oicGI3TeV9Ihe52aPpLUlrZCTEMlTdnd3xHYxHV7%2BQVTiJs6w0FwLs%2BREq3JCHxSLEDs65nB21lqRBU%2FpwA8J7iQMOvThAVY%2BamcKElz%2FVJX%2By6j00vcD7EQO5fd3WMInUs2iHU2pPKFcjbixqNJ2h%2B10BG4Mar1tTbtk3PP6QLqJGyaX48tsBTxHtanY5WZul%2BBueyCS17EU9qx1NRZaTkM%2FK8PBeti%2FGba73eGwHfbb%2FQ1cdFH%2BAv9t%2FlIu9uIJT592w8Eg6G4Me0GmC8JVRafjh%2BeLophIkXX7g1tQ2C487PPXT6u6V217Cs3ttwP8hahUnpaoVET5pBmOeRdXKkD5XanRIDbCUIIEY4iyLk1Mx0ad7mfOFfb2%2Bno0iUFxbDUXeieSuFdEXB%2FqQB%2FWY116iq4XIlm%2FBHFqy8jNCl%2FEsZeaWNERPlcwvaSTbZmL5WU8FpSawVbFkAtxdzbyET5%2B70dnaE4z8RAPFFutvz%2BfuPsX39C35xrmGbCg3B8vfvLEmg%2Bz%2F1p6ZyROF9BvzNTVBI8NzCZSiaTg3apu%2BlDHeG7AO25VUIYSSUrUJlE4rGyebVIsredmY4XN5CgMusGgMjj9DFTgbY4OzyyeHV4C3iYlVbr011e%2F9X991Wq1MlRxKZgIh0G%2F9nb9cHHt3GAPjI4aofN%2BjxHfeIo51PQ%2Fh9TrBbfqJFaXFdYHvrPQPEjOLnFcbNVnA%2BuXn8dP%2FjdYvd6FNjWobr9fb59%2BskmqyCQ7ojjL5TO8JoGl0bMGyKs%2Fn3x9AZBeL6wTOf3UR6lkr0r9gQaKShnnYYUnx7txCepdb6gHAelok15GqbF%2Fon%2BPWUZLVv4G).

**Commentary:** All included studies were assessed for potential bias in patient selection and outcome reporting. Studies scoring were classified as high quality. The two cohorts scoring 6/9 (Dimbu et al. [[5]](https://dummy-citation.com/citation?d=z%3AzVY9byNFGM5FAtEiUVBQDAIkDp0d2%2FE5Ng1ynOQ%2BYt%2BZ8%2BlOQjSzM7O7bzI745uP5Jzq0FFTUiM6iquoUkfwd2igQkI8u2uSFFkqCiTbWs%2B%2BX8%2Fzfs035xubqXWUkdmUp9Ydn2%2B8m%2FPEB8dFWG1u7pIyhrhmIVeOL1UMJJhKUxJcrFhhDQVom4yRZ5wJF0UpDF06obBiMM2U8bESgYkrVZsyEZ1TJugVi15Jxl1QBXkyZFoJL0%2BELRIyPJA1a%2F8rRoaNTWY1b7NJTlrCAjulkMNnDIpFA6Wlho8AA3PNfWElxYKlXAtacodHMqlCgDAKYx4RkFGaeQrKlydlmLvKZFFpfod9xcmpO4wbyabRSM4WUbO5sydkBORPlVMsOFW5q%2BMoYagy3JaOhUq5CUCv2Kfj6W0GOsr3PgKVavEyNv4i1q8X4y9vV37WrMJgSV9vyCRfeRYs494r75nQ4EiUNEMYkDhCt9pm1ZlTfmkN5NpsZrUSUXMHIsF0DfmUe7ZUDpYLOIigOwMHJ2CxIOGsR1xagwpWcHesnG9rLgAU6djDq6zX6Q1anbuFsCZwxO0emNSeb7yV8Rhy6%2FwrTcUS6T%2FA1zq53e72FXkf1aYKFLR6%2BckYfFeukrJSxhn4R%2BkAySRXhV2nOT2xJXnvj3XpyFESYc1%2Fj2KVhheqLNqUF6RXao%2BKJKqMgEDPlXSWPeEpVygMoN%2B4QeG%2BdXBSKWRPbKJc4E2y%2BeTiRyNJ2lr8xdhwFMHFG8%2FZjAQob1Q8UCCcHK8V7YRDEJUMtbKsmtSyCYpBCVVr0YHS5AuoNcmn97hDv9Xi6TSecd0sOzbSXfxUy%2BaH4B6cN0pnc1UkSq8jsaWfizecPeJnHEYaYT%2BETYvP2sueRfSZ9Y252L0SLnaVLsAYe2h9s4esRMHlOi6DNj3CuWGPSIXGbKRTnkS5pilfoGFyNms3BjVVFz%2BbWnh5jyeOMBzmFt3OZFlfN7s4RNeoVa2lHkZNjdSqedli6xTv5S6eUKKAqsHw4oSyS%2FEFLzjG5aIxeHrKNabUGYl1KPtaUrPxaRQipzUxjzjSxZ432k5n1nHy%2F2C0%2FF8K8wBNptai%2BQFpW6CKGpM6w0wkzKy6i6sCYAeOY5Q3qRzNNTbNL9%2F643X0RzOwGv3ZZWLTnPu5TH8XJP35xttCWgrdTrvb7Q23OBftTnfUH7V6HUk%2BMa%2Fw681r6nQGg1Z%2F2OlTt7PTaQ16w208jYatwd3RQBQ8O9rujHaG3WF%2FtJPbpTIcE%2B34%2BdWhXBYk8%2B3t7mA46PRh9TH%2F46iaf9X4PML47LS63Vann6P%2FssgzheQXy5ig1TH66ml6q0L78sNxoRyGumELK1Dg9TrF5ClnZznwV5mPSVgtS2KOSw6FVk%2Fx9xYGvbLpITJ%2F%2FVgFrvUKstiEWFe7q4mNJrz%2FAOsrxRpDhcvnuAH46vi99byeTDCAyQAAm9pTtn9tf4%2BvNt30%2Bqa7XNHlUw9Ep8EuSfjXyEMqbYG9gSASkrxfAbX3FdfYnAuALJeqStFyshZJejuVTD5TkgSs4yx92u10Rt2apA9mmGSOOHsCRYwpLOByl5RbA2XgsfnUn1%2F89tfXP2xsbOSg65pt2dvZHtVW%2BvMyA4LVkdxh%2B%2BaEnDWoWnBWWXwsRFxWVxEc1GL%2FIZzuYFAH8tHMelwIgm0l1oFLlJDCXajekNUuvIYqfsf%2Bz6hQ7HfrQD67r0ovAZfEZ%2BRw2%2FNsEaKkNa79JUlc%2FaqSvgR3%2BOvNKevViT%2BqTFYqsmyBd3Cb1C8%2FzkNY%2Bs%2B3ttDubeuyrZta%2Fri85z7D3Qawb%2F0N) and Kiaco et al. [[6]](https://dummy-citation.com/citation?d=z%3ArVfNbmNJFU5GGmDFAjaMhERtRiRgO762Y8fdQijtZKYznT%2BS9LRmWb5V97o6datuV9VN4l23EBK8BbwAEgsWWSDxt%2BQpeAB2IyGx4TtVtpNMd3YjdSd21anz%2B53vnLy7XfuosE6Vynwkrq27vF374YxPfXA8D%2FPv6me7kxefn528PN57wvZcUzInvfKBm1wyZdip5r6yQjUVK7jOVc0dPs64Z7X1UjBumIUynmvJgmWyKGQe1BW%2BOMlDJU2AiGD5jGstTSk9q7iZ44fmTnGWWxOc1ax2tnS8qnAPo9IIWamccejwHXZg2K4preYt1pigNOt1u%2F0WdGrr7JtGGck2Jr%2FaZNfwqolOeVYo50Nb012YScfrOUMW8D63Va1VzgPkFl7AQoiPnaw1z3GhjAoKHs%2FZdM44xc%2BTHcTSIg%2FhwbAVL12Aqx4PTHvKyTgMTJXhQVmzMr2xO7mAfyrM0gNJF23dVLLgSECMYPewxSY2Xm%2F8%2FY%2Bbmx22XxRwNJ8zHxoxZ7ZgUIO4rRFNTv7DkZQYNpXhWsro1yAmHB%2B2W8zP7DXlg5FrMRkoUNIJbf%2F%2B3W%2FH40877Gj%2F4vnJ3vkTlnX7q8rUiADV8%2BxaOjwzqJImVXlunVCmpGq%2Fen5ClculpBNU6vTBowJPov2mTrHncAHWdfQQSOJeBattGc%2FkFddNTBuqh1L1dpjgc99aCkompMFv5ByvFb4ERbHERFP1FgpgD3UBfJz1ua3nLaoCq4vK173kVymNDfM6CRrp8aR9OjmDoGUC2EcsjfKzO8NO5q4R0ueSuqJwtsKza6Q%2Fgh0OP002rJ5X1tUz5SvPeEClBEWzM4wuZ73BkNIOV4TLyAvZihej4Tid81AP43nyk3sv8S%2B6Cf%2FaZ58dnsYHXr5p4ApVgTJVF5dZv41C1BI1cqv4Ouxs%2F%2Fzl4QUqewH38gY6HRIUszXOOn3Uni7sNHBgQ1DrNxrVW8AmzBBDjJaAsQREC4F0hp%2ByDcN%2BwbL%2BJtCptAYlrErK0RGcZVQ%2BxosAjyg7d4RA5gsgwc%2Bk6LATEwW7LcoBCY4H90rpOdpVwui10qJNUTFiEhylHC1xkLph1O8MFp4Nx5uL5MRsH%2B8MvyLKQl8DLpREa9DeFrlOJljOnSM15ELVgADBXNES2%2FjqgSqq41OWwVBy2C9VrBKnTE7MhU%2BLJ4RDZppqKh1edrsLH3vZZszbN2MjxShpggLCwml0i3iAXFzGgY7bOzifvDw%2FPzg5jkWGL98oIpg30i7UrPAMqBQKZnwDSNdBTZWmvgL8dw9JkDgSJOCJwWy0vOIPwQNPmADD9Oh6RTmJ75boI3DCNk0ZCVVB5YsuWZGAbQIMIeYEkTt4EFXAEaEEgyoG%2BUtQGDyCkgbstAJ4Er17WXClAfKnNJKku7oXvbBRFQK%2FAmIQONEgwRlQIwhRW6PC%2FJ4ydLMCTyYN4PomkPsg46aERvqC7sAEw5AqGhoVlcXUwKRF4MqDIKRIoAzggzjKOhgueezoPWC77HV7w3Z3u6IZSP3nDkxhb9c%2BLnkTZtb5t1qhDHn4DP%2FBuf1O1pfK%2B0auy6CClvZoQdVf2MYZrosrSxPl%2B%2FVK4aEFPVp3u7aOuV3KM4oE6wDaBcPxFCefrP93hgG%2F%2FKzjOFYIFeZ%2FA0lheEUvioJXSs%2FlC8VzK0vMd1O%2BwJAzJRcYDHLtfdnZhVQ3UjmexOUXlptHhcsjjjyLhWp5hFI%2FrvjM%2Bn%2F8wamFsPlSufKff%2FrXO63A3vaxV%2FIQ7OjTk2JPGesWBgrsMqei%2BDpXwt%2BufScXVt18knU7WbYz3PJZb9xHkbLtdtbNdtp9jIepeYuf3rxT2WC03e7tjLbzipevez2IDcaj%2Fs4MplAPeXP56u5Q1JUSs95wOOoOBz1oOOFfv47ljGh4TYLtrAdbqAgGECoylaaqm6kmolyAYz2GZZ4pewRwTYBTx%2FUlWh4QyOVLp2%2BOZyHU%2FsnW1mKQv07g6EwVek1ggtGLDhpvK7fYpqTbqkWx9WjApW%2Bm1GrI5GVsQC0v8HUd7CNtAQyI%2B8cy0NIEWWyKYO9n8wnZ%2BNHP0VUF2BptJl5hBfXpuJVwfPPn%2FdXoJ%2BZ7bENarS6xp%2B4PFDz64GKXNgVwISapzdViVSDOSLyM3YJmLrgtTegXD8bow3EOo4cNhLCEpp2rCLYGqf0aoCmErdBwiHuqBB%2FEGtnnkmtYOieaQddLzCgtksi0N4oyMxQRNGQkzooLzIVxFs9vfrxs7TM85C7H2gj3JmlZxh5infzfL%2F8Tzt%2Bura3NUKF7ukVv1B8nLYNTAk%2FOkicttm%2BulLOGUrbYwU7yvKG5boGQhdh74SSXqkNVyA8Hk%2FWTuRZQmc9oGw4OW9fnaGRi%2FWjoCG4n7oYQNr55iniQ9bfT459M6C8OMDCEAdWoh4YEoB0TsAw6%2FODjv34g6Kyf9aKiN%2B9Z%2BvbqM%2BiNFt7%2B7DmCBNWTh1EGvx%2BU6mKJTWxwyfHyb3%2F5%2FQerNVjU%2FKdnXCSPW%2By4QUMhhKX9qPMALIPhIqgdv4dlTt9sLnsdpNWxrny8jS%2FpL78vpfMo9fr%2FAQ%3D%3D)) were retained due to the high technical rigor of their molecular sequencing protocols and their critical contribution to the longitudinal data for Central Africa [[7,8]](https://dummy-citation.com/citation?d=z%3A5VnPbyNXHc9WakUlxAE4bCskngRlEyl2ZmzHmyyg4jjpbrpJnNpeVqhU9HnmjedtxvNm580k8Z6WcuMERzj1xpljzvAfIHHpAakSEhfgQg8ICfH5vjdjT2J7EaJSK5BWK8%2FM933f9%2Ff38%2F3mg6u1lwKVyrGMX%2FIvVHp2tfaVkI90lnIvm778h71O9%2BH9fu%2FRyf49dsS9M6YCxpMkVUkqeSZYKhKVZjIe04eJyELlq0iNpccj5ouMy0izkGuWpOJcqlxHUzYSImY6VBcxyxTzpc7AgaVSG%2BYjCWqutdB6IuJMMxmzlMe%2BmshnwmeeirNURRF%2BZpAg0nU2DAXTfCLYRI7DDOzxJRcMWjE10iI955lUMeTRWe5LUZwYK7zBfVkotfkyZRe4GRJ5apLwFFxAdSIuPK6zSNR6WcYvOBtAMcHWT3qDjYqU5rJrGoxEdkF6ktriQqTQKfYZz2Eg%2FAaVp%2FArK2WCll6U%2B9AK6nKW5KNI6hCPeqozMYEGXsGKKTClmwI4iHjR1VqcC0is8gzCG24swZnCfoHwMrC6kFlIT1Eu4me8zo4Phg96%2B4N7rLtaFLLBogikylzEUtWG4%2BzUGo7rMugrZmysuT2VpkInKvZNrKQskKnOZgYhm0eCpORM5%2Bm5mOIVdCJiHkUM9mYSUsB5faHzCHqZSwpfFcoZj4Gy4hjEaiRJyGnFFcbtSxSrs8M4E2ktRWTDnyKSfCQjmdnQgOe9PMInn%2BWaBDsDc87WH27AdOCiwYjkOxg8OhrCrJ25s9vbW%2B5Og603228QsbXzXGcjjNHaGos0L7XWsJww92s5jmWAzIozZFGIYIeQ6wn7DnPqjuNulP6a6Xmnaoj1ifAlj9l3WfvboISCT3OOzEU0I73Ggr17%2BE7%2FPdautTcWrKq0IAOWzpqz2t5kOMVaOLTadjhJrrPX%2BCxI1YTpyOTq%2BkMwceou%2BOxuv0HJHUhfxB5FD9icI0Xf7R6%2BB6IaqHY3Qdva2aAkTRRiyBzHF6dtz3cP7XOjQZSuA6H6FUEoU8hApXnnpr3OzWldZ%2Be6hp0Ldt3eSffo0eCwdwIH78sgQBBCXJNx1ahDgkQKiTJOhTBvXlwPdD4eC6QDSRcLiggqQ3GGFJ%2FRkPTIXpVSxKIIxCqrJCHuL%2BPqIsQ9FPdTCtIyJ4jgZrzresQ9SK%2BFvw%2B3jRtOo11ztifmahmL9BD3Xa29PC5keB5JBIaXvWVKj9%2Bs320KqXUubolMokhesr3jLjtGdFD5R6IKnnohXpRtYRqcqyifiC8lsyuOlMfB7Grtlkz4WPQpSNCTEOioD6d4c7sWRijB9mdk6r8c5VT6fgI6P0blpx4W8ImMpqMjJcbyXMRRl6caVnpYZ0d1Hx1DrC1Si2ORZs%2FsgfE%2BOkwsz1bR%2BuA8sqT%2BMU8LugC97dQPPvGkr6%2FWXvF8JS9cipWd9pbbuuvWGo2dnZrbqrW20ehG8XP8r%2BMfy9k3b8LHTxpuu%2B00tneaTqgSgVYlLs8ez1%2F6yUT6YaPV3kWFbYBDj3%2FyxNjduO0Jam6r5uCfC0vF4xyWGol4UgZH4cVbRqF4Tyo4iHURlEiCMxRllaeeeJRGl70wyxJ9b2trNPGQ4vhUNvT6SKKv%2BJ49VEd92PJUTim6lfjB1gqNxzofZdOEjHhGpcaLxBCPtxC9QgUPZexXXwMsRNEUtJGHcuHvTbt0w2uv%2FjxLRZFm%2FmMAFG3ef7GIuAfLO%2FS9ooZRBsxS7uPnv6DEKoKZnio4I8hUgvqNmHolANhAbEKUkfR5w9hNDZQHuAHu6KpIGRFIEfmWZOTuGJqn%2B8KTmnKzpMLHYOg6DadtCC6%2FiVTgNQ4PTzVgB5WAxaQUpi79482PWr%2F%2FcG1tLYQVK5f57o7TstzuDGZtZ5OdpmpUFjri%2Bwj3p5RiGWz6qavUbDq7Voj1aoWlizuzmoeScyw4Gpt5LNX63S%2FP3v%2FM1Go2l9JYpRqtlv18%2BbV9ESWhZEPhhbF8CjQJXcp6Viry4R%2Ff%2F94SRZpN1wpy%2BXW6xZQ9I%2FqpghamMhbX%2BpQcX%2FBypN6dMvVQQOoqHa9KqTOC6N9HLMMltxah%2B5fn0P3VHw0WIsuIgZwuQxDNAp0PQJ1n3PZl5KQ8l34%2BR8uUMWOBQk1on5ewIwLgNPAZjG3DBhmQt0DLzg1EQtcf5%2FgWochbyA0QUcoBACchmQCOMUTMA5mES%2BIiKTJ%2BJkIV%2BdQoYfpQ8CgLa0AVhrdfBCX4PlAXhH03TZ%2BbtUcUaeS1YQWUY4KnKquexiCnDJQxtEFwQhBfoPT6BK%2BJVeXYRcgzM8OYwYUMZvotFDamRJm0bQ5D0RMgbQh1jPggkLGJI1NA3pscKwj7prk3IQCE4TdmKbwyCBp4ANdQy940gxEZhLITwiwWkiXuhiATLaJzughAkpj5OSzCC8QGNEiH4EXpL7EX7E0vNqvKLDcCULtGqQVAoocllq8gRIKOdg4jo5Ho1p0l8gFPOk%2Bjwew4jaH%2BOaAwmp1VFH3xxhsqDcwLOeUDRglTUeigRi8rZ4CJAuwKUoEcN1GQAwvBmgpjK7UK0x%2Buu27RY4uW31w0fZ0NEoQt4Xd40ypo7wGk0VCP8LZB8NCOspmtnEF1dQYtomuALhmP8SsuQV%2B%2FOpn3rg3Cg9mkxw4SXD0pqtT6YNjv7R1srJ6drdxdSIxihrIBmVGtY5%2BnvjHt%2FNKhmc7ZOhDzoNcfbtiTb3OfI71JgZKVF2IwQGmoTs9kGIZ5qa%2F2GlaaF60ALKdTAxPIinMhDmlkNOcrxbBfyY7r%2FXj9tH84OO7MrdqZQ3qIdkxlCyPbMmYQtnM8GHb6DWhq2qB5qp1iWjAC3AiGxw86Q3Y4YJ2j%2FkFn%2Fwfs4Unv8cmb98z493kpY3v%2Fa8WrNPrJwWOY%2Bv%2Bi6Jz2hgcnw8POETs8Pu10hxRhFZ2xOcLagRZupGhGcBxpkyjEhR1Q2SiVIii6NIQjo2YysIgmj6lBU%2F5btUh1I8Nq%2FSgVrIJlnarouWwDdS047XqkdBHFPBaJRZ2UKROXCUxe7A%2FK4ZjuMfsxWO8asxmcKIYQqsTFyskvJnsYIQWCtW3OTrW6gIz%2F3ejs1t3W9dH5tVMoMuGeyAuMOAPBxcz81f9kZr6az8y%2FuTYz%2F3Rxsh0f5R6sJIs5uBtxeEGtmoNDoE%2FMb1pY8uBEeiriK6lPVAbrwIqWeQdm1nIluT%2BAIAXjt7lgh9EqyvF9isOokCK8LxWyMJYr5biPBvIMba6Q49FkhDFipZLBQE01epSV5PS3v47R4FbQPoFOlzLWsGFB30Ouynzl0mEwwVbNknpHWPaspFPRpGApsHHwCmmLlcPfKiuHhKYFx2lsJTqrY9jcqewaPpDudnO35radVvnLbZitQ9PZubvdarXbO5Wtw%2Fyl3To0G7vN7db27rKtQ8OpOeC3%2Fe%2B3DuIxXDX9NPYBtz9aug64XSvSKLct%2B4XFxZSBj5%2F%2FrCwE94ouvjUfWMr%2BaSoZMhd4AkV%2Bxq6y6jNl7fO3PPjrP3%2F42S0PGo694xsHKD1AbXSF%2FSlQiNAP6Y63ZMxxstByJtf2AzPrFRhrdhAaHtjZkua%2BTfbO3LFHMhCl2n%2F506%2FYErXxp4liJn99JpIRoirVgqZWIlUKtHSfYH2TFjSwY0DRBxGtXu7dVqHXt26wma9HjRwAMka1UpE%2F%2Fz1dulxoO4VtX0cBND1t8erKXoEt2ytUK8W1hcK%2FAA%3D%3D). This rigorous appraisal supports the reliability of the pooled prevalence estimate of **6% (95% CI: 2.1%–11.8%)** reported in this study.
